# Protocol for a controlled human infection with genetically modified *Neisseria lactamica* expressing the meningococcal vaccine antigens NadA and FHbp: *The GM-Nlac Pilot Study*

**DOI:** 10.64898/2026.11.06.26367448

**Authors:** Giles Chick, Diane F. Gbesemete, Pamela Jump Salcedo, Matthew W. Wilson, Alison R. Hill, Tyween J. Coutinho, Rebecca Anderson, Ryan S. Elliott, Dinesh Mohanraj, David W. Cleary, Jay Lucidarme, Ray Borrow, Saul N. Faust, Andrew R. Gorringe, Robert C. Read, Adam P. Dale, Jay R. Laver

## Abstract

**Introduction:** Previous work has shown that experimentally induced colonisation with *Neisseria lactamica* (Nlac) is safe and associated with reduced meningococcal colonisation. This pilot study aims to establish the safety, induction of colonisation and immunogenicity following intranasal inoculation with a mixture of four strains of genetically modified Nlac (GM-Nlac). Each of these strains express meningococcal vaccine antigens present in current vaccines: *Neisseria* adhesin A (NadA) and one of four different variants of Factor H-binding protein (FHbp), the latter of which is specifically mutated to abrogate its ability to bind the human complement protein, Factor H.

**Methods and analysis:** In this controlled human infection model experiment, 10 healthy participants aged 18-45 years will be inoculated with a mixture of 4 strains of GM-Nlac (collectively referred to as the challenge agent, 4xrNlac). They will be followed up over 28 days to assess colonisation, immunogenicity and efficacy of eradication with a single oral dose of ciprofloxacin (500 mg). Participant contacts will also be recruited to assess for onward transmission of GM-Nlac.

**Ethics and dissemination:** This study involves the deliberate release of four genetically modified organisms, which has been approved by the Department for Environment, Food and Rural Affairs (reference: 25/R50/01). The clinical study has also been approved by South-Central Oxford A Research Ethics Committee and the Health Research Authority (reference: 26/SC/0072). All participants will provide written informed consent prior to enrolment. Findings of the study will be submitted for publication in a peer-reviewed scientific journal and for presentation at scientific conferences.

**Registration:** This study is registered on the ISRCTN registry with study registration number ISRCTN10930766

**Strength and limitations of the study:** *Strengths:*

- A platform controlled human infection model that will enable the study of human mucosal immune responses to single gene products of *Neisseria meningitidis*.
- A pathfinder study that will guide potential future mucosal immunoprophylaxis.
- Rigorous protocol for safety evaluation of a deliberately released multistrain genetically modified live bacterial product.

*Limitations:*

- Small exploratory pilot study.

## Introduction

### Meningococcal disease and vaccination strategies

The development of invasive meningococcal disease (IMD) requires prior nasopharyngeal colonisation with *Neisseria meningitidis* (Nmen), which is governed by complex interactions between the organism and its host [1]. The majority of meningococcal colonisation events remain asymptomatic, with only a minority progressing to invasive disease. Nmen colonisation prevalence varies by location and age. In high-income settings, between approximately 2.5% to 12% of the population are colonised, increasing from very low prevalence in infants, to a peak of approximately 25% in some demographics of young adults [2–4]. This surge in colonisation prevalence is associated with increased IMD incidence, particularly amongst university and college students [5]. Sadly, such outbreaks continue to occur; in the UK an outbreak of serogroup B meningococcal disease in 2026 was associated with 21 confirmed cases including 2 fatalities [6].

Glycoconjugate meningococcal vaccines such as the quadrivalent ‘ACWY’ have dramatically reduced the incidence of IMD in a serogroup-specific manner. Such vaccines provide individual protection against IMD through induction of serum bactericidal antibody (SBA) activity, but also contribute to herd protection by reducing the acquisition of asymptomatic nasopharyngeal colonisation, thereby interrupting transmission [7]. However, for serogroup B meningococcal strains, conjugate vaccination strategies are precluded by poor immunogenicity of the polysaccharide capsule, which results from structural homology between the capsular polysaccharide (α-2,8-polysialic acid) and neural carbohydrates [8]. Vaccines for serogroup B meningococcal strains have therefore focused on utilisation of surface exposed, sub-capsular proteins, with two such products currently licenced (4CMenB and MenB-FHbp). While both provide individual protection from IMD via induction of SBA, they do not impact upon acquisition of asymptomatic colonisation and are therefore unlikely to induce herd protection [2,9,10]. In keeping with the World Health Organization’s mission to defeat meningitis by 2030, alternative strategies are required to interrupt the asymptomatic colonisation state for serogroup B meningococcal strains in order to induce herd protection, noting that the meningococcus resides in a strictly human niche [11,12].

### Exploiting *Neisseria lactamica* to reduce meningococcal colonisation and disease

The human pharynx is colonised by a wide range of bacterial species which, over time and repeated exposure, provide natural immunity against diseases caused by opportunistic pathobionts such as Nmen [13–15]. *Neisseria lactamica* (Nlac) is a non-capsulate, non-pathogenic, obligate human commensal that is closely related to Nmen. Like its pathobiont cousin, the exclusive reservoir of Nlac is the human upper respiratory tract. Nlac colonises a high proportion of young children (up to ∼ 60-70% in some cross-sectional surveys) [16–18], but colonisation frequency wanes through adolescence into adulthood, where it is encountered only rarely. Age-specific rates of meningococcal colonisation and disease inversely correlate with Nlac colonisation prevalence, and a causative relationship has been demonstrated between Nlac colonisation and protection against meningococcal colonisation in the controlled human infection (CHI) setting [16,19,20]. While the mechanism(s) underpinning this effect remain incompletely understood, mathematical modelling predicts that the protective effect of Nlac against meningococcal colonisation persists for several years, suggesting immune-mediated component [21].

In support of this hypothesis, we have previously demonstrated that experimental colonisation with Nlac induces systemic adaptive immune responses, including Nmen-cross-reactive functional (opsonophagocytic) IgG, IgA/IgG-secreting plasma cells, and IgG memory B cells [22,23]. Indeed, Nlac and Nmen are closely related at the genomic level, and both species express many similar, surface-expressed proteins that likely form the basis of observed immunological cross-reactivity [24]. Notably, colonisation with wild-type Nlac does not reliably induce cross-reactive anti-meningococcal SBA, the defined immune correlate of protection for IMD upon which new meningococcal vaccines are licenced. However, when considering the development of novel strategies to protect against meningococcal colonisation, generation of SBA may be redundant, noting that currently licenced serogroup B meningococcal vaccines induce potent SBA but offer no protection against nasopharyngeal colonisation with serogroup B strains [10]. On the contrary, experimental data pertaining to immunological studies of other upper respiratory tract pathobiont/pathogenic bacterial species, for example *Streptococcus pneumoniae* and *Bordetella pertussis*, suggest that protection against nasopharyngeal colonisation is mediated via resident mucosal adaptive immune responses, specifically via induction of IL-17^+^ CD4^+^ T cells and/or mucosal secretory IgA responses [25–29].

### *Neisseria lactamica* Controlled Infection Model: Safety Considerations

Nlac is a strict commensal species of the upper respiratory tract and is non-pathogenic in the immunocompetent host. Rare case reports of invasive disease caused by Nlac are described but such cases are restricted either to patients with bony injury of the skull, which facilitates contiguous spread of this organism from the upper respiratory tract to the cerebrospinal fluid, or to those with severe immunocompromise [30].

To date, seven studies have been performed utilising wild-type Nlac (strain Y92-1009), including one study performed in late pregnancy [20,22,23,31–33]. In addition, one further study involved inoculation of participants with genetically modified (GM) derivatives of Nlac strain Y92-1009, each either expressing the meningococcal vaccine antigen, NadA (strain 4NB1), or a silent construct that rendered the GM strain effectively wild type with respect to gene content (strain 4YB2) [34]. ln all previously performed studies, inoculation was via the intra-nasal route using inocula prepared from frozen or lyophilised inoculum stocks. Doses of Nlac utilised across the studies to-date range from ∼10^4^ - 10^7^ colony forming units (CFU). Dose-escalation studies performed in different settings have demonstrated variable kinetics relating to dose saturation. In the UK, an intra-nasal inoculum dose of ∼ 1x10^5^ CFU has resulted in a colonisation fraction of ∼90-100**%** [23,32–34], while in Mali the maximal attainable colonisation fraction was ∼65% despite dose-escalation to 10**^7^** CFU [33].

Across all eight previous studies (∼500 Nlac-inoculated participants in total), no serious adverse events associated with Nlac-attributable symptoms or signs have been reported following Nlac inoculation. Furthermore, in studies that were placebo-controlled, there has been no evidence that inoculation or colonisation with Nlac has induced adverse clinical symptoms in the respiratory tract or elsewhere above the frequency observed amongst placebo recipients [20,23].

In nature, Nlac is transmitted amongst close household contacts, with evidence suggesting that transmission may be more common from children to adults [32]. Although data are limited, factors associated with increased likelihood of Nlac colonisation include increasing number of people per bedroom, presence of runny nose, and young age [18]. However, in our previous GM-Nlac study, bedroom sharers of study contacts were enrolled but no detectable transmission events occurred (*n* = 8 contact participants across a 90-day study period) and no environmental shedding of viable GM-Nlac bacteria was detected by culture of mask and exhaled air samples, routinely collected from all colonised participants [34].

Importantly, Nlac strains utilised across our CHI studies remain exquisitely sensitive to first line antibacterials that may be utilised to treat disease caused by *Neisseria* spp., including susceptibility to third-generation cephalosporins and fluoroquinolones. This significantly mitigates the risk to participants should rescue antimicrobials be considered necessary by clinical investigators. Consistent with the broader meningococcal literature, single dose oral ciprofloxacin (500mg) has been demonstrated to eradicate wild type or GM Nlac colonisation in 100% (*n* = 31) of experimentally*-*colonised participants in studies where antimicrobial eradication therapy was attempted [23,34].

### A multi-strain, genetically modified *Neisseria lactamica* challenge agent: 4xrNlac

The established safety, colonisation characteristics and antimicrobial susceptibility of Nlac provide a platform through which defined determinants of immunity elicited in response to nasopharyngeal colonisation can be investigated experimentally. In the current study, GM-Nlac strains have been developed to express two meningococcal surface antigens: Neisseria adhesin A (NadA) and factor H-binding protein (FHbp).

NadA is a type V trimeric autotransporter expressed by a relatively restricted subset of meningococcal strains. Expression of the *nadA* gene is negatively regulated by the NadR repressor protein and is subject to phase variation. It is a component of the licensed four-component serogroup B meningococcal vaccine, 4CMenB, and induces functional antigen-specific antibody responses [35,36]. Although the receptor for NadA binding has yet to be formally identified, eliminating NadA expression in Nmen strains reduces the capacity of knock-out mutants to bind epithelial cell lines [37]. Previous work has demonstrated that NadA can be functionally expressed by GM-Nlac and be safely delivered to the human nasopharyngeal mucosa. Sustained, prolonged nasopharyngeal colonisation with NadA-expressing GM-Nlac leads to induction of NadA-specific IgG-secreting plasma cells, increases in the proportion of circulating NadA-specific IgG memory B cells and, in a subset of colonised participants, increases reciprocal SBA titres against a NadA-overexpressing strain of Nmen (strain 5/99) from <4 to ≥4, consistent with protection against IMD caused by that strain [34]. Note that in the current study, each strain expresses the NadA variant, NadA-1.

FHbp is a surface-expressed lipoprotein produced by the large majority of circulating meningococcal strains, although its sequence and antigenic properties vary considerably between isolates [38]. The only definitive function of FHbp is its ability to bind human complement regulator Factor H (hFH), thereby reducing complement activation on the meningococcal surface and enhancing bacterial survival in human serum [39]. FHbp is included in both currently licensed serogroup B meningococcal vaccines and is an established target of serum bactericidal antibodies [40]. The family of FHbp proteins resolves phylogenetically into two subfamilies (A and B), which can be further subdivided into three variant groups (1, 2 and 3). Whilst Subfamily B contains only variant group 1 *fhbp* alleles, subfamily A is comprised of *fhbp* alleles from variant groups 2 and 3. Variants within subfamilies are more antigenically similar to one another than they are to members of the other subfamily, such that antibodies raised against variant 1 FHbp are only mildly cross-reactive with FHbp from variant groups 2/3, and vice versa. Therefore, to represent the antigenic diversity of FHbp in this experiment, the bacterial inoculum consists of four otherwise-closely-related GM-Nlac strains, each one expressing a genetically attenuated version of one of the following four FHbp variants: 1.4, 1.13, 2.19 or 3.45. These variants are among the most prevalent FHbp alleles in invasive meningococcal strains circulating in the UK [38]. As a precaution against potentially introducing gain-of-function (*i.e.* hFH-binding) into Nlac, each heterologous antigen has undergone site-specific mutagenesis to reduce its ability to bind hFH. Multiple reports have identified amino acid residues specific for the interaction of FHbp variants and hFH, which have been demonstrated not only to attenuate hFH binding [41,42], but to enhance the immunogenicity of FHbp in animal models [43,44]. Additionally, in the case of variant group 2 FHbp, a pair of specific mutations have been identified that simultaneously disrupt FHbp-hFH interactions and thermally stabilise the relatively heat-labile N-terminal domain of the protein [45]. These thermostabilising mutations were utilised in this project. The FHbp variants expressed in this study are therefore: FHbp1.4(H248L), FHbp1.13(H248L), FHbp2.19(L130R G133D) and FHbp3.45(E254A).

The alleles coding for these proteins were rationally designed, synthetic genes which are designed to be as different from one another as possible at the nucleotide sequence level. This diversity has enabled the design of bespoke primer/probe combinations for discriminatory quantitative PCR (qPCR) assays. Transcriptional control of *fhbp* expression is driven by the same UAS-enhanced, hybrid *porA/porB* promoter as used in the previous GM-Nlac study [34]. All four strains challenge strains contain identical, Nlac-codon-optimised *nadA-1* alleles, expression of which is driven by a second copy of the hybrid *porA/porB* promoter. Due to the endogenous expression of the *lacZ* gene (coding for β-D-galactosidase) by wild type Nlac, our cloning strategy exploits the activity of this gene product to screen for new transformants. Because β-D-galactosidase activity is a key diagnostic criterion for identification of Nlac, allowing Nlac to grow as blue coloured colonies on bacteriological agar containing 5-bromo-4-chloro-3-indolyl-β-D-galactopyranoside (X-Gal), we had to ensure all resulting GM-Nlac strains retained β-D-galactosidase activity. As a result, each strain was transformed with a synthetic, rationally-designed copy of the *lacZ* gene. All nucleic acid constructs are stably incorporated into an intergenic locus in the Nlac chromosome (NHCIS1).

Extensive pre-clinical characterisation of each strain has been conducted, the results of which were submitted to the Advisory Committee on Releases to the Environment (ACRE) as part of our successful application for permission to conduct a Deliberate Release of these genetically modified organisms (GMOs) from the Department for Environment, Food and Rural Affairs (DEFRA) (https://www.gov.uk/government/publications/genetically-modified-organisms-university-of-southampton-25r5001). Briefly, we demonstrated: (i) surface co-expression of NadA and FHbp variants, (ii) attenuation of hFH binding to GM-Nlac strains expressing specifically mutagenised, as opposed to wild type FHbp variants, such that each of the 4 challenge strains bound only a similar amount of hFH as did control GM-Nlac strain, 4YB2, (iii) that each of the 4 challenge strains remained acutely susceptible to killing by 50% (v/v) native human plasma, (iv) that each of the 4 challenge strains remained acutely susceptible to frontline, clinically relevant antibiotics: ciprofloxacin, rifampicin and ceftriaxone, (v) that there were no changes to the internalisation of the 4 challenge strains by a human epithelial cell line, suggesting that expression of the heterologous antigens was unlikely to provoke a radical shift in the dynamics of infection nor the typical infective life course of the bacteria following challenge, (vi) that the genetic modifications to the Nlac chromosome were targeted to the correct locus, and that each strain contained only very minor nucleotide sequence differences (consistent with, for example, mechanisms of phase variation) from the wild type precursor Nlac strain, and (vii) that each strain remains refractory to transformation with exogenous nucleic acids derived from serogroup B Nmen strains.

The four strains have been combined into a single multistrain inoculum, enabling concurrent evaluation of immune responses to epidemiologically relevant FHbp variants spanning all three variant groups within a controlled colonisation and homologous rechallenge model. The ratio of GM-Nlac strains: 4NFA313_H (expressing NadA and FHbp1.4(H248L)), 4NFB313_H (expressing NadA and FHbp1.13(H248L)), 4NFC313 (expressing NadA and FHbp2.19(L130R G133D)) and 4NFD313_E (expressing NadA and FHbp3.45(E254A)) within this multistrain inoculum was empirically determined to be 1.05 : 1.00 : 2.33 : 1.69, respectively by colony genotyping of *fhbp* alleles.

### Study Rationale

This study aims to establish the safety, colonisation potential and immunogenicity of nasal inoculation with a single multistrain inoculum containing four GM-Nlac strains, each expressing NadA and one of four FHbp variants.

Data generated from this pilot study will inform the development of a subsequent randomised, double-blind, placebo controlled clinical study to determine whether colonisation with GM-Nlac induces sterilising protection against homologous rechallenge. If GM-Nlac colonisation induces homologous protection, subsequent studies will assess its impact on Nmen colonisation and determine whether GM-Nlac could be utilised to induce herd protection against Nmen by depleting the strictly human colonisation reservoir.

## Methods and analysis

### Study overview

In this prospective, single-arm, open label, controlled human infection model (CHIM) experiment, 10 healthy adults, enrolled as “challenge participants”, will be challenged with 4xrNlac, administered as a nasal inoculum. Challenge participants will be followed up over 28 days to assess safety and nasopharyngeal colonisation with the four GM-Nlac strains, and to assess immunogenicity. Any individual who regularly shares a bedroom with a challenge participant will be enrolled as a “contact participant” and will be reviewed at day 28 post challenge of the challenge participant to assess transmission of the GM-Nlac strains. All challenge participants, and any colonised contact participants, will then receive a single dose of oral ciprofloxacin to clear GM-Nlac colonisation, with a final visit to confirm clearance. All visits will be conducted in the NIHR Clinical Research Facility located within University Hospital Southampton (see **Figure 1** for study schedule).

**Figure 1.**
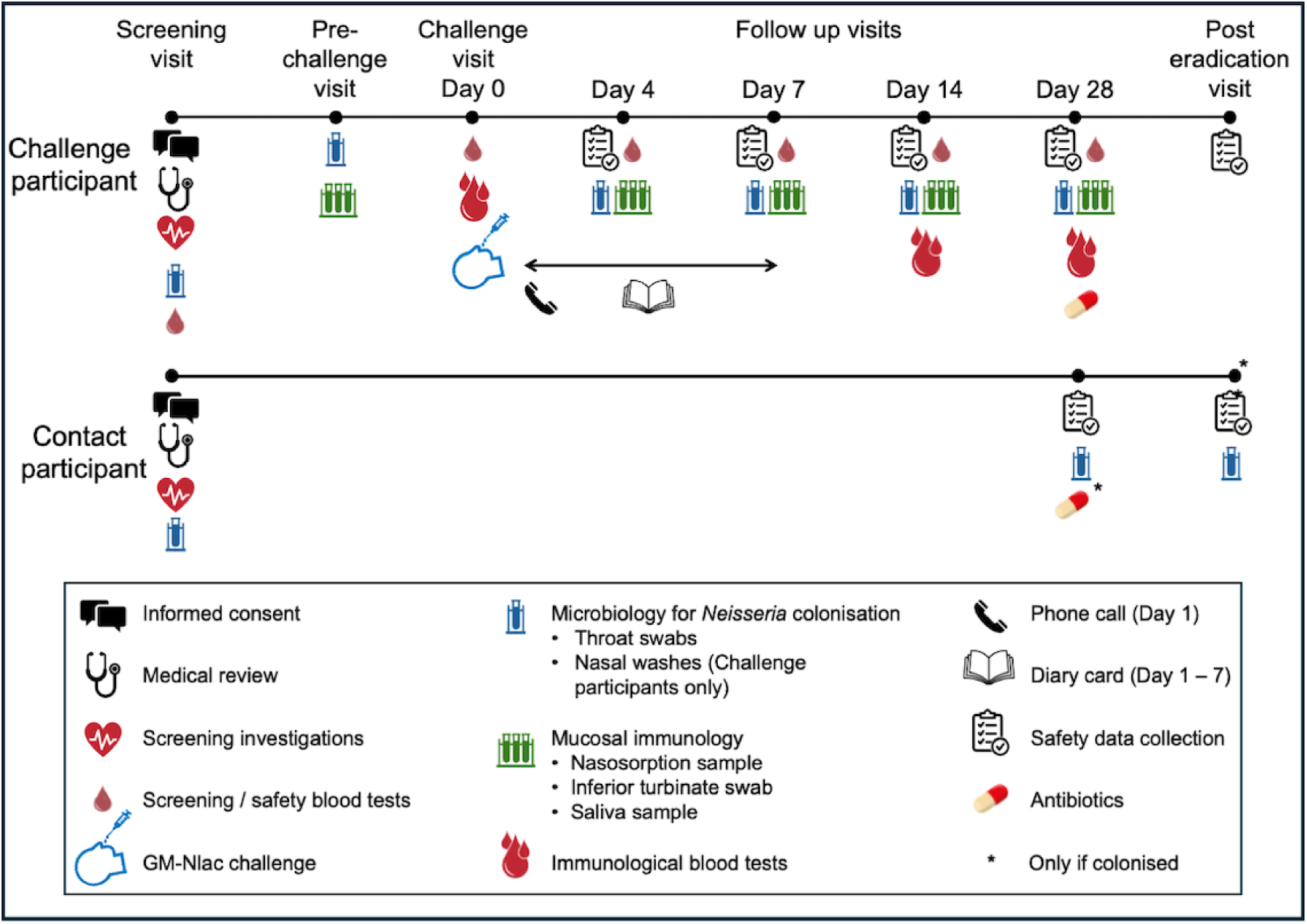
– Study overview. Schematic illustration of the study schedule for challenge and contact participants. All participants will be given a 24-hour telephone number to contact the study team if any safety concerns arise between visits, and additional visits and investigations will be arranged if clinically

This study aims to establish the safety, colonisation potential and immunogenicity of nasal inoculation with 4xrNlac, to assess for transmission to close contacts, and to demonstrate the efficacy of a single dose of oral ciprofloxacin in clearing nasopharyngeal colonisation. Study objectives are detailed in **Table 1** and study endpoints in **Table 2**.

**Table 1.**
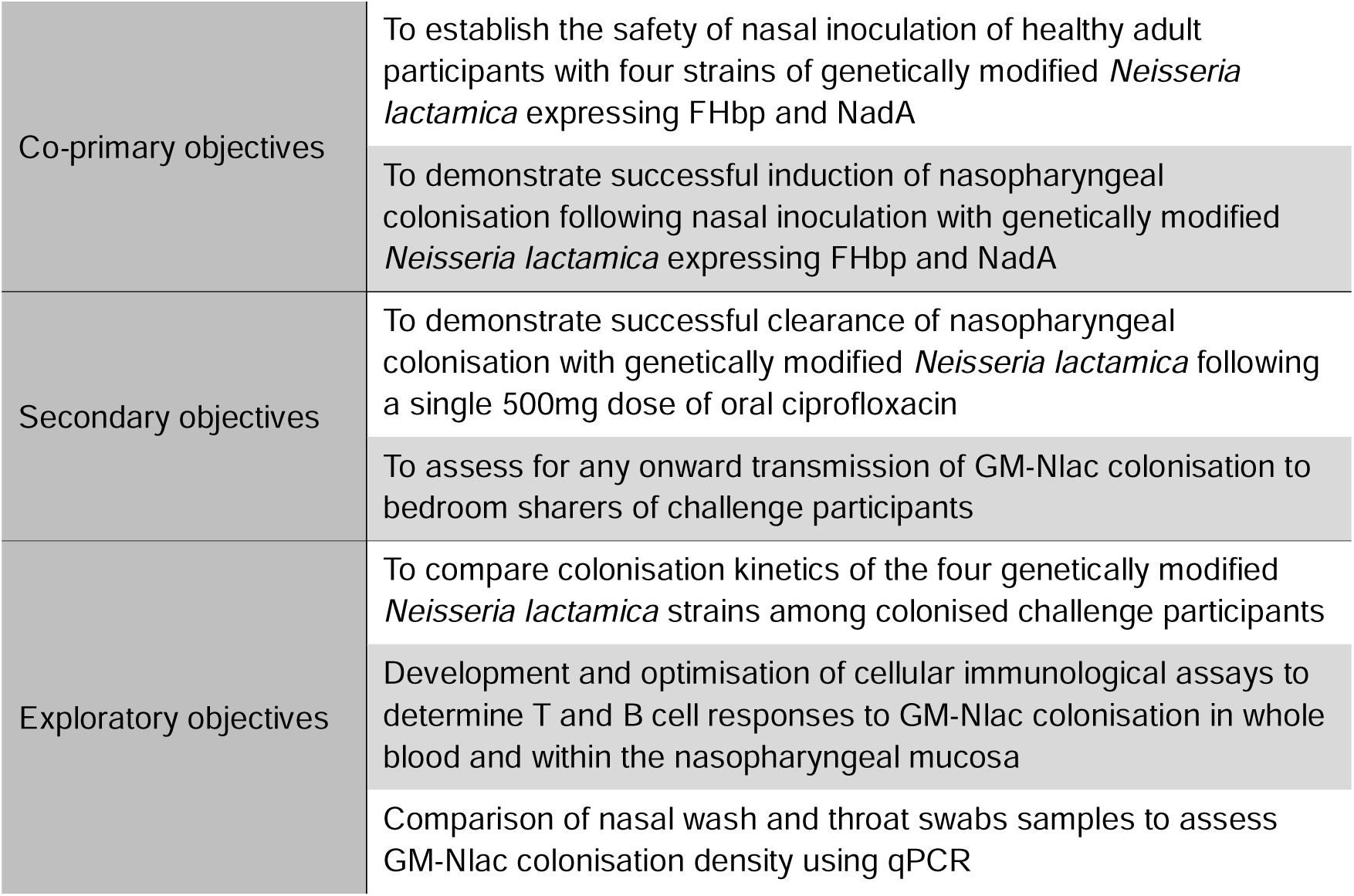
– Study objectives.

**Table 2.**
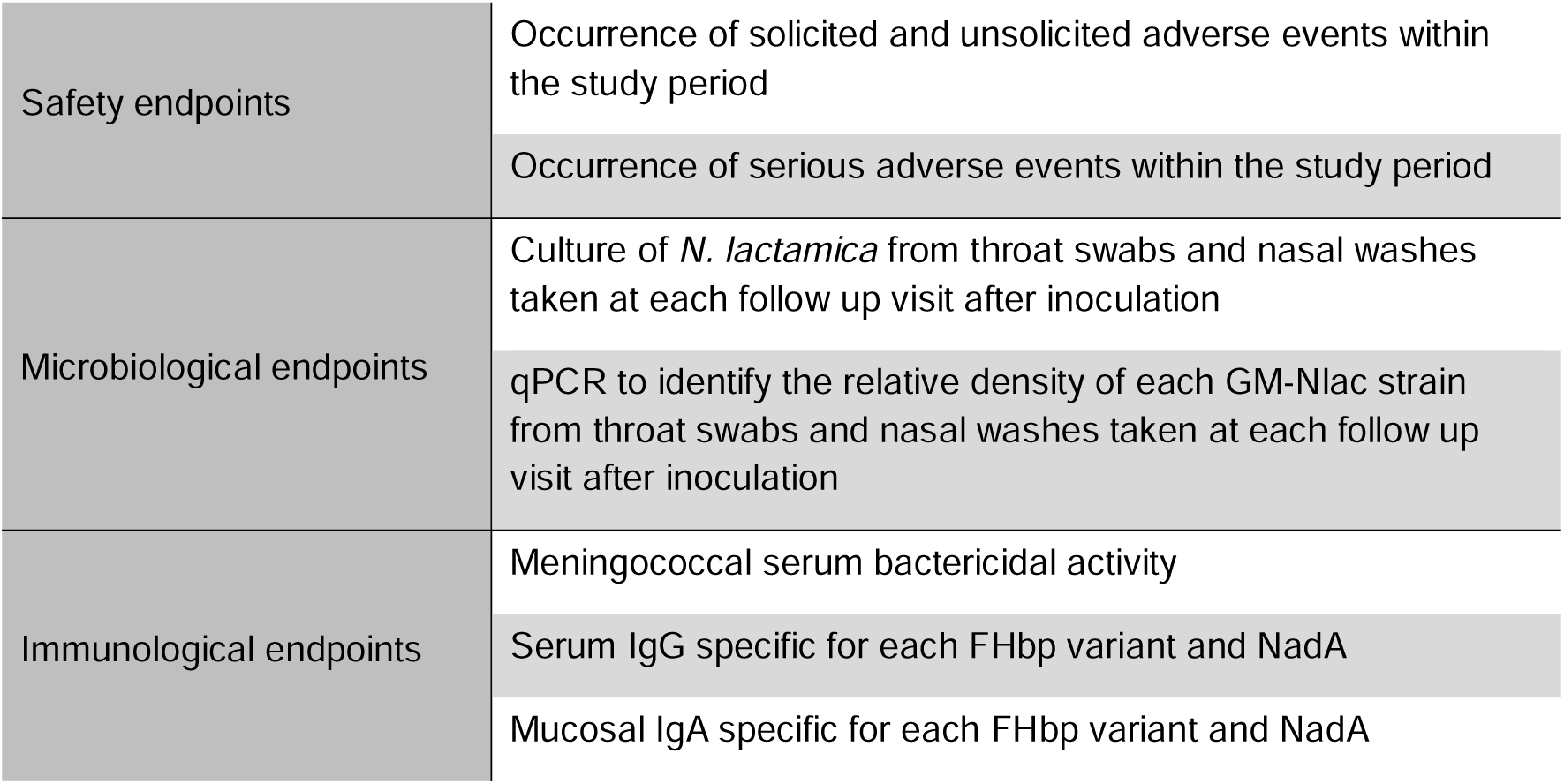
– Study endpoints.

|  |  |
|---|---|
| Safety endpoints | Occurrence of solicited and unsolicited adverse events within the study period |
|  | Occurrence of serious adverse events within the study period |
| Microbiological endpoints | Culture of *N. lactamica* from throat swabs and nasal washes taken at each follow up visit after inoculation |
|  | qPCR to identify the relative density of each GM-Nlac strain from throat swabs and nasal washes taken at each follow up visit after inoculation |
| Immunological endpoints | Meningococcal serum bactericidal activity |
|  | Serum IgG specific for each FHbp variant and NadA |
|  | Mucosal IgA specific for each FHbp variant and NadA |

### Safety and infection control considerations

Although pre-clinical testing has demonstrated no potential for increased virulence or transmissibility of the GM-Nlac strains, strict safety and infection control measures will be adhered to throughout the study to ensure the safety of participants and to minimise the potential for onward transmission to study team members, other contacts of participants, and the wider community. Eligibility criteria for both challenge and contact participants will exclude individuals who are at increased risk of disease, and those who have close contact with any individuals who are vulnerable or have higher transmission potential (see **Supplementary Tables 1 and 2** for full eligibility criteria). Bedroom-sharers of challenge participants, who are at the highest risk of onward transmission, will be invited to participate as contact participants. A maximum of one contact participant will be enrolled per challenge participant, and if a bedroom-sharer is unwilling or ineligible, the challenge participant will not be eligible to participate.

As 4xrNlac has not previously been administered to human participants, enrolment will follow a first-in-human process with sentinel participants and a safety review prior to further challenges. Challenge participants will be monitored closely for safety concerns during the first seven days following challenge with a day 1 telephone contact, daily diary card for seven days, and face-to-face visits on days 4 and 7 post challenge. This period corresponds to the period of highest risk of meningococcal disease following Nmen colonisation. Throughout the study, challenge and contact participants will be encouraged to contact the study team via a 24-hour telephone number if they become unwell. In the event of any participant developing symptoms suggestive of respiratory or systemic infection, additional visits and investigations will be conducted at the discretion of the investigators, and early antibiotic clearance (single-dose) or treatment course administered if the study team suspect it to be related to GM-Nlac colonisation. In the event of systemic illness suggestive of GM-Nlac bacteraemia or sepsis, the participant will be referred to the acute medical services in University Hospital Southampton.

The study will be paused for a safety review in the event of any participant developing a respiratory or systemic infection assessed as related to GM-Nlac and requiring a treatment course of antibiotics. Additionally, if early antibiotic clearance therapy (single dose) is given due to safety concerns to two of the first five participants, or more than three participants overall, a study pause will be triggered. An independent Data and Safety Monitoring Board (DSMB) will oversee study conduct and review all safety data. Hampshire and Isle of Wight Health Protection Team South-East will be informed when the study commences. Study participants’ GPs will be notified about their inclusion in the study, and any occurrence of disease in participants will be notified to the DSMB.

The GM-Nlac inocula and all clinical samples potentially containing any GM-Nlac bacteria will be handled within Class II microbiological safety cabinets, and clinical procedures including inoculation will be performed within an environmental chamber. Study personnel will be trained in handling GM-contaminated materials and will wear appropriate personal protective equipment. Any potentially GM-contaminated materials will be disposed of according to study-specific operating procedures.

### Recruitment and screening

Study participants will be recruited using a wide range of ethically approved methods across both traditional and digital platforms. Challenge and contact participant information sheets will be provided to interested individuals, which will include full details of the study requirements, risks and safety measures.

Potential challenge and contact participants will have a face-to-face medical screening visit to ensure fully informed consent and to assess eligibility. This will take place up to 90 days prior to the challenge visit and will include a detailed medical history, physical examination and screening investigations as detailed in **Supplementary Tables 3 and 4**.

Challenge participants must be healthy, non-smoking adults aged 18-45 years with no baseline colonisation with Nlac or Nmen, and no history of serogroup B anti-meningococcal vaccination. Contact participants will be defined as healthy adults aged 18-55 years who regularly share a bedroom with a potential challenge participant. All participants must demonstrate understanding of the study by successful completion of a pre-consent questionnaire, must provide written informed consent including agreement to abide by infection control measures throughout the study, and must have no regular contact with vulnerable individuals who may be an increased risk of disease or onward transmission of the GM-Nlac strains. The study will not recruit from vulnerable groups, or those who lack capacity to consent to the study. Full eligibility criteria are detailed in **Supplementary Tables 1 and 2** and the infection control rules are detailed in **Table 3**.

**Table 3.**
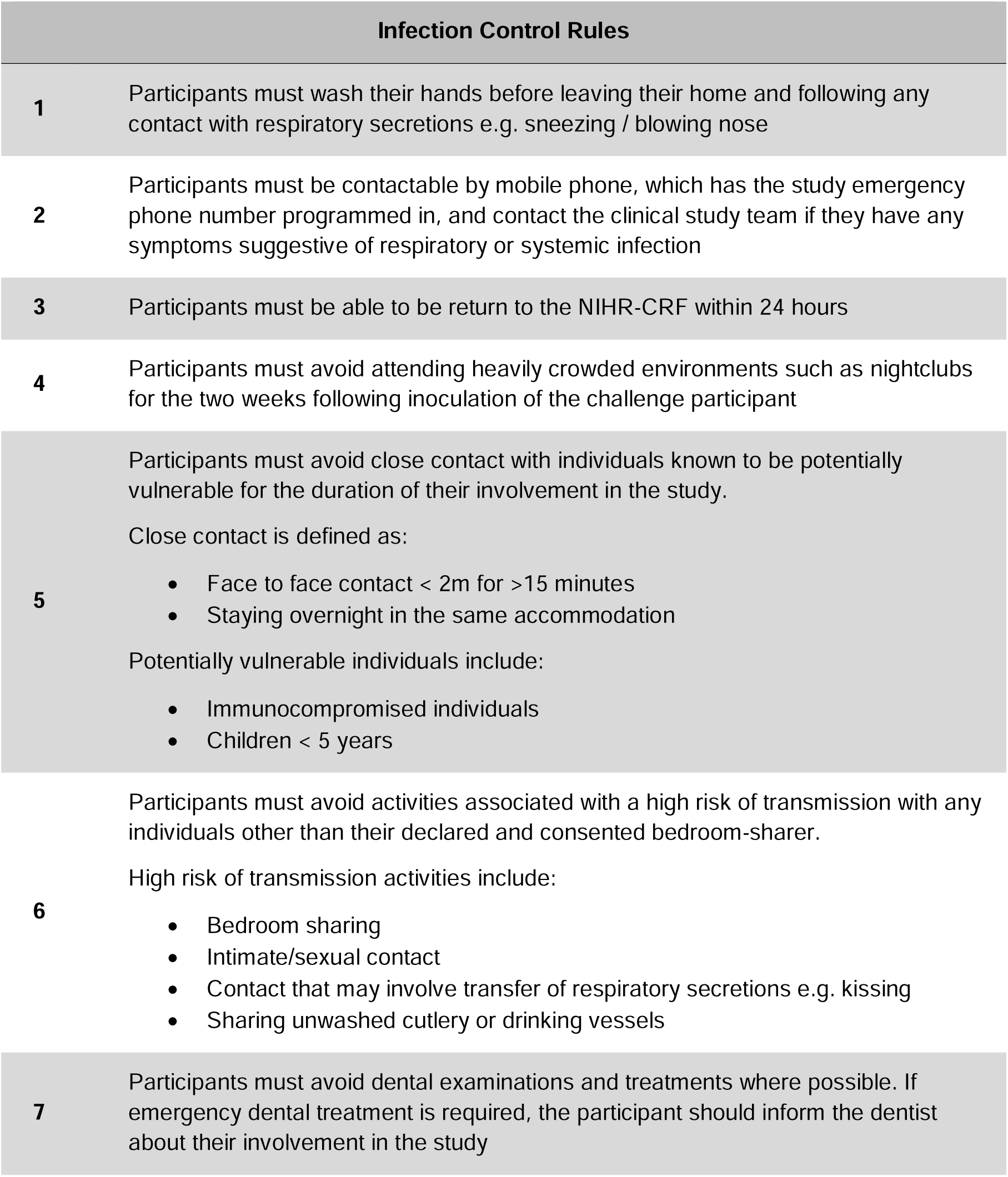
– Infection control rules. Infection control rules that both challenge and contact participants will be required to abide by from the time of challenge of the challenge participant, until completion of the challenge participant post-eradication visit

### Challenge with 4xrNlac

Challenge participants will attend for a pre-challenge visit five to 10 days prior to GM-Nlac challenge to ensure that they remain eligible. A throat swab will be taken to assess for acquisition of Nlac or Nmen colonisation since the screening visit, for which they would be deemed ineligible. Additional respiratory samples will also be taken to assess baseline mucosal immunological parameters. On the day of challenge, understanding, consent and eligibility will be re-confirmed and blood samples will be taken for baseline systemic immunological parameters.

Once eligibility for challenge has been confirmed, a dose of inoculum will be prepared from a single inoculum stock vial. The vial will be thawed and diluted in sterile phosphate-buffered saline (PBS) with dilution steps calculated using the known batch viability of the stock vials, aiming for an inoculum viability of 4 x 10^5^ CFU suspended in 1ml PBS. The inoculum will be transferred immediately to the participant and administered by a study doctor within 30 minutes of preparation, as previously described [34,46]. Briefly, the participant will lie supine with their neck extended and 0.5 mL inoculum will be administered per nostril via automated pipette. Following challenge, the participant will be asked not to swallow for one minute and then remain supine for five minutes.

Inoculum purity will be confirmed by culture of the inoculum prior to inoculation, and the administered inoculum dose calculated by viable count of the inoculum immediately prior to, and following inoculation. Some variation in administered inoculum dose is expected, but if an administered dose is found to be less than 4 x 10^4^ CFU/mL then this will be considered an inadequate inoculum and the participant may be replaced. If an inoculum contains >5 times the intended dose (i.e. 2 x 10^6^ CFU), it will be reported as an adverse event. Any inoculum >10 times the intended dose (i.e. 4 x 10^6^ CFU) will be classed as an adverse event of special interest and will trigger reassessment of inoculum preparation procedures and batch viability.

### Assessment of study endpoints

Study endpoints are detailed in **Table 2** and include safety, microbiological and immunological parameters. Challenge participant safety endpoints will be assessed by collection of adverse event data throughout the study. Solicited symptoms suggestive of possible respiratory or systemic infection (**Table 4**), any unsolicited adverse events and concomitant medication use will be recorded at the Day 1 telephone contact, via a paper diary card daily for seven days following challenge, and at each participant visit. Vital signs will be monitored at each participant visit and blood samples will be taken for assessment of haematological and biochemical parameters at each challenge participant visit to Day 28 post challenge. Adverse event severity, seriousness and relatedness to study interventions and/or colonisation with the GM-Nlac strains will be reported. Contact participant adverse event data will be collected at the Day 28 visit and any additional visits if required.

**Table 4.**
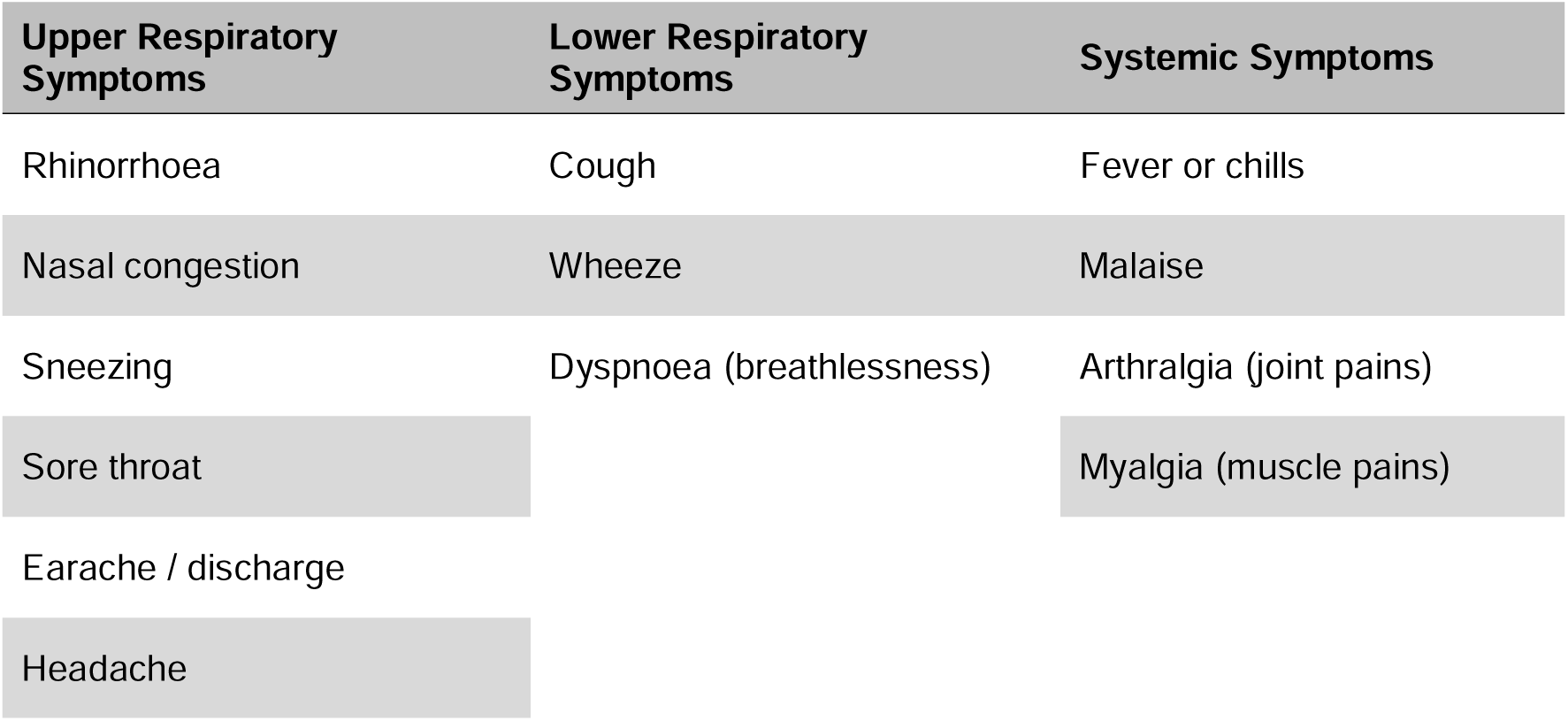
– Solicited symptoms. Solicited upper respiratory, lower respiratory and systemic symptoms which will be assessed in the diary card for seven days following challenge, and at each post-challenge study visit for challenge participants

Nasopharyngeal colonisation of challenge participants will be assessed by culture of throat swabs and nasal wash samples collected at all post-challenge visits, while transmission to contact participants will be assessed with throat swabs at day 28 and any additional visits if required. Positive colonisation or transmission will be defined as the detection of one or more colonies of any challenge strain in samples taken up to and including the day 28 visit. The composition of colonising GM-Nlac strain(s) will be determined by bespoke qPCR assay performed directly on participant throat swabs.

Baseline systemic immunological parameters will be assessed in serum and peripheral blood mononuclear cells (PBMCs) extracted from baseline blood samples taken immediately prior to challenge, with comparison to samples taken at Days 14 (PBMCs only) and 28 post challenge (PBMCs and serum). Comparison will be made between those participants who are colonised, and those who are not colonised with the GM-Nlac strains. For challenge participants only, mucosal immunological parameters will be assessed using: (i) nasosorption strips (for collection of nasal mucosal fluid), (ii) saliva samples, and (iii) inferior turbinate swabs, comparing baseline with post-challenge timepoints. Existing immunological assays will be optimised to detect responses against heterologous antigens throughout this study, for later use in the planned GM-Nlac randomised placebo-controlled study.

### Antibiotic clearance of GM-Nlac colonisation

A single 500mg oral dose of ciprofloxacin will be used to clear nasopharyngeal colonisation with GM-Nlac strains. This will be administered to all challenge participants under direct supervision at the Day 28 visit. Contact participants will be provided with a dose of ciprofloxacin to take home from the day 28 visit, and once their colonisation status is known, they will be instructed to either take or dispose of the antibiotic. Participants with any contra-indications to ciprofloxacin will be excluded at screening, and all female participants of child-bearing age will perform a pregnancy test prior to receiving the antibiotic.

Any participants who withdraw prior to the day 28 visit, or in the event of safety concerns, may receive early antibiotic clearance.

All study participants who have taken antibiotic therapy to clear colonisation with GM-Nlac will have a further visit 1-4 days after antibiotic administration to confirm clearance of colonisation. If a participant shows microbiological evidence of persistent colonisation despite receiving the antibiotic, they will be invited for a further visit for consideration of additional treatment by the study steering committee, with consultation with DSMB members where necessary.

### Withdrawal of participants

In accordance with the Declaration of Helsinki, a participant can withdraw from the study at any time, and is not obliged to give a reason for this. Similarly, a participant may be withdrawn by the study team if there are concerns surrounding their health, or if they do not adhere to study requirements. In the event of a challenge participant withdrawing from the study, their corresponding contact participant will also be withdrawn.

### Statistical considerations

Ten challenge participants is considered to be sufficient to estimate colonisation fraction and provide initial safety data. Therefore, no formal sample size calculation has been performed. The pilot study will also enable the optimisation and validation of immunological and microbiological assays for use in subsequent studies.

Safety endpoints will be assessed using data from all inoculated challenge participants and enrolled contacts, including both solicited and unsolicited adverse events, with comparisons between colonised and non-colonised participants. Microbiological and immunological endpoints will be evaluated in challenge participants deemed evaluable. Transmission analyses will focus on contact participants linked to colonised challenge participants, using their microbiological data.

### Patient and Public Involvement

A patient and public involvement workshop was held online in November 2025 to explore the acceptability of the proposed study and inform how it would be communicated to potential participants. Five public contributors, all healthy adults within the planned study’s eligible age range of 18–45 years, were recruited through the University Hospital Southampton Patient and Public Involvement and Engagement database and took part in a 90-minute facilitated session.

Contributors discussed the study rationale, participant journey, risks and benefits, infection control requirements and reimbursement arrangements. They considered the study and participant commitments broadly acceptable, while identifying areas requiring clearer explanation, particularly the rationale for safety and infection control measures and the role of contact participants. These questions led to the expansion and clarification of relevant sections of the participant information sheets. Contributors subsequently reviewed the information sheets and consent forms for both challenge and contact participants, together with the recruitment poster. Their feedback on terminology, document length and ease of navigation informed the final versions of the participant-facing materials.

### Ethics and Dissemination

This study has been approved by the Department for Environment, Food and Rural Affairs (reference: 25/R50/01), South-Central Oxford A Research Ethics Committee and the Health Research Authority (reference: 26/SC/0072). The study will be conducted according to the principles of the International Conference on Harmonisation Tripartite Guideline on Good Clinical Practice and will comply with applicable regulatory requirements. All challenge and contact participants will provide written informed consent prior to enrolment in the study.

Following completion of the study, findings will be disseminated by publication in a peer-reviewed scientific journal, and by presentation at relevant scientific meetings.

## Discussion

IMD remains a significant global health concern despite current vaccination strategies, with transmission underpinned by asymptomatic pharyngeal colonisation. While conjugate meningococcal vaccines reduce asymptomatic colonisation with subsequent induction of potent herd protection through interruption of transmission, currently licenced serogroup B vaccines do not impact upon this crucial endpoint [10]. Novel strategies are required to reduce asymptomatic Nmen colonisation to prevent asymptomatic transmission and associated IMD burden, particularly for serogroup B strains. Mucosal vaccination strategies, including those employing live-attenuated organisms, have shown promise in eliciting sterilising mucosal immunity for other upper respiratory tract bacterial pathogens, providing proof of principal that similar approaches may help overcome this limitation of non-conjugate meningococcal vaccines [26,47,48].

This study builds on over 15 years’ work where we have demonstrated that colonisation with wild type Nlac is safe and immunogenic [20,22,23,31–34]. Although colonisation with wild type Nlac protects against Nmen acquisition events throughout the active colonisation period, any immune-mediated protection conferred by natural Nlac colonisation is anticipated to be limited, as induction of Nmen-cross-reactive adaptive immune responses is not universally observed among Nlac-colonised participants [20,22,23]. Genetic modification offers a potential solution to this problem and, in the case of GM-Nlac, will enable sustained heterologous antigen delivery to the nasopharyngeal mucosa, with subsequent induction of resident adaptive immune responses. In support of this claim, in our first GM-Nlac CHIM, we demonstrated that colonisation with a NadA-expressing strain of Nlac (4NB1) was safe and immunogenic, eliciting anti-meningococcal SBA activity in ∼50% of participants who were serologically susceptible to IMD caused by Nmen strain 5/99 at baseline, but who became serologically protected by IMD caused by this strain after becoming colonised by 4NB1 [34]. Here, we expand upon this earlier work and plan to determine whether inoculation with 4xrNlac, a multi-strain GM-Nlac inoculum comprising four stains expressing NadA and one of 4 FHbp variants representative of UK-circulating meningococcal strains, results in safe colonisation and enhances anti-meningococcal immune responses. If safe colonisation is confirmed, a subsequent randomised, placebo-controlled study will be performed within the same funding envelope where we will establish whether colonisation with 4xrNlac induces sterilising protection against homologous rechallenge, and outline associated immune correlates of protection, including generation of resident memory adaptive immune responses. If positive, such a study would pave the way for future investigation to determine whether 4xrNlac might induce heterologous protection against Nmen.

## Supporting information

Clinical Study Protocol - GMNlac Pilot, V1.2

## Data Availability

There are no experimental data outlined in this manuscript that are not already freely available, noting that this manuscript is a clinical study protocol only.

## Authors’ Contributions

The clinical study was designed by DFG, APD and JRL with contributions from RCR, ARG, PJS and SNF. Pre-clinical work and microbiological and immunological assay development relevant to this study protocol were performed by ARH, TC, RA, RSE, DM, DWC, JL, RB, APD, DFG and JRL. The first drafts of the manuscript were prepared by GC, PJS, DFG, APD and JRL. All authors contributed to editing and approval of the final version of the manuscript.

## References

1 Stephens DS, Greenwood B, Brandtzaeg P. Epidemic meningitis, meningococcaemia, and Neisseria meningitidis. Lancet. 2007;369:2196–210. doi: 10.1016/S0140-6736(07)61016-2

2 Marshall HS, McMillan M, Koehler AP, et al. Meningococcal B Vaccine and Meningococcal Carriage in Adolescents in Australia. N Engl J Med. 2020;382:318–27. doi: 10.1056/NEJMoa1900236

3 Arnold FW, Parrish LW, Marimuthu S, et al. Meningococcal carriage and transmission dynamics in college students in Louisville, Kentucky. PLoS One. 2026;21:e0344194. doi: 10.1371/journal.pone.0344194

4 Harvey-Cowlishaw T, Wartnaby S, Trujillo-Mendoza J, et al. Meningococcal carriage in UK university students post-pandemic: evidence for MenACWY vaccine impact and ongoing genogroup B risk. Front Public Health. 2026;14:1917586. doi: 10.3389/fpubh.2026.1917586

5 Weil LM, Crowe SJ, Rubis AB, et al. Risk Factors for Serogroup B Meningococcal Disease Among College Students. Open Forum Infect Dis. 2023;10:ofad607. doi: 10.1093/ofid/ofad607

6 I’Anson J, Anderson C, Arora S, et al. Large outbreak of group B invasive meningococcal disease in young adults in South East England, March 2026. Euro Surveill. 2026;31:2600288. doi: 10.2807/1560-7917.ES.2026.31.15.2600288

7 Carr JP, MacLennan JM, Plested E, et al. Impact of meningococcal ACWY conjugate vaccines on pharyngeal carriage in adolescents: evidence for herd protection from the UK MenACWY programme. Clin Microbiol Infect. 2022;28:1649.e1–1649.e8. doi: 10.1016/j.cmi.2022.07.004

8 Diaz Romero J, Outschoorn IM. Current status of meningococcal group B vaccine candidates: capsular or noncapsular? Clin Microbiol Rev. 1994;7:559–75. doi: 10.1128/CMR.7.4.559

9 Read RC, Baxter D, Chadwick DR, et al. Effect of a quadrivalent meningococcal ACWY glycoconjugate or a serogroup B meningococcal vaccine on meningococcal carriage: an observer-blind, phase 3 randomised clinical trial. Lancet. 2014;384:2123–31. doi: 10.1016/S0140-6736(14)60842-4

10 McMillan M, Chandrakumar A, Wang HLR, et al. Effectiveness of Meningococcal Vaccines at Reducing Invasive Meningococcal Disease and Pharyngeal Neisseria meningitidis Carriage: A Systematic Review and Meta-analysis. Clin Infect Dis. 2021;73:e609–19. doi: 10.1093/cid/ciaa1733

11 Defeating Meningitis By 2030: A Global Road Map. 1st ed. Geneva: World Health Organization 2021.

12 Laver JR, Hughes SE, Read RC. Neisserial Molecular Adaptations to the Nasopharyngeal Niche. Adv Microb Physiol. 2015;66:323–55. doi: 10.1016/bs.ampbs.2015.05.001

13 Goldblatt D, Hussain M, Andrews N, et al. Antibody responses to nasopharyngeal carriage of Streptococcus pneumoniae in adults: a longitudinal household study. J Infect Dis. 2005;192:387–93. doi: 10.1086/431524

14 Hall DB, Lum MK, Knutson LR, et al. Pharyngeal carriage and acquisition of anticapsular antibody to Haemophilus influenzae type b in a high-risk population in southwestern Alaska. Am J Epidemiol. 1987;126:1190–7. doi: 10.1093/oxfordjournals.aje.a114758

15 Jones GR, Christodoulides M, Brooks JL, et al. Dynamics of carriage of Neisseria meningitidis in a group of military recruits: subtype stability and specificity of the immune response following colonization. J Infect Dis. 1998;178:451–9. doi: 10.1086/515622

16 Cartwright KA, Stuart JM, Jones DM, et al. The Stonehouse survey: nasopharyngeal carriage of meningococci and Neisseria lactamica. Epidemiol Infect. 1987;99:591–601. doi: 10.1017/s0950268800066449

17 Bennett JS, Griffiths DT, McCarthy ND, et al. Genetic diversity and carriage dynamics of Neisseria lactamica in infants. Infect Immun. 2005;73:2424–32. doi: 10.1128/IAI.73.4.2424-2432.2005

18 Simmons G, Martin D, Stewart J, et al. Carriage of N. lactamica in a population at high risk of meningococcal disease. Epidemiol Infect. 2000;125:99–104. doi: 10.1017/s095026889900415x

19 Olsen SF, Djurhuus B, Rasmussen K, et al. Pharyngeal carriage of Neisseria meningitidis and Neisseria lactamica in households with infants within areas with high and low incidences of meningococcal disease. Epidemiol Infect. 1991;106:445–57. doi: 10.1017/s0950268800067492

20 Deasy AM, Guccione E, Dale AP, et al. Nasal Inoculation of the Commensal Neisseria lactamica Inhibits Carriage of Neisseria meningitidis by Young Adults: A Controlled Human Infection Study. Clinical Infectious Diseases. 2015;60:1512–20. doi: 10.1093/cid/civ098

21 Coen PG, Cartwright K, Stuart J. Mathematical modelling of infection and disease due to Neisseria meningitidis and Neisseria lactamica. Int J Epidemiol. 2000;29:180–8. doi: 10.1093/ije/29.1.180

22 Evans CM, Pratt CB, Matheson M, et al. Nasopharyngeal colonization by Neisseria lactamica and induction of protective immunity against Neisseria meningitidis. Clin Infect Dis. 2011;52:70–7. doi: 10.1093/cid/ciq065

23 Dale AP, Theodosiou AA, Gbesemete DF, et al. Effect of colonisation with *Neisseria lactamica* on cross-reactive anti-meningococcal B-cell responses: a randomised, controlled, human infection trial. Lancet Microbe. 2022;3:e931–43. doi: 10.1016/S2666-5247(22)00283-X

24 Finney M, Vaughan T, Taylor S, et al. Characterization of the key antigenic components and pre-clinical immune responses to a meningococcal disease vaccine based on Neisseria lactamica outer membrane vesicles. Hum Vaccin. 2008;4:23–30. doi: 10.4161/hv.4.1.4806

25 Borkner L, Curham LM, Wilk MM, et al. IL-17 mediates protective immunity against nasal infection with Bordetella pertussis by mobilizing neutrophils, especially Siglec-F+ neutrophils. Mucosal Immunol. 2021;14:1183–202. doi: 10.1038/s41385-021-00407-5

26 Solans L, Debrie A-S, Borkner L, et al. IL-17-dependent SIgA-mediated protection against nasal Bordetella pertussis infection by live attenuated BPZE1 vaccine. Mucosal Immunol. 2018;11:1753–62. doi: 10.1038/s41385-018-0073-9

27 Hill AR, Gbesemete DF, Coutinho T, et al. BPZE1 vaccination induces IL-17+ and IL-22+ CD4+ T cells associated with nasal mucosal secretory IgA responses in humans: effects on virulent Bordetella pertussis colonisation. npj Vaccines. 2026;11:121. doi: 10.1038/s41541-026-01443-7

28 Moffitt KL, Gierahn TM, Lu Y, et al. TH17-Based Vaccine Design for Prevention of Streptococcus pneumoniae Colonization. Cell Host & Microbe. 2011;9:158–65. doi: 10.1016/j.chom.2011.01.007

29 O’Connor W, Zenewicz LA, Flavell RA. The dual nature of TH17 cells: shifting the focus to function. Nat Immunol. 2010;11:471–6. doi: 10.1038/ni.1882

30 Dorey RB, Theodosiou AA, Read RC, et al. The nonpathogenic commensal Neisseria: friends and foes in infectious disease. Curr Opin Infect Dis. 2019;32:490–6. doi: 10.1097/QCO.0000000000000585

31 Pandey A, Cleary DW, Laver JR, et al. Microevolution of Neisseria lactamica during nasopharyngeal colonisation induced by controlled human infection. Nat Commun. 2018;9:4753. doi: 10.1038/s41467-018-07235-5

32 Theodosiou AA, Bogaert D, Cleary DW, et al. Controlled human infection model of Neisseria lactamica in late pregnancy investigating mother-to-infant transmission in the UK: a single-arm pilot trial. Lancet Microbe. 2025;6:100986. doi: 10.1016/j.lanmic.2024.100986

33 Gbesemete DF, Haidara F, Laver JR, et al. Controlled Human Infection of Healthy Adults With Lyophilized Neisseria lactamica Induces Asymptomatic, Immunogenic Nasopharyngeal Carriage in the United Kingdom and Mali. Open Forum Infect Dis. 2026;13:ofaf809. doi: 10.1093/ofid/ofaf809

34 Laver JR, Gbesemete D, Dale AP, et al. A recombinant commensal bacteria elicits heterologous antigen-specific immune responses during pharyngeal carriage. Sci Transl Med. 2021;13:eabe8573. doi: 10.1126/scitranslmed.abe8573

35 Metruccio MME, Pigozzi E, Roncarati D, et al. A novel phase variation mechanism in the meningococcus driven by a ligand-responsive repressor and differential spacing of distal promoter elements. PLoS Pathog. 2009;5:e1000710. doi: 10.1371/journal.ppat.1000710

36 Comanducci M, Bambini S, Brunelli B, et al. NadA, a novel vaccine candidate of Neisseria meningitidis. J Exp Med. 2002;195:1445–54. doi: 10.1084/jem.20020407

37 Capecchi B, Adu-Bobie J, Di Marcello F, et al. Neisseria meningitidis NadA is a new invasin which promotes bacterial adhesion to and penetration into human epithelial cells. Mol Microbiol. 2005;55:687–98. doi: 10.1111/j.1365-2958.2004.04423.x

38 Clark SA, Willerton L, Claus H, et al. Exploring the sequence diversity and surface expression of Factor H-Binding Protein among invasive serogroup B meningococcal strains from selected European countries. Hum Vaccin Immunother. 2024;20:2427471. doi: 10.1080/21645515.2024.2427471

39 Madico G, Welsch JA, Lewis LA, et al. The meningococcal vaccine candidate GNA1870 binds the complement regulatory protein factor H and enhances serum resistance. J Immunol. 2006;177:501–10. doi: 10.4049/jimmunol.177.1.501

40 Masignani V, Comanducci M, Giuliani MM, et al. Vaccination against Neisseria meningitidis using three variants of the lipoprotein GNA1870. J Exp Med. 2003;197:789–99. doi: 10.1084/jem.20021911

41 Pajon R, Beernink PT, Granoff DM. Design of meningococcal factor H binding protein mutant vaccines that do not bind human complement factor H. Infect Immun. 2012;80:2667–77. doi: 10.1128/IAI.00103-12

42 van der Veen S, Johnson S, Jongerius I, et al. Nonfunctional variant 3 factor H binding proteins as meningococcal vaccine candidates. Infect Immun. 2014;82:1157–63. doi: 10.1128/IAI.01183-13

43 Granoff DM, Giuntini S, Gowans FA, et al. Enhanced protective antibody to a mutant meningococcal factor H-binding protein with low-factor H binding. JCI Insight. 2016;1:e88907. doi: 10.1172/jci.insight.88907

44 Konar M, Rossi R, Walter H, et al. A Mutant Library Approach to Identify Improved Meningococcal Factor H Binding Protein Vaccine Antigens. PLoS One. 2015;10:e0128185. doi: 10.1371/journal.pone.0128185

45 Konar M, Pajon R, Beernink PT. A meningococcal vaccine antigen engineered to increase thermal stability and stabilize protective epitopes. Proc Natl Acad Sci U S A. 2015;112:14823–8. doi: 10.1073/pnas.1507829112

46 Dale AP, Gbesemete DF, Read RC, et al. Neisseria lactamica Controlled Human Infection Model. Methods Mol Biol. 2022;2414:387–404. doi: 10.1007/978-1-0716-1900-1_21

47 Gbesemete D, Ramasamy MN, Ibrahim M, et al. Efficacy, immunogenicity, and safety of the live attenuated nasal pertussis vaccine, BPZE1, in the UK: a randomised, placebo-controlled, phase 2b trial using a controlled human infection model with virulent *Bordetella pertussis*. The Lancet Microbe. 2025;101211. doi: 10.1016/j.lanmic.2025.101211

48 de Graaf H, Gbesemete DF, Hill AR, et al. Safety, colonisation kinetics, transmissibility, and immune correlates of protection in healthy adults inoculated with Bordetella pertussis in England: a single-centre, open-label, phase 1, controlled human infection study. Lancet Microbe. 2026;7:101313. doi: 10.1016/j.lanmic.2025.101313

