## Supplementary tables for "Protocol for a controlled human infection with genetically modified *Neisseria lactamica* expressing the meningococcal vaccine antigens NadA and FHbp: *The GM-Nlac Pilot Study*"

| **Inclusion criteria** | **Challenge participant** | **Contact participant** |
| --- | --- | --- |
| Healthy adults aged 18 to 45 years inclusive on the day of enrolment | 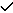 |  |
| Healthy adults aged 18 to 55 years inclusive on the day of enrolment |  | 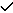 |
| Fully conversant in the English language | 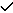 | 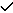 |
| Able and willing (in the investigator’s opinion) to comply with all study requirements | 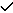 | 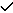 |
| Able to correctly answer all questions in the pre-consent questionnaire | 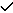 | 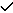 |
| Provide written informed consent to participate in the study including agreement to abide by infection control guidelines during the study period | 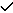 | 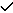 |
| Provide written consent to allow the study team to discuss the participant’s medical history with the General Practitioner and access electronic medical records | 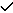 | 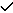 |
| Written informed consent provided by any bedroom sharer who is eligible to be enrolled as a contact participant (if applicable) | 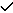 |  |
| For females of child-bearing potential, willingness to practice continuous effective contraception during the study and a negative pregnancy test on the days of screening (both) and challenge (challenge participants only) | 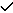 | 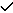 |
| Use of continuous effective contraception during the study | 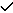 | 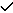 |
| Agreement to take antibiotic eradication therapy according to the study protocol | 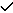 | 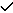 |

**Supplementary Table 1 – Inclusion criteria**

| Exclusion criteria | Challenge  participant | Contact participant |
| --- | --- | --- |
| Individuals who have a current infection at the time of inoculation | 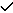 |  |
| Use of systemic antibiotics within the period 30 days prior to inoculation | 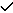 |  |
| Current active smokers defined as having smoked or vaped in the week prior to inoculation | 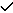 |  |
| *N. lactamica* or *N. meningitidis* detected on throat swab or nasal wash taken at screening or at the pre-challenge visit | 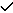 |  |
| Individuals who have been challenged with wild-type or GM-Nlac in a previous controlled human infection study | 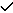 |  |
| Individuals who have received one or more doses of a meningococcus B vaccine (Bexsero or Trumenba) | 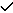 |  |
| History of allergy or intolerance to any component of the inoculum | 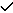 |  |
| Contraindications to the use of ciprofloxacin, specifically:   - Hypersensitivity to quinolones - History of tendon disorders related to quinolone use - Epilepsy - Personal or family history of aneurysm or congenital heart valve disease - Prolonged QT interval | 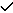 | 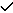 |
| Contraindications to the use of ceftriaxone, specifically:   - Known hypersensitivity to cephalosporins - Severe penicillin allergies (e.g. anaphylaxis or Stevens Johnson Syndrome) | 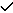 | 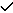 |
| Any confirmed or suspected immunosuppressive or immune-deficient state, specifically terminal complement component deficiencies, eculizumab use, known HIV infection, malignancy, asplenia, recurrent severe infections and chronic (more than 14 days) immunosuppressant medication within the past 6 months (topical steroids are allowed) | 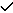 | 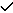 |
| Use of immunoglobulins or blood products within 3 months prior to enrolment | 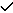 | 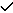 |
| Household, close social or regular occupational contact with persons known to be immunosuppressed, specifically HIV infection with a CD4 count <200 cells/mm3; asplenia; any malignancy, recurrent, severe infections and chronic (more than 14 days) immunosuppressant medication within the past 6 months (topical steroids are allowed) | 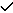 | 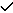 |
| Household or regular occupational contact with children under 5 years or an older child with a tendency to co-sleep with the participant | 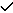 | 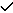 |
| Any abnormal finding on clinical examination or screening investigations assessed by the investigator to be clinically significant. In the event of abnormal test results, confirmatory repeat tests may be requested. | 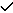 | 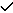 |
| Pregnancy, lactation or intention to become pregnant during the study | 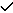 | 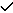 |
| Any other significant disease, disorder, or finding which may significantly increase the risk to the participant, affect their participation in the study or impair interpretation of the study data, for example recent surgery to the nasopharynx | 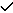 | 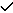 |

**Supplementary Table 2 – Exclusion criteria**

| **Visit** | | **Scr** | **PC** | **C0** | **C1^a^** | **C4** | **C7** | | **C14** | **C28** | **PE^b^** |
| --- | --- | --- | --- | --- | --- | --- | --- | --- | --- | --- | --- |
| Window (Days) | | C0 -5-90 | C0 -5-10 | 0 | C0 +1 | C0 +3-5 | C0 +6-8 | | C0 +12-16 | C0 +25-31 | 1-4 post abx |
| Visit Type | | Screening | Pre-challenge | Challenge | Surveillance | | | | | Eradication | Post eradication |
|  |  |  |  |  | Daily diary card | | |  | |  |  |
|  | TOPS | x |  |  |  |  |  | |  |  |  |
|  | PCQ | x |  |  |  |  |  | |  |  |  |
| **Challenge participant** | Informed consent | x |  |  |  |  |  | |  |  |  |
|  | Request GP records  / electronic medical records | x |  |  |  |  |  | |  |  |  |
|  | Medical history | x |  |  |  |  |  | |  |  |  |
|  | Physical examination | x |  |  |  |  |  | |  |  |  |
|  | Height and weight | x |  |  |  |  |  | |  |  |  |
|  | Reaffirm consent |  | x | x | x | x | x | | x | x | x |
|  | Reconfirm eligibility |  | x | x |  |  |  | |  |  |  |
|  | Review of adverse events & concomitant medications |  | x | x | x^c^ | x^c^ | x^c^ | | x | x | x |
|  | Targeted history and examination |  | (x) | (x) |  | (x) | (x) | | (x) | (x) | (x) |
|  | Vital Signs | x | x | x |  | x | x | | x | x | x |
|  | Urinalysis | x |  |  |  |  |  | |  |  |  |
|  | Urine toxicology | x |  |  |  |  |  | |  |  |  |
|  | Pregnancy Test *(females only)* | x |  | x |  |  |  | |  | x |  |
|  | ECG | x |  |  |  |  |  | |  |  |  |
|  | Throat swab x 1-2 | x | x |  |  | x | x | | x | x | x |
|  | Nasal wash |  |  |  |  | x | x | | x | x | x |
|  | Nasosorption sample |  | x |  |  | x | x | | x | x |  |
|  | Inferior turbinate swab |  | x |  |  | x | x | | x | x |  |
|  | Saliva sample |  | x |  |  | x | x | | x | x |  |
|  | Safety bloods | 8 |  | 8 |  | 8 | 8 | | 8 | 8 |  |
|  | Research bloods (maximum ml) |  |  | 60 |  |  |  | | 50 | 60 |  |
|  | Maximum total blood | 8 |  | 76 |  | 84 | 92 | | 150 | 218 |  |
|  | Inoculation |  |  | x |  |  |  | |  |  |  |
|  | Eradication (Ciprofloxacin 500 mg) |  |  |  |  |  |  | |  | x |  |
|  | CRF/ eCRF completion | x | x | x |  | x | x | | x | x | x |

Supplementary Table 3: Schedule of procedures – Challenge participants (x) if clinically indicted / optional, ^a^Telephone contact, ^b^PE visit planned for Day 30 but if early antibiotics are required then it will occur 1-4 days following antibiotic administration, ^c^Including diary card review

| **Visit** | | **Scr** |  | **C28** | | **PE^a^** |
| --- | --- | --- | --- | --- | --- | --- |
| Window (Days) | | C0 - 2-90 |  | C0 + 25-31 | | 1-4 post abx |
| **Contact participant** | PCQ, TOPS, Informed consent | x | 24-hour telephone number for study team, additional visit with throat swab and other investigations as clinically indicated if symptoms suggestive of respiratory or systemic infection, early antibiotics if found to be colonised | |  |  |
|  | Reaffirm consent |  |  |  | x | (x) |
|  | Medical history and examination including Ht and Wt | x |  |  |  |  |
|  | Review of adverse events & concomitant medications |  |  |  | x | (x) |
|  | Targeted history and examination |  |  |  | (x) | (x) |
|  | Vital Signs | x |  |  | x | (x) |
|  | Pregnancy Test *(females only)* | x |  |  | x |  |
|  | ECG | x |  |  |  |  |
|  | Throat swab x 1-2 | x |  |  | x | (x) |
|  | Eradication (Ciprofloxacin 500 mg) TTO^b^ |  |  |  | x |  |
|  | CRF/ eCRF completion | x |  |  | x | (x) |

Supplementary Table 4 : Schedule of procedures – Contact participants

(x) if clinically indicted / optional, ^a^Contact participants will only attend PE visit if antibiotics are indicated. ^b^Antibiotic eradication therapy will be supplied as a TTO at the C28 visit but contact participants will only be instructed to take them if their C28 throat swab is positive
