## Supplementary material for "Protocol for a controlled human infection with genetically modified *Neisseria lactamica* expressing the meningococcal vaccine antigens NadA and FHbp: *The GM-Nlac Pilot Study*": Clinical Study Protocol - GMNlac Pilot, V1.2

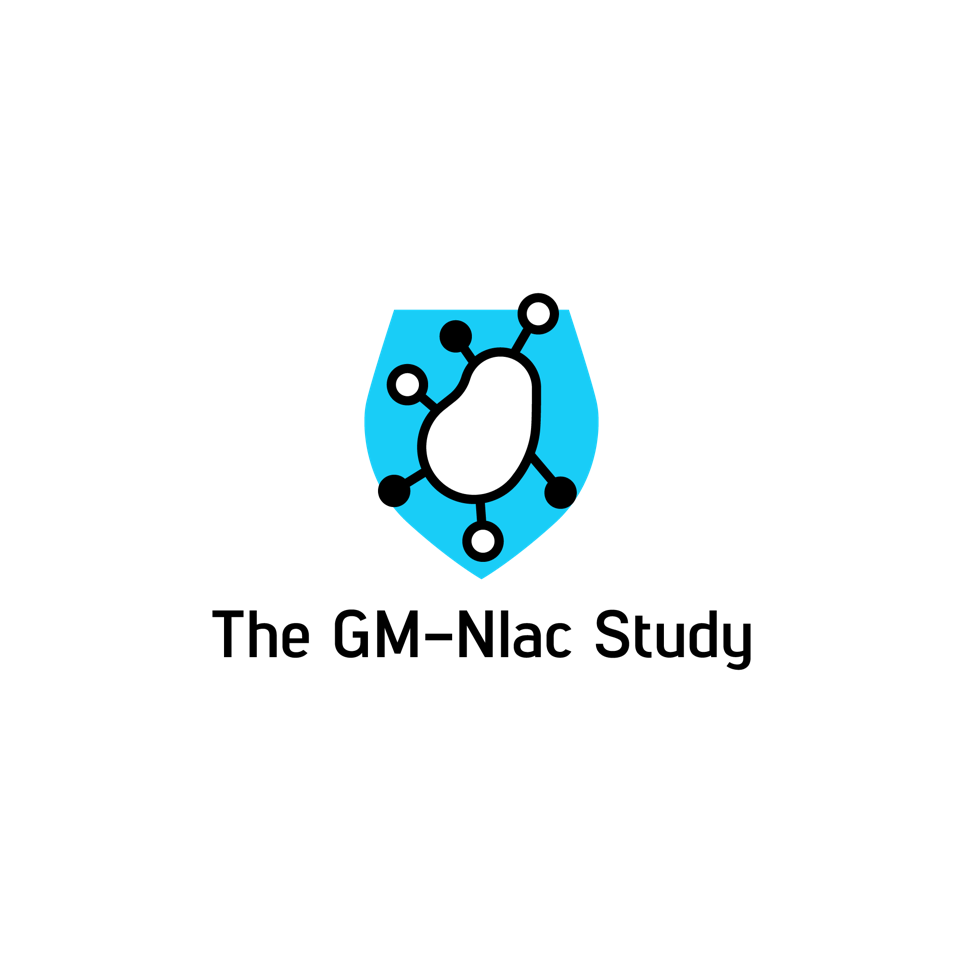

Pilot

**Safety, colonisation and immunogenicity following nasal inoculation with genetically modified *Neisseria lactamica* expressing Factor H binding protein and Neisseria adhesin A – a pilot controlled human infection study**

**Sponsor Study reference: ERGO 81849**

**NHS REC Study Reference: 26/SC/0072**

**Protocol version: 1.2**

**Date: 29/03/2026**

**Chief Investigator: Dr Adam Dale**

**Lead Scientific Investigator: Dr Jay Laver**

**Principal Investigator: Dr Diane Gbesemete**

**University of Southampton**

**University Hospital Southampton NHS Foundation Trust**

| **Version** | **Date** | **Authors** | **Modifications** |
| --- | --- | --- | --- |
| 1.0 | 04/12/25 | Diane Gbesemete  Jay Laver  Adam Dale |  |
| 1.1 | 06/02/26 | Diane Gbesemete  Jay Laver  Adam Dale | Response to sponsor comments:   - Additional infection control rule regarding dental treatment (as per ACRE) - Updated recruitment strategy - Clarified reporting timelines for SUSARs - Updated REC SOP version |
| 1.2 | 29/03/26 | Diane Gbesemete  Jay Laver  Adam Dale | Response to REC / HRA review   - Clarification of requirements for contraception use - Update study start date - Minor formatting corrections |

**The GM-Nlac study – Pilot**

**Safety, colonisation and immunogenicity following nasal inoculation with genetically modified *Neisseria lactamica* expressing Factor H binding protein and Neisseria adhesin A – a pilot controlled human infection study**

**Study references:**	ERGO 81849

RHM MED 2124

**Study steering committee:**

**Chief Investigator:	Dr Adam P. Dale**

Associate Professor

University of Southampton

C Level South Lab & Pathology Block

University Hospital Southampton

Tremona Road, Southampton, SO16 6YD

**Lead Scientific Investigator:	Dr Jay R. Laver**

Principal Research Fellow

University of Southampton

C Level South Lab & Pathology Block

University Hospital Southampton

Tremona Road, Southampton, SO16 6YD

**Principal Investigator:	Dr Diane F. Gbesemete**

Specialist in Clinical Research

University of Southampton

C Level South Lab & Pathology Block

University Hospital Southampton

Tremona Road, Southampton, SO16 6YD

**Additional study steering**

**committee members:	Professor Robert C. Read**

University of Southampton

C Level

South Lab & Pathology Block

Southampton

University Hospital Southampton

Tremona Road, Southampton, SO16 6YD

**Professor Andrew Gorringe**

UK Health Security Agency

Porton Down

Salisbury

SP4 0JG

**Co-investigators:	Professor Saul N. Faust**

NIHR Clinical Research Facility

University of Southampton

C Level, West Wing, Mailpoint 218

University Hospital Southampton

Tremona Road, Southampton, SO16 6YD

**Professor Ray Borrow**

Consultant Clinical Scientist

Meningococcal Reference Unit/Vaccine Evaluation Unit

UK Health Security Agency

Manchester Medical Microbiology Partnership

Clinical Sciences Building II

Manchester Royal Infirmary

Oxford Road

Manchester M13 9WL

**Dr Jay Lucidarme**

Senior Scientist

Meningococcal Reference Unit

UK Health Security Agency

Manchester Medical Microbiology Partnership

Second floor, Clinical Sciences Building II

Manchester Royal Infirmary

Oxford Road

Manchester M13 9WL

**Professor James Wing**

Human Single Cell Immunology team

Center for Infectious Disease Education and

Research (CiDER)

Immunology Frontier Research Center (IFReC)

Osaka University

**Statistician:	Dr Helen Moyses**

NIHR Southampton Biomedical Research Centre
University Hospital Southampton NHS Foundation Trust
Mailpoint 218
Southampton General Hospital
Tremona Road, Southampton, SO16 6YD

**Study Site:	NIHR-CRF**

University Hospital Southampton NHS Foundation Trust

C Level, West Wing, Mailpoint 218

University Hospital Southampton NHS Foundation Trust

Tremona Road, Southampton, SO16 6YD

**Sponsor	The University of Southampton**
University Road

Southampton

SO17 1BJ

**Funded by**	Medical Research Council Grant MR/X019284/1

**Confidentiality Statement**

This document contains confidential information that must not be disclosed to anyone other than the sponsor, the investigator team, and members of the independent ethics committee. This information cannot be used for any purpose other than the evaluation or conduct of the clinical investigation without the prior written consent of Dr. Adam Dale.

**Investigator Agreement**

I have read this protocol and agree to abide by all provisions set forth therein.

I agree to comply with the principles of the International Conference on Harmonisation Tripartite Guideline on Good Clinical Practice.

According to the Declaration of Helsinki, 2008, I declare no conflict of interest.

Dr. Adam P. Dale	………………………	…………………

Chief Investigator	Signature	Date

Dr. Jay R. Laver	…………………………	………………….

Lead Scientific Investigator	Signature	Date

Dr. Diane F. Gbesemete	…………………………	………………….

Principal Investigator	Signature	Date

### 1 Synopsis

| **Title** | **The GM-Nlac study - Pilot**  **Safety, colonisation and immunogenicity following nasal inoculation with genetically modified *Neisseria lactamica* expressing Factor H binding protein and Neisseria adhesin A – a pilot controlled human infection study** |
| --- | --- |
| **Sponsor** | University of Southampton |
| **Study centre** | NIHR Clinical Research Facility, Southampton University Hospital NHS Foundation Trust, Southampton, SO16 6YD |
| **Sponsor Reference** | ERGO 81849 |
| **Study design** | Single arm controlled human infection model experiment |
| **Population** | Challenge participants: Healthy adult volunteers aged 18-45 years  Contact participants: Bedroom sharers of challenge participants – healthy adult volunteers aged 18-55 years |
| **Sample size** | Challenge participants: 10  Contact participants: maximum of 1 per challenge participant, estimated 5) |
| **Follow up duration** | 35 days from pre-challenge visit |
| **Microbial challenge material** | 4xrNlac –10^5^ of each of 4 strains of genetically modified *N. lactamica* expressing FHbP and NadA in phosphate buffered saline  Total volume 1ml |
| **Planned Study period** | 1^st^ May 2026 – 30^th^ April 2036 |

### 3 Abbreviations

| AE | Adverse Event |
| --- | --- |
| ALP | Alkaline phosphatase |
| ALT | Alanine transaminase |
| AR | Adverse Reaction |
| B_PLAS_ | Plasma B cells |
| CFU | Colony Forming Unit |
| CI | Chief Investigator |
| CRF | Case Report Form |
| DEFRA | Department for Environment, Food and Rural Affairs |
| DNA | Deoxyribonucleic acid |
| DSMB | Data safety and monitoring board |
| ECG | Electrocardiogram |
| FHbp | Factor H binding protein |
| GCP | Good Clinical Practice |
| GDPR | General Data Protection Regulation |
| GM | Genetically modified |
| GM-Nlac | Genetically modified *Neisseria lactamica* |
| GMO | Genetically modified organism |
| GP | General Practitioner |
| hFH | Human Factor H |
| HRA | Health Research Authority |
| ICH | International Conference on Harmonisation |
| IMD | Invasive meningococcal disease |
| MHRA | Medicine and Healthcare products Regulatory Agency |
| NadA | Neisserial Adhesin A |
| NHS | National Health Service |
| NIHR | National Institute for Health and Care Research |
| NIHR-CRF | NIHR Clinical Research Facility |
| Nlac | *Neisseria lactamica* |
| Nmen | *Neisseria meningitidis* |
| PBS | Phosphate Buffered Saline |
| PCR | Polymerase Chain Reaction |
| PI | Principal Investigator |
| PPE | Personal Protective Equipment |
| QA | Quality assurance |
| QC | Quality control |
| QTc | Corrected QT interval |
| qPCR | Quantitative Polymerase Chain Reaction |
| REC | Research Ethics Committee |
| RNA | Ribonucleic acid |
| SAE | Serious Adverse Event |
| SAR | Serious Adverse Reaction |
| SOP | Standard Operating Procedure |
| SUSAR | Serious Unexpected Serious Adverse reaction |
| TCR | T-cell receptor |
| TOPS | The Over-volunteering Prevention System |
| UAR | Unexpected Adverse Reaction |
| UHS | University Hospital Southampton |
| UK | United Kingdom |
| UoS | University of Southampton |
| X-gal | 5-bromo-4-chloro-3-indolyl β-D-galactopyranoside |

### 4 Background and rationale

The purpose of the study described in this protocol is to establish the safety, induction of nasopharyngeal colonisation, and subsequent immunogenicity following intranasal inoculation of healthy human adults with an inoculum containing four genetically modified (GM) strains of *Neisseria lactamica* (hereafter, GM-Nlac). These data will be used to inform the design of a subsequent randomised, double-blind placebo controlled human infection study to establish the impact of colonisation with GM-Nlac on protection at homologous rechallenge.

In addition to a full, wild type complement of endogenous *Neisseria lactamica* proteins, each of the four GM-Nlac strains also express two meningococcal antigens: *Neisseria* Adhesin A (NadA, a type V meningococcal autotransporter), and one of four different variants of functionally-attenuated Factor H binding protein (FHbp, a meningococcal outer membrane protein).

**As the study involves the deliberate release of four genetically modified bacteria into the community it has been considered and approved by the responsible government ministry (DEFRA – ref 25/R50/01).**

#### 4.1 *Neisseria meningitidis* and *Neisseria lactamica*

*Neisseria meningitidis* (Nmen) and *Neisseria lactamica* (Nlac) are closely related bacteria which exclusively colonise the human nasopharynx (1).

Nmen is a pathobiont; the vast majority of Nmen colonisation episodes are non-invasive and asymptomatic, but in a minority of cases Nmen invades and rapidly proliferates, resulting in meningococcal disease. This is characterised by an acute onset and rapid progression, typically in previously healthy children and young people, often resulting in death or devastating sequelae (2). Cases occur sporadically, in outbreaks and in larger epidemics, particularly within the meningitis belt of sub-Saharan Africa, with the potential to overwhelm health systems (3). While effective vaccines have substantially reduced the burden of disease in many populations, meningococcal disease remains a globally important health concern (4). In 2021 the WHO published a roadmap aiming to defeat meningococcal disease by 2030, recognising the ongoing need for effective strategies for prevention and control of disease (5).

Nlac is commensal organism which does not cause disease in immunocompetent hosts. Case reports of association with symptomatic infection or disease are extremely rare, and almost exclusively occur in the context of significant immunocompromise (6-17).

##### 4.1.1 Microbiological features

Nlac and Nmen are both Gram-negative aerobic, oxidase positive diplococci. Nlac is acapsulate and is the only member of the genus to ferment lactose due to the action of β-D-galactosidase, which is encoded by the gene *lacZ*. This allows Nlac to be differentiated from other *Neisseria* by the appearance of colonies on media containing the chromogenic substrate 5-bromo-4-chloro-3-indolyl β-D-galactopyranoside (X-gal). Colonies of Nlac appear blue whereas colonies of other *Neisseria* species, including Nmen, will appear white or off-white (18). Nmen strains are often capsulate, with classification into serogroups on the basis of capsular structure. The expression of capsule is the key virulence factor allowing evasion of the host immune response (19).

##### 4.1.2 Colonisation with *Neisseria* species

The highest rate of natural Nlac carriage occurs in infants at approximately 40% by one year (20). Carriage rates wane in toddlers and older children and by adulthood, it is about 1% (21). This differs to the pattern of Nmen carriage. In the UK, at any given time, an average of 10% of the human population are carrying this organism in the nose and throat (detectable by throat swabbing) but this varies with age. The highest rates of Nmen carriage are seen in teenagers and University students. Activities associated with Nmen carriage include attendance at pubs or clubs, smoking, residence in student halls of residence and saliva exchange, for example kissing (22, 23). Infants have a low carriage rate, but this increases gradually as childhood progresses. Although Nmen carriage is quite common, disease is extremely rare – currently less than 1 per 100,000 per annum in the UK. Therefore, the likelihood of disease, even when a person is carrying a virulent strain of Nmen, is very low (24).

##### 4.1.3 Epidemiological relationship between *N. meningitidis* and *N. lactamica*

There is an epidemiological relationship between carriage of Nlac and Nmen. As described above, observed age-specific rates of Nmen carriage are inversely proportional to those of Nlac (21). The mechanism underpinning this relationship is undetermined, but is unlikely due to the induction of cross-reactive serum bactericidal activity (SBA) by Nlac, as the early years of life associated with high rates of Nlac carriage predate the development of natural meningococcal SBA (24). Colonisation with Nlac in adults does result in generation of cross-reactive humoral responses and memory B cell responses, including development of cross-reactive anti-meningococcal opsonophagocytic activity (25, 26). However, the role these responses may play in protection against Nmen colonisation and disease remains undefined. Co-carriage of both species is rare (27, 28). A negative association between Nlac carriage and meningococcal disease was demonstrated during an outbreak of meningococcal disease in the Faroe Islands. Comparison was made between distinct areas of high and low incidence of meningococcal disease. Areas of high disease incidence had significantly lower Nlac carriage rates than areas of low disease incidence (29).

These observations have led to the hypothesis that Nlac carriage may provide protection from both Nmen carriage and meningococcal disease. Mathematical modelling has estimated a potential duration of protection of 4-5 years (30).

##### 4.1.4 Vaccine-induced protection from *N. meningitidis* colonisation and disease

As colonisation is a pre-requisite to both invasive infection and onward transmission, an ideal vaccine should induce protection against asymptomatic colonisation as well as invasive disease. Glycoconjugate meningococcal vaccines have been shown to induce such protection, and the reduction in disease seen following vaccination is partly due to herd protection conferred by reduced carriage and transmission (31, 32). Glycoconjugate vaccines targeting serogroups A, C, W-135 and Y have been available for several years, and more recently a pentavalent vaccine including serogroup X has been developed for use in the African meningitis belt (33). In contrast, vaccines marketed for protection against serogroup B disease, such as 4CMenB (Bexsero) and MenB-FHbp (Trumenba), target subcapsular antigens which induce bactericidal antibodies against a range of strains, but have not been shown to induce protection from asymptomatic colonisation (34-36).

Future prevention strategies will aim to target colonisation, using antigens known to induce immunity critical for colonisation, in age groups most likely to transmit to others. A direct and effective, but safe, way to assess colonisation with a pathogen is using a controlled human infection model known to induce colonisation with a defined organism expressing specific antigens of interest.

##### 4.1.5 Transmission of *Neisseria* species

Nmen is known to transmit to close contacts of those colonised with the organism, particularly those who share households. In a cardinal study undertaken in the UK between 1986 and 1987, 55 cases of meningococcal disease were identified in the South-West of England, an attack rate of 1.54 per 100,000 during the study period. The Nmen carriage rate in 384 close contacts was 18.2% for any Nmen strain, and 11.1% for the case strain. The carriage rate of indistinguishable strains in household contacts (16.0%) was higher than the carriage rate in contacts living at other addresses (7.0%, *p* < 0.05) (37). In a Norwegian study of the contacts of index cases of meningococcal disease, there was a high rate of carriage of the case strain of Nmen in patients' household members and kissing contacts, and this supports the practice of giving chemoprophylaxis to these contacts. The prevalence of carriage among other contacts was 2-3 times that found in the general population (0.7%) (38). After an outbreak of meningococcal disease in Singapore caused by Nmen serogroup W, associated with the Hajj pilgrimage in 2001, 15% of returning vaccinated pilgrims carried a single serogroup W clone, and 55% of these were still carriers 6 months later. Transmission to 8% of their unvaccinated household contacts occurred within 2 weeks, but no late transmission took place (39).

There is much less information on transmission of Nlac, but it is likely to be similar to that of Nmen. Carriage of Nlac among household contacts of meningococcal disease cases was investigated during an epidemic in Auckland, New Zealand. The overall Nlac carriage rate was 10.5% (95% CI 7.4-13.5%) with a peak colonisation rate in 2-year-olds of 61.5% (95% CI 26.6-88.1%). Factors associated with a significant (*p* < 0.05) increase in the likelihood of colonisation included runny nose, the number of people per bedroom and youth. Genetic analysis of isolates revealed a striking correlation of strains within the same household but a high level of diversity between households, suggesting that household contact is an important factor in acquisition (40). However, other studies have suggested that such household transmission may occur chiefly from infants and children rather than from adults (20, 41).

##### 4.1.6 Antibiotic clearance of Nmen carriage

Both Rifampicin and Ciprofloxacin, as oral antibiotics, have been shown to be effective in eradicating colonisation of Nmen, and are outlined as suitable agents for use as post-exposure prophylaxis in guidance (42). In adults, ciprofloxacin 500mg administered as a single oral dose is standard prophylaxis. In a cardinal study, single dose oral ciprofloxacin was given to all personnel in a naval training establishment as part of the management of an outbreak of meningococcal meningitis. Two thousand one hundred personnel received the drug and Nmen was eradicated from the pharynx of 97% of 570 who were swabbed two to four days later. In a cohort of 277 personnel who were followed for up to nine weeks, pharyngeal colonisation was eliminated from 93% of 104 carriers. The overall prevalence of colonisation fell from 19% to less than 1.5% as a result of the use of ciprofloxacin. Few side effects were encountered, compliance was good, and meningococci resistant to the antibiotic were not found after therapy (43). Another study showed eradication of Nmen from 100% of 21 previously colonised individuals within 24 hours of ciprofloxacin treatment (44).

#### 4.2 The *Neisseria lactamica* controlled human infection model

##### 4.2.1 Purpose and previous studies

The Nlac controlled human infection model was developed to investigate and potentially exploit the presumed epidemiological relationship between Nlac and Nmen. The wild-type model has been developed using Nlac strain Y92-1009, a strain originally isolated during a school carriage study in Northern Ireland in 1992. There have been seven wild-type Nlac CHI studies conducted so far, including healthy UK adults, healthy Malian adults (a setting within the meningitis belt) and healthy UK pregnant women (25, 26, 41, 45, 46). In these studies, the inoculum has been prepared from either frozen bacterial stocks or by reconstitution of lyophilised Nlac (LyoNlac). These studies are summarised in Table 1 on p19. A dose of 10^5^ colony forming units (CFU) has been established as the standard inoculum dose in UK healthy adults. This has achieved a colonisation fraction of 0.71-1.0 by Day 14 post inoculation in naïve UK adults using both frozen and lyophilised Nlac stocks (25, 41, 46). Nasopharyngeal colonisation has been shown to persist for several months following inoculation in a high proportion of individuals (25, 26, 45).

| Study | Population | Storage of bacterial stocks | Number of participants inoculated | Number of inoculations | Dose (CFU) | Colonisation fraction |
| --- | --- | --- | --- | --- | --- | --- |
| Lactamica 1 (26) | Healthy UK adults | Frozen | 54 | 78 | 10^4^ | 0.61 |
|  |  |  | 6 (non-colonised at 10^4^ CFU) | 6 | 10^5^ | 0.50 |
| Lactamica 2 (45) | Healthy UK adults | Frozen | 292 | 419 | 10^4^ | 0.34 |
| Lactamica 3 (25) | Healthy UK adults | Frozen | 12 | 12 | 10^5^ | 0.50 (by Day 4) |
|  |  |  | 6 | 6 | 10^5^ | 0.83 |
| Lactamica 5 (46) | Healthy UK adults | Lyophilised | 10 | 10 | 10^4^ | 0.60 |
|  |  |  | 10 | 10 | 5x10^4^ | 0.60 |
|  |  |  | 10 | 10 | 10^5^ | 1.00 |
| Lactamica 7 (25) | Healthy UK adults | Frozen | 20 | 20 | 10^5^ | 0.85 |
| Lactamica Etape 1 (46) | Healthy Malian adults | Lyophilised | 5 | 5 | 10^5^ | 0.60 |
|  |  |  | 15 | 15 | 10^6^ | 0.60 |
|  |  |  | 20 | 20 | 10^7^ | 0.65 |
| Lactamica 9 (41) | Pregnant UK women | Lyophilised | 21 | 21 | 10^5^ | 0.71 |

Table 1: Wild-type *N. lactamica* Controlled Human Infection studies

##### 4.2.2 Safety

There have been no adverse events attributed to nasopharyngeal colonisation with Nlac or Nlac disease in these studies to date. One adverse event of special interest occurred in the ‘Lactamica 5’ study in which a participant received an inoculum which was reported to have more than 10 times the intended viability. This participant remained asymptomatic. No serious adverse events have occurred. In Lactamica 5 several participants reported transient reactogenicity symptoms most probably due to the use of sterile water in reconstituting the lyophilised inoculum. LyoNlac appropriately reconstituted in PBS was well tolerated in Lactamica Etape 1 and Lactamica 9 (25, 26, 41, 45, 46).

##### 4.2.3 Immunogenicity

Experimentally induced nasopharyngeal colonisation with wild-type Nlac has been repeatedly demonstrated to be immunogenic, inducing Nlac-specific, and cross-reactive Nmen-specific humoral immune responses. Colonised participants develop serum IgG responses, IgA- and IgG-secreting plasma cell responses, and IgG memory B cell responses specific to Nlac and cross-reactive with Nmen. While Nlac colonisation induces Nmen- cross-reactive opsonophagocytic activity, there is no induction of anti-meningococcal SBA following wild-type Nlac colonisation. Of note, anti-Nlac/Nmen adaptive immune responses are not observed amongst non-colonised participants (25, 26, 41, 46).

#### 4.3 Controlled Human Infection with GM-Nlac

##### 4.3.1 Controlled human infection with GM-Nlac expressing antigens of interest

Controlled human infection with wild-type Nlac has been shown to be safe and to induce long-standing immunogenic nasopharyngeal colonisation in a high proportion of participants. This model has been further developed as an experimental medicine tool to study the immunological response to specific antigens by using a transformed strain of Nlac expressing antigens of interest (section 3.3.3).

Additionally, such a strain could have utility as a proxy to investigate the immunological mechanisms of protection against colonisation with pathobionts which express specific antigens. If colonisation with GM-Nlac expressing meningococcal antigens were able to induce protection against colonisation at rechallenge with a homologous strain, then it would suggest that such a strain may also protect against colonisation with Nmen expressing those antigens.

##### 4.3.2 Meningococcal antigens of interest

The specific antigens of interest in this study are the meningococcal antigens *Neisseria* adhesin A (NadA) and Factor H binding protein (FHbp). Neither are expressed by wild-type Nlac.

NadA is an adhesin protein expressed in a small subset of meningococcal strains. In a recent survey of European invasive Nmen isolates, only 16 of 235 possessed the *nadA* gene (47), suggesting the gene product is dispensible for pathogenesis. NadA is one of the four strongly immunogenic protein components of the anti-serogroup B meningococcal disease vaccine, Bexsero (4CMenB) (47).

The NadA protein is expressed as a single polypeptide, with three structurally significant domains: (1) the globular head domain, responsible for molecular interaction(s) with the as-yet unidentified epithelial cell receptor(s), (2) the helical ‘stalk’ domain, which passes through the cytoplasmic membrane and is exposed to the extracellular milieu and (3) the membrane-associated ‘pore’ domain, which self-assembles into the outer membrane and provides an appropriately-sized channel through which the rest of the polypeptide can pass. All three domains are essential for function. NadA-expression in Nmen is associated with an increased level of adhesion to and uptake by human epithelial cell lines (48). In wild-type Nmen, expression of the *nadA* gene is phase variable, in which DNA replication errors alter the number of repeat sequences immediately upstream of the *nadA* promoter (5’-TAAA-3’), modulating the transcriptional activity of the gene (49). Changes in the level of gene activity are reflected as increases or decreases in the level of NadA protein expression. In a longitudinal study of nasopharyngeal meningococcal carriage, it was shown that NadA expression in serial Nmen isolates decreased over time, hypothesised to be a result of seroconversion against NadA and the development of an antibody-mediated selective pressure against NadA expression (50). This hypothesis is partially corroborated by the finding that immunisation with recombinant NadA, prior to attempted Nmen colonisation in a transgenic mouse model, leads to sterilising immunity, whereby strains expressing a cognate NadA antigen were unable to colonise the murine nasopharynx (51). In each of the 4xrNlac strains discussed herein, *nadA* expression is instead controlled by a hybrid, constitutively active promoter that drives expression of the gene to high levels.

FHbp is an approximately 27 kDa outer membrane-anchored lipoprotein originally discovered using reverse vaccinology (52) and later shown to be responsible for protection of the meningococcus against complement-mediated killing through its ability to bind the human negative complement regulator, Factor H (53) through molecular mimicry of host carbohydrates (54). Unlike NadA, FHbp expression is widespread among circulating strains of the meningococcus, although invasive disease isolates that lack FHbp expression have been characterised (55). The amino acid sequence of FHbp is highly variable, phylogenetically resolving into three variant groups (var1, var2 and var3) or two subfamilies (1 and 2/3), depending on the classification system (herein, we refer exclusively to the variant group system of classification). Antibodies directed against one subfamily are poorly cross-reactive with the other, and there is only limited cross-reactivity of antibodies targeting epitopes from variant groups 2 and 3. FHbp is present in both the 4CMenB (Bexsero) and Trumenba anti-serogroup B invasive meningococcal disease vaccines, generating multiple antibody clones which, whilst not necessarily bactericidal on their own, can work synergistically to trigger C1q-mediated killing of the meningococcus (56).

##### 4.3.3 Controlled human infection with GM-Nlac expressing NadA

A previous GM-Nlac controlled human infection study has been conducted at the University of Southampton: **Experimental challenge of the human nasopharynx with recombinant *Neisseria lactamica* expressing the meningococcal type V autotransporter protein *Neisseria* Adhesin A (NadA): Lactamica 4**, (DEFRA REF 17/R50/01, REC REF 18/SC/0133) (57).

In this study, participants were nasally inoculated with one of two strains of GM-*N. lactamica* at a dose of 10^5^ CFU. The intervention group (n=12) received a strain which expressed NadA (strain 4NB1), while the control group (n=14) received a strain which contained a gene expression cassette without a coding sequence (i.e. a wild type-equivalent, but genetically modified control) (strain 4YB2). Challenged participants were monitored as inpatients for the first 4.5 days following inoculation, and then discharged and followed up as outpatients until day 90 post challenge. Shedding was monitored by culture of air and mask samples from colonised participants, and transmission was monitored by throat swabbing bedroom sharers over 90 days. Challenged participants and bedroom sharers received a single dose of oral ciprofloxacin at day 90 post inoculation, with clearance confirmed with a further throat swab after 24-48 hours.

In this work we demonstrated that:

(i) Colonisation of participants with both strains was safe
(ii) The colonisation fraction was 0.92 (11/12) with the NadA-expressing strain 4NB1 and 0.79 (11/14) with the control strain 4YB2
(iii) Colonisation persisted to day 90 in the majority of colonised participants in both groups
(iv) There was no detectable shedding of either strain or transmission to bedroom-sharers at any point over the 90-day colonisation period
(v) Colonisation was cleared by 1-2 days post ciprofloxacin in all colonised participants
(vi) A subset of participants colonised by the NadA-expressing strain produced NadA-specific IgG-secreting plasma B cells (B_PLAS_) and experienced an at-least 2-fold increase in serological anti-NadA IgG titres
(vii) All participants colonised by the NadA-expressing commensal experienced an increase in the proportion of circulating NadA-specific IgG-memory B cells (B_MEM_)
(viii) A subset of participants colonised by the NadA-expressing commensal seroconverted to become protected against invasive meningococcal disease (IMD) caused by a NadA-overexpressing strain of *N. meningitidis*, 5/99, consistent with the universally acknowledged correlate of protection against IMD (i.e. a strain-specific, serum bactericidal antibody (SBA) reciprocal titre greater than or equal to 4)

##### 4.3.4 4xrNlac

In this current study, participants will be inoculated with 4xrNlac, which is a multi-strain challenge agent, comprised of four different GM-Nlac strains, each at a target dose of 10^5^ CFU. Each strain expresses NadA and one of four different variants of FHbp, as summarised in Table 2.

The four FHbp variants are 1.4, 1.13, 2.19 and 3.45, but each have been specifically mutated to disrupt their binding interaction with human complement component Factor H (see 3.4.1). These mutagenised variants are FHbp1.4(H248L), FHbp1.13(H248L), FHbp2.19(L130R G133D) and FHbp3.45(E254A).

| GM *N. lactamica* strain | NadA | FHbp 1.4 (H248L) | FHbp 1.13 (H248L) | FHbp 2.19  (L130R G133D) | FHbp 3.45  (E254A) |
| --- | --- | --- | --- | --- | --- |
| Nlac 4NFA313_H | ✓ | ✓ |  |  |  |
| Nlac 4NFB313_H | ✓ |  | ✓ |  |  |
| Nlac 4NFC313 | ✓ |  |  | ✓ |  |
| Nlac 4NFD313_E | ✓ |  |  |  | ✓ |

Table 2: Expression of meningococcal antigens by each of the 4xrNlac GM-Nlac strains

#### 4.4 Deliberate release of 4xrNlac – safety considerations

##### 4.4.1 Potential for increased virulence of GM-Nlac strains

The major difference between Nmen and Nlac, and the primary virulence determinant of the former, is the expression of capsular polysaccharide by Nmen. The main role of this capsule is to offer the meningococcus significant protection against killing by normal human serum, enabling the bacterium to survive within the bloodstream of naïve or unvaccinated individuals. The genetic modification of the Nlac strains in 4xrNlac does not confer the ability to synthesise and deposit a polysaccharide capsule on their surface.

While the presence of the *nadA* gene in the genome is associated with hypervirulent lineages of Nmen, the expression of NadA is dispensible for pathogenesis (Section 3.3.3). NadA expression by 4NB1 in Lactamica 4 was not associated with any disease in colonised participants (Section 3.3.3).

FHbp expression by the meningococcus is not prerequisite for pathogenesis and is dispensable during infection, however the established function of FHbp during an episode of IMD is to bind human complement Factor H (hFH). This has the effect of inhibiting the activation of the complement cascade via the alternative pathway at the bacterial surface, which in turn reduces the intensity of complement-mediated killing of the bacterium in the bloodstream. No other functions for FHbp beyond this immunomodulatory role have been satisfactorily demonstrated, meaning that heterologous expression of this antigen in Nlac is not expected to result in changes to the trophic requirements of the bacterium, nor to the interactions it makes with host cells or other bacterial species resident in the nasopharynx. We therefore expect there to be no significant changes to the niche, lifestyle or tropism of genetically modified FHbp-expressing Nlac, and therefore that any risks to colonised human participants posed by heterologous expression of FHbp in Nlac are negligible. As a further precaution however, and considering published evidence that FHbp can be specifically mutated to disrupt its ability to bind human complement Factor H with no detriment to its immunogenicity, the decision was made to abrogate the hfH-binding ability of the expressed FHbp variants to be no more than a wild-type Nlac strain.

Therefore, there is no theoretical reason why the GM-Nlac strains should have increased virulence in comparison to wild-type Nlac.

##### 4.4.2 Pre-clinical testing

The four strains contained in 4xrNlac have undergone extensive pre-clinical testing which has demonstrated:

(i) Each strain expresses moderate to high levels of NadA and FHbp on its surface, in comparison to the wild-type equivalent strain 4YB2
(ii) Each strain has been demonstrated to have no more hFH binding than the wild-type equivalent strain 4YB2. In contrast, other GM-Nlac strains expressing wild-type FHbp variants (i.e. not the mutagenised FHbp variants expressed by the 4xrNlac strains) showed a significantly increased level of hFH binding
(iii) During a period of incubation with HEp-2 cells, binding of the four 4xrNlac strains to the HEp-2 cells was demonstrated, but with no increase in internalisation, in comparison to the wild-type equivalent strain 4YB2, suggesting that the expression of NadA and FHbp does not increase the ability of Nlac to invade cells to which it binds
(iv) All 4xrNlac strains remain refractory to horizontal gene acquisition from Nmen indicating that there is no increased risk of these strains acquiring the genes necessary for the synthesis and deposition of extracellular capsule, the key virulence determinant of Nmen
(v) All 4xrNlac strains remain acutely susceptible to complement-mediated killing
(vi) All 4xrNlac strains have confirmed susceptibility to front-line antibiotics (Table 3: Antimicrobial susceptibility of GM-Nlac strains)

| Strain | MIC (mg/L) | | | |
| --- | --- | --- | --- | --- |
|  | Benzylpenicillin | Ceftriaxone | Rifampicin | Ciprofloxacin |
| Nlac 4NFA313_H | 0.25 | <0.002 | 0.5 | 0.006 |
| Nlac 4NFB313_H | 0.25 | <0.002 | 0.5 | 0.006 |
| Nlac 4NFC313 | 0.25 | <0.002 | 0.5 | 0.006 |
| Nlac 4NFD313_E | 0.25 | <0.002 | 0.5 | 0.006 |
| Nlac 4YB2  (wild-type equivalent) | 0.25 | <0.002 | 0.5 | 0.006 |

Table 3: Antimicrobial susceptibility of GM-Nlac strains

Minimal Inhibitory Concentration (MIC) breakpoints for *Neisseria meningitidis* are: benzylpenicillin = > 0.25 mg/L, rifampicin = >1 mg/L, ciprofloxacin = >0.06 mg/L and ceftriaxone = >0.12 mg/L.

Following consideration of all pre-clinical data, the four GM-Nlac strains contained within the inoculum 4xrNlac are not expected to have any increased pathogenicity or risk to human health in comparison to the wild-type organism. All data have been reviewed in full by DEFRA who have given approval for the deliberate release of these four strains in this pilot study and a further randomised placebo-controlled CHI with rechallenge (DEFRA REF 25/R50/01).

##### 4.4.3 Antibiotic eradication of colonisation

Challenge participants will receive oral antibiotics to clear proven or potential colonisation at the end of the study. Any contact participants who are found to be colonised will also receive antibiotic clearance. As shown in Table 3Table 1 above, all strains contained in 4xrNlac have confirmed susceptibility to ceftriaxone, rifampicin and ciprofloxacin.

In our prior studies, a single dose of oral ciprofloxacin has been used to clear Nlac colonisation with 100% of participants cleared within 48 hours of administration, including the GM-Nlac strains 4NB1 and 4YB2 (25, 57). The recent MHRA warning regarding ciprofloxacin use is noted, with the recommendation that fluroquinolones should only be prescribed where other commonly recommended antibiotics are inappropriate, due to the risk of long-lasting adverse drug reactions including tendonitis (58). However, the risk of complications following a single dose of ciprofloxacin as meningococcal prophylaxis is considered to be extremely small and hence is still recommended in UKHSA guidance (59). Other potential antibiotic agents were considered such as rifampicin, which would require a longer course and interferes with the oral contraceptive pill, and parenteral ceftriaxone, which would involve the risks associated with venous access.

A single dose of oral ciprofloxacin is therefore felt to be the safest and most effective method to clear GM-Nlac colonisation. To mitigate the very low risk associated with this, participants will be screened for any contra-indication to ciprofloxacin (60) such as hypersensitivity to quinolones, a history of tendon disorders related to quinolone use, epilepsy, a personal or family history of aneurysm or congenital heart valve disease, and prolonged QT interval, with an ECG check at screening.

##### 4.4.4 Risk of onward transmission

As discussed in section 3.1.5, the highest risk of transmission of *Neisseria* species from a colonised individual is to household members, and in particular bedroom-sharers. Considering Nlac specifically, some studies have suggested that such household transmission may occur chiefly from infants and children rather than from adults (20, 41).

In previous CHI studies, we have assessed shedding of Nlac by microbiological analysis of face mask and exhaled air samples, as well as onward transmission to bedroom sharers and neonates. No shedding was detected in any samples taken from participants known to be colonised with wild-type Nlac (25) or with the GM-Nlac strains 4NB1 and 4YB2 (57). No onward transmission was detected from 4NB1 or 4YB2 colonised participants to their bedroom-sharers (57), or from wild-type Nlac colonised mothers to their offspring during the peri-natal period (41).

To mitigate this low risk of onward transmission, bedroom-sharers of participants receiving the inoculum will be enrolled as “contact participants” to assess for transmission (section 7.2.2). All participants, including these contact participants, will be screened according to eligibility criteria (section 7.6) to ensure that neither they, nor any close contacts, might be more vulnerable to Nlac disease (i.e. immunocompromised), and will be required to adhere to strict infection control rules (section 7.5.2). Study specific training and standard operating procedures will also be in place to minimise any potential for transmission within the laboratory or clinical areas (sections 9.4 and 9.5).

Onward transmission to other contacts, or into the wider community, is extremely unlikely and will be minimised by the strategies outlined above. It is not feasible to routinely monitor further onwards transmission through the wider community. However, there is no reason to anticipate that the GM-Nlac strains would propagate widely, relative to wild-type Nlac.

##### 4.4.5 Public health concerns and emergency response

In the unlikely event that unexpected spread occurred beyond participants enrolled in the study, the GM-Nlac strains are not expected to cause any disease. However, in the extremely unlikely event of any action being required, public health authorities would have the option of using the same strategy that is used in outbreaks of meningococcal disease, i.e. single dose ciprofloxacin to clear colonisation in close contacts (59), or alternatively to vaccinate with the NadA and FHbp containing vaccine Bexsero, which has been shown to protect against the occurrence of invasive meningococcal disease (61).

#### 4.5 Rationale for the current study

This study is designed as a pilot study to establish the safety and colonisation potential of controlled human infection with 4xrNlac in a small cohort of participants, with an open-label study design. Early immunological data will be assessed, and participant samples will be used to optimise microbiological and immunological assays. Efficacy of antibiotic clearance will also be assessed.

This pilot study will inform the design of “**The GM-Nlac study – Main**”, a double-blind randomised study of inoculation with 4xrNlac versus placebo with subsequent 4xrNlac rechallenge. In this future study we will investigate whether an episode of nasopharyngeal colonisation with a strain or strains of GM-Nlac expressing NadA and FHbp leads to a given participant becoming ‘protected from colonisation with 4xrNlac’, following experimental rechallenge. The primary outcome measure will be to observe and compare the proportion of participants who become colonised by one or more of the bacterial strains comprising 4xrNlac following challenge and, where appropriate, rechallenge. If a ‘protected from colonisation following homologous rechallenge’ phenotype is evident, then it implies 4xrNlac might have utility in preventing natural acquisition of strains of Nmen circulating in the community, on the basis that Nlac and Nmen are closely related and share several similar and immunologically cross-reactive surface structures. Indeed, the purpose of the genetic modification is to further enhance the structural similarity between these organisms without increasing the pathogenic potential of the recipient species.

The impact of reducing the meningococcal carriage frequency would most plausibly be to break meningococcal transmission chains, lowering the frequency with which susceptible individuals become exposed to the meningococcus, and therefore reducing the incidence of invasive meningococcal disease in the community. To realise this vision, and with a view to informing the rational design of future vaccines capable of generating so-called ‘herd protection’ against Nmen, we will collect longitudinal measurements of serological and cell-mediated adaptive immune responses to look for correlations and to subsequently disambiguate the mechanisms that underpin this protected phenotype.

### 5 Objectives and endpoints

#### 5.1 Co-primary objectives

1. To establish the safety of nasal inoculation of healthy adult participants with four strains of genetically modified *Neisseria lactamica* expressing FHbp and NadA
2. To demonstrate successful induction of nasopharyngeal colonisation following nasal inoculation with genetically modified *Neisseria* *lactamica* expressing FHbp and NadA

#### 5.2 Secondary objectives

1. To demonstrate successful clearance of nasopharyngeal colonisation with genetically modified *Neisseria lactamica* following a single 500mg dose of oral ciprofloxacin
2. To assess for any onward transmission of GM-Nlac colonisation to bedroom sharers of challenge participants

#### 5.3 Exploratory objectives

1. To compare colonisation kinetics of the four genetically modified *Neisseria lactamica* strains among colonised challenge participants
2. Development and optimisation of cellular immunological assays to determine T and B cell responses to GM-Nlac colonisation in whole blood and within the nasopharyngeal mucosa
3. Comparison of nasal wash and throat swabs samples to assess GM-Nlac colonisation density using qPCR

#### 5.4 Safety endpoints

1. Occurrence of solicited and unsolicited adverse events within the study period
2. Occurrence of serious adverse events within the study period

#### 5.5 Microbiological endpoints

1. Culture of *N. lactamica* from throat swabs and nasal washes taken at each follow up visit after inoculation
2. qPCR to identify the relative density of each GM-Nlac strain from throat swabs and nasal washes taken at each follow up visit after inoculation

#### 5.6 Immunological endpoints

1. Meningococcal serum bactericidal activity
2. Serum IgG specific for each FHbp variant and NadA
3. Mucosal IgA specific for each FHbp variant and NadA

### 6 Description and justification of the study design

#### 6.1 Overview

This is a prospective, single-arm, open label, controlled human infection model experiment, in which ten healthy adult volunteers will be nasally inoculated with 4xrNlac. This is a multi-strain challenge agent containing approximately equal doses of four genetically modified (GM) strains of *Neisseria lactamica* (Nlac), each expressing the meningococcal antigen *Neisseria* adhesin A (NadA) and one of four variants of the meningococcal antigen Factor H binding protein (FHbP) on its surface (section 3.3.4).

Following inoculation, participants will be followed up over 28 days to review safety, nasopharyngeal colonisation with the GM-Nlac strains and immunogenicity. A single dose of oral ciprofloxacin will be administered at day 28 with a final visit at day 30 to confirm clearance of colonisation. Bedroom contacts will be enrolled to look for transmission of the GM-Nlac strains.

The aims of this study will be to establish that nasopharyngeal colonisation with the GM-Nlac strains can be induced safely in an adequate fraction of participants, and successfully cleared with oral antibiotic treatment, with equivalence to that seen following inoculation with wild-type Nlac (25), and the GM-Nlac strains 4YB2 and 4NB1 in our previous studies (57).

#### 6.2 Challenge and contact participants

Volunteers who receive an intranasal inoculum of 4xrNlac will be enrolled as “Challenge participants”. Challenge participants will attend a screening visit to complete informed consent and medical screening, a pre-challenge visit approximately 5 days prior to inoculation, and a challenge visit at which they will receive the intranasal inoculum of 4xrNlac. Following inoculation, they will attend regular outpatient visits over 30 days to assess safety, colonisation and immunogenicity, with antibiotic administration to clear colonisation at day 28 post inoculation, regardless of colonisation status. The schedule of procedures for challenge participants is detailed in Table 4 on p30.

To assess for any onward transmission of the GM strains, individuals who share a bedroom with challenge participants will be enrolled as “Contact participants”. Contact participants will be screened and consented prior to inoculation of the challenge participant, with a maximum of one contact participant per challenge participant. Challenge participants will not be eligible for the study if they intend to share a bedroom with anyone who is ineligible or unwilling to be enrolled as a contact participant. Contact participants will attend for a single visit at day 28 post inoculation of the challenge participant, at which a throat swab will be taken to assess for colonisation with the GM-Nlac strains. Contact participants will also be screened for GM-Nlac colonisation at an additional visit if symptoms suggestive of respiratory or systemic infection occur during the study period, or if they, or their corresponding challenge participant, withdraw prior to the end of the study. A single dose of ciprofloxacin will be administered for the contact participant to take home, and if the throat swab is positive for GM-Nlac then the contact participant will be instructed to take the ciprofloxacin and invited to a further visit to confirm clearance of colonisation. The schedule of procedures for contact participants is detailed in Table 5 on page 31.

| **Visit** | | **Scr** | **PC** | **C0** | **C1^a^** | **C4** | **C7** | | **C14** | **C28** | **PE^b^** |
| --- | --- | --- | --- | --- | --- | --- | --- | --- | --- | --- | --- |
| Window (Days) | | C0 -5-90 | C0 -5-10 | 0 | C0 +1 | C0 +3-5 | C0 +6-8 | | C0 +12-16 | C0 +25-31 | 1-4 post abx |
| Visit Type | | Screening | Pre-challenge | Challenge | Surveillance | | | | | Eradication | Post eradication |
|  |  |  |  |  | Daily diary card | | |  | |  |  |
|  | TOPS | x |  |  |  |  |  | |  |  |  |
|  | PCQ | x |  |  |  |  |  | |  |  |  |
| **Challenge participant** | Informed consent | x |  |  |  |  |  | |  |  |  |
|  | Request GP records  / electronic medical records | x |  |  |  |  |  | |  |  |  |
|  | Medical history | x |  |  |  |  |  | |  |  |  |
|  | Physical examination | x |  |  |  |  |  | |  |  |  |
|  | Height and weight | x |  |  |  |  |  | |  |  |  |
|  | Reaffirm consent |  | x | x | x | x | x | | x | x | x |
|  | Reconfirm eligibility |  | x | x |  |  |  | |  |  |  |
|  | Review of adverse events & concomitant medications |  | x | x | x^c^ | x^c^ | x^c^ | | x | x | x |
|  | Targeted history and examination |  | (x) | (x) |  | (x) | (x) | | (x) | (x) | (x) |
|  | Vital Signs | x | x | x |  | x | x | | x | x | x |
|  | Urinalysis | x |  |  |  |  |  | |  |  |  |
|  | Urine toxicology | x |  |  |  |  |  | |  |  |  |
|  | Pregnancy Test *(females only)* | x |  | x |  |  |  | |  | x |  |
|  | ECG | x |  |  |  |  |  | |  |  |  |
|  | Throat swab x 1-2 | x | x |  |  | x | x | | x | x | x |
|  | Nasal wash |  |  |  |  | x | x | | x | x | x |
|  | Nasosorption sample |  | x |  |  | x | x | | x | x |  |
|  | Inferior turbinate swab |  | x |  |  | x | x | | x | x |  |
|  | Saliva sample |  | x |  |  | x | x | | x | x |  |
|  | Safety bloods | 8 |  | 8 |  | 8 | 8 | | 8 | 8 |  |
|  | Research bloods (maximum ml) |  |  | 60 |  |  |  | | 50 | 60 |  |
|  | Maximum total blood | 8 |  | 76 |  | 84 | 92 | | 150 | 218 |  |
|  | Inoculation |  |  | x |  |  |  | |  |  |  |
|  | Eradication (Ciprofloxacin 500 mg) |  |  |  |  |  |  | |  | x |  |
|  | CRF/ eCRF completion | x | x | x |  | x | x | | x | x | x |

Table 4: Schedule of procedures – The GM Nlac Study – Pilot: Challenge participants (x) if clinically indicted / optional, ^a^Telephone contact, ^b^PE visit planned for Day 30 but if early antibiotics are required then it will occur 1-4 days following antibiotic administration, ^c^Including diary card review

| **Visit** | | **Scr** |  | **C28** | | **PE^a^** |
| --- | --- | --- | --- | --- | --- | --- |
| Window (Days) | | C0 - 2-90 |  | C0 + 25-31 | | 1-4 post abx |
| **Contact participant** | PCQ, TOPS, Informed consent | x | 24-hour telephone number for study team, additional visit with throat swab and other investigations as clinically indicated if symptoms suggestive of respiratory or systemic infection, early antibiotics if found to be colonised | |  |  |
|  | Reaffirm consent |  |  |  | x | (x) |
|  | Medical history and examination including Ht and Wt | x |  |  |  |  |
|  | Review of adverse events & concomitant medications |  |  |  | x | (x) |
|  | Targeted history and examination |  |  |  | (x) | (x) |
|  | Vital Signs | x |  |  | x | (x) |
|  | Pregnancy Test *(females only)* | x |  |  | x |  |
|  | ECG | x |  |  |  |  |
|  | Throat swab x 1-2 | x |  |  | x | (x) |
|  | Eradication (Ciprofloxacin 500 mg) TTO^b^ |  |  |  | x |  |
|  | CRF/ eCRF completion | x |  |  | x | (x) |

Table 5: Schedule of procedures – The GM-Nlac study – Pilot: Contact participants

(x) if clinically indicted / optional, ^a^Contact participants will only attend PE visit if antibiotics are indicated. ^b^Antibiotic eradication therapy will be supplied as a TTO at the C28 visit but contact participants will only be instructed to take them if their C28 throat swab is positive

#### 6.4 Safety considerations

An independent Data and Safety Monitoring Board (DSMB) will be established prior to the start of the study to provide oversight of safety and study conduct (section 11.5).

Although unlikely, we recognise that onward transmission to other human contacts is possible. In order to minimise the risk of transmission, both challenge and contact participants will be required to abide by infection control rules (section 7.5.2), including abstinence from intimate contact or bedroom sharing with any individual other than those consented and enrolled as their bedroom-sharer. These rules will form part of a pre-consent questionnaire, and the written informed consent form which will be signed by all participants prior to enrolment. Potential challenge and contact participants will not be enrolled to the study if they have any household, regular occupational or close social contact with individuals known to be immunocompromised, or pre-school children, during the study period.

As a first-in-human study, participants will be inoculated according to the following schedule. Following each set of inoculations, a review of safety data up to and including the day 7 visit will be conducted.

- Challenge participant 1 – inoculated individually
- Challenge participants 2-5 – inoculated in groups of 1-2
- Challenge participants 6-10 – inoculated in groups of up to 5

The decision to inoculate the next group of challenge participants will be taken by the investigators, with discussion with the DSMB if there are any safety concerns.

Challenge and contact participants will be given a 24-hour telephone number to contact the study team if they become unwell during their enrolment in the study. During the immediate post-challenge period there will be close contact between the challenge participant and study team to monitor for the development of any symptoms. This will include a planned telephone contact on day 1 post challenge (section 8.3.1), daily safety data collected via a paper diary card (section 8.3.2) and face-to-face visits at days 4 and 7 post challenge (section 8.3.3). The diary card will be reviewed at the day 4 and 7 visits, but participants will be instructed to contact the study team if they develop any grade 3 solicited symptoms or any other concerns in between planned visits.

Throughout the study, additional visits will be arranged if required, with testing for GM-Nlac colonisation, additional investigation if required, and early antibiotic treatment if GM-Nlac disease is suspected by the study team (section 8.4.4). Study holding criteria are detailed in section 8.5.3.

#### 6.5 Sample size

The planned sample size is ten challenge participants. This has been chosen to allow an estimation of colonisation fraction and efficacy of antibiotic eradication therapy, and to provide initial safety data which will be used to inform a follow-up double-blind, randomised controlled human infection study comparing 4xrNlac to placebo control.

Challenge participants who are considered by the study-steering committee to be non-evaluable for safety and colonisation endpoints may be replaced. Challenge participants may be considered non-evaluable if they:

- Receive an inadequate nasal inoculation (see section 8.2.4)
- Do not complete adequate follow up to assess safety and colonisation status (i.e. inability to provide adequate microbiological samples or withdrawal prior to the end of the study)
- Receive antibiotics likely to clear GM-Nlac colonisation prior to C28, either due to safety concerns as per protocol, or for non-study related reasons
- Acquire Nmen nasopharyngeal colonisation during the study period as this is likely to impact Nlac colonisation efficacy

A maximum of one contact participant may be enrolled per challenge participant, therefore there will be up to a maximum of ten contact participants enrolled. Contact participants will only be enrolled if the challenge participant has a regular bedroom sharer at the start of the study, and therefore there will be no minimum target for contact participant enrolment. If additional challenge participants are enrolled as replacements as described above, then they may each have a maximum of one contact participant enrolled.

#### 6.6 Randomisation and blinding

There will be no randomisation in this single arm open-label pilot study. Laboratory staff performing meningococcal serum bactericidal assays will be blinded to the colonisation status of challenge participants.

#### 6.7 Definition of the start and end of the study

The start of the study is defined as the date of screening of the first participant. The clinical component of the study is expected to be completed within one year of the start of recruitment. However, the study will remain open for a period of 10 years to allow for ongoing clinical sample storage and further ethically approved sample processing and analysis.

#### 6.8 Duration of participation

The duration of involvement of challenge and contact participants in the study will be from the screening visit (up to 90 days prior to inoculation) until 30 days after inoculation.

#### 6.9 Potential benefits for participants

Participants will not benefit directly from enrolment in this study. However, it is envisaged that the information gained from this study will contribute to knowledge about nasal colonisation with GM-Nlac expressing NadA and FHbp, the development of systemic and mucosal immunity to Nmen, and protection against Nmen colonisation. In the future this may lead to the development of safe and effective protective strategies against meningococcal infection. Participants will also receive information about their general health status.

#### 6.10 Regulatory approval

This study has received approvals from the DEFRA for the deliberate release of the multi-strain challenge agent (DEFRA REF 25/R50/01). All necessary approvals from the National Research Ethics Service (NRES) and Health Research Authority (HRA) (REC Reference 26/SC/0072 the University of Southampton as Sponsor (ERGO 81849), and University Hospital Southampton R&D (RHM MED 2124) will be in place before this study is commenced.

The study will be conducted at the Southampton NIHR Clinical Research Facility (NIHR-CRF) at University Hospital Southampton according to Good Clinical Practice (GCP) and according to documented legal and local procedures and guidelines prior to study initiation. All study staff will be given study-specific training.

### 7 Inoculum

#### 7.1 4xrNlac

4xrNlac is a multi-strain challenge agent, comprised of four different GM-Nlac strains. Each strain expresses NadA and one of four different variants of FHbp which have been specifically mutated to disrupt their binding interaction with human complement component Factor H (see 3.4.1). The four strains and the heterologous antigens that they express are summarised in Table 2 on p19.

#### 7.2 Manufacture and storage of the inoculum

Aliquots of the inoculum strains will be grown and prepared in the Medical School laboratory LC70 at the University of Southampton by Dr Jay Laver, following the standard operating procedures **GMNL07 (Lab) - Growth and preparation of genetically modified *Neisseria lactamica* for use as inocula in human challenge experiments** and **GMNL11 (Lab) - Dilution of MASTER Stocks of genetically modified Neisseria lactamica to INOCULUM stocks, suitable for use as inocula in controlled human infection model experiments**. Master and inoculum stocks will be stored in a -80^o^C freezer in secured facilities within the University of Southampton with restricted access. A chain of accountability will be recorded at a GMP-like standard.

#### 7.3 Quality assessment of the inoculum

Master and inoculum stocks will be assessed for viability and contamination using standard microbiological procedures and according to the standard operating procedures **GMNL08 (Lab) - Quantification and microbiological purity assessment of frozen MASTER stocks of genetically modified *Neisseria lactamica*** and **GMNL12 (Lab) - Quantification and microbiological purity assessment of frozen INOCULUM stocks of genetically modified *Neisseria lactamica***. A chain of accountability will be recorded at a GMP-like standard.

### 8 Study setting and recruitment

#### 8.1 Study site

The study will be conducted in the Southampton NIHR-CRF, at University Hospital Southampton (UHS), Tremona Rd, Southampton, SO16 6YD. Relevant facilities at the Southampton NIHR-CRF include 13 consulting rooms (2 configured for infectious participants); a containment level 2 laboratory, two environmental laboratories with containment level 2 environmental chambers; state-of-the-art physiological monitoring and physical and management systems to ensure Regulatory Compliance such as computerised sample inventory, and tracking system (https://www.southamptoncrf.nihr.ac.uk/our-facilities).

#### 8.2 Recruitment

Care will be taken not to recruit from vulnerable groups (mental health or other capacity issues or those under 18 years old).

##### 8.2.1 Challenge participants

Challenge participants will be recruited through various media and by use of an advertisement +/- registration form formally approved by the ethics committee and distributed or posted in the following places:

- In public places, including buses and trains, with the agreement of the owner / proprietor
- In newspapers or other literature for circulation inclusive of digital media used by these outlets
- On radio via announcements
- On a website operated by our group or with the agreement of the owner or operator (including online recruitment through our website). We intend to use several different websites which advertise clinical trials and health research including CRF, BRC, Be Part of Research.
- As a post on a Twitter, Facebook or Gumtree account owned and operated by our group or on social media private groups with the permission of the Group Moderator
- By e-mail distribution to a group or list only with the express agreement of the network administrator or with equivalent authorisation and according to the relevant communications policy
- By email distribution to individuals who have already expressed an interest in taking part in any clinical study at the NIHR-CRF Southampton
- On stalls or stands run by Southampton NIHR CRF or BRC, The University of Southampton or Southampton Controlled Human Infection Group at exhibitions and fairs with the permission of the event organiser
- Via presentations (e.g. presentations at lectures or invited seminars)
- Southampton NIHR-CRF Database of Healthy Volunteers: We may contact individuals from this database who have previously expressed an interest in receiving information about future studies for which they may be eligible
- NIHR Be Part of Research website and/ or database
- SMS (Text messages) sent out via GP surgeries – use of PIC sites
- Use of a healthy volunteer database owned and managed by hVIVO where relevant approvals and contracts are in place

##### 8.2.2 Contact participants

All contact participants will be bedroom sharers of challenge participants defined as an individual who will share a bedroom with a challenge participant on at least one occasion during the study period. Challenge participants will be asked to inform their bedroom contacts of their interest and potential involvement in this study and provide contact details for the study team. Bedroom sharers who are interested in being enrolled as contact participants will contact the study team for further information and to arrange a contact participant screening visit.

#### 8.3 Participant information sheets

##### 8.3.1 Challenge participant information sheet

A challenge participant information sheet will be made available to interested healthy volunteers at least 24 hours before their screening visit. This information sheet will include all risks of participating in this study and safety measures that are involved in the study and will be formally approved as part of the REC and HRA application.

##### 8.3.2 Contact participant information sheet

A contact participant information sheet will be made available to bedroom sharers of potential challenge participants at least 24 hours prior to the contact participant screening visit. This information sheet will include information about the study and contact participant involvement as well as potential risks and benefits and will be formally approved as part of the REC and HRA application.

#### 8.4 Pre-screening procedures

Potential challenge and contact participants will have a brief telephone pre-screening prior to their face-to-face medical screening appointment. This pre-screening will collect demographic and basic medical information using a REC approved proforma. Verbal consent to collect this information will be taken and documented on the proforma.

#### 8.5 Screening visit

Individuals who have expressed an interest in taking part in the study as a challenge or contact participant will be invited to attend a screening visit. The screening visit can be up to 90 days prior to the challenge visit (C0). Challenge and contact participant screening visits can take place together or separately. Screening visit procedures are summarised in Table 4 on p30 for challenge participants and Table 5 on p31 for contact participants, with further detail given in the sections below.

##### 8.5.1 Informed consent and pre-consent questionnaire

The study aims, participant involvement and obligations, study procedures, risks and benefits, safety measures and infection control procedures (Table 6, page 38) will be explained to the participant by a study doctor, with an opportunity to ask questions, and time to consider whether or not to participate.

The following will be emphasised:

- Participation in the study is entirely voluntary
- Refusal to participate involves no penalty or loss of medical benefits
- The participant may withdraw from the study at any time
- The participant is free to ask questions at any time to allow him or her to understand the purpose of the study and the procedures involved
- There is no direct benefit from participating
- The participant will be registered on the TOPS database (The Over-volunteering Prevention System; www.tops.org.uk)

If they wish to participate, both challenge and contact participants will be asked to complete a pre-consent questionnaire testing their understanding of the study. Incorrect answers will be explored and further explained to ensure full understanding of the study, and the participant will be asked to re-attempt the question. Once each question has been answered correctly with understanding demonstrated to the investigator, then the participant will be asked to sign and date the informed consent form, which will also be signed and dated by the investigator. The original ICF and pre-consent questionnaires will be stored in the ISF. Copies of the ICF will be stored in the participant’s medical file and given to the participant. The informed consent form will be completed before any study specific procedures are performed for that participant. If the challenge participant has a bedroom sharer, then they will be screened, consented, and declared eligible as a contact participant prior to the challenge visit of the challenge participant.

##### 8.5.2 Infection control guidelines

To mitigate the theoretical risk of onward transmission to other contacts, both challenge and contact participants will be asked to abide by infection control guidelines from the challenge visit of the challenge participant (C0) until the completion of their participation in the study. These guidelines will be detailed in the challenge and contact participant information sheets and explained verbally at the screening appointment. Understanding of these guidelines will be assessed by the investigator as part of the pre-consent questionnaire, and agreement to abide by these guidelines will be included in the informed consent form for both challenge and contact participants.

| Infection Control Guidelines | |
| --- | --- |
| 1 | Participants must wash their hands before leaving their home and following any contact with respiratory secretions e.g. sneezing / blowing nose |
| 2 | Participants must be contactable by mobile phone, which has the study emergency phone number programmed in, and contact the clinical study team if they have any symptoms suggestive of respiratory or systemic infection |
| 3 | Participants must be able to be return to the NIHR-CRF within 24 hours |
| 4 | Participants must avoid attending heavily crowded environments such as  nightclubs for the two weeks following inoculation of the challenge participant |
| 5 | Participants must avoid close contact with individuals known to be potentially vulnerable for the duration of their involvement in the study.  Close contact is defined as:   - Face to face contact < 2m for >15 minutes - Staying overnight in the same accommodation   Potentially vulnerable individuals include:   - Immunocompromised individuals - Children < 5 years |
| 6 | Participants must avoid activities associated with a high risk of transmission  with any individuals other than their declared and consented bedroom-sharer.  High risk of transmission activities include:   - Bedroom sharing - Intimate/sexual contact - Contact that may involve transfer of respiratory secretions e.g. kissing - Sharing unwashed cutlery or drinking vessels |
| 7 | Participants must avoid dental examinations and treatments where possible. If emergency dental treatment is required, the participant should inform the dentist about their involvement in the study |

Table 6: Infection Control Guidelines

##### 8.5.3 Electronic medical records and communication with the General Practitioner

Written consent will be taken to access electronic medical records and communicate with the participant’s General Practitioner (GP) regarding their potential involvement in this study and their medical history. A letter describing the study and the participant’s involvement will be sent to the GP following the screening visit. This will include contact details for the research team and the GP will be invited to ask questions or raise any concerns about the participant’s enrolment prior to the challenge visit. For challenge participants only, if an electronic record of medical and immunisation summary is not accessible then a print-out of the medical summary will be requested.

##### 8.5.4 Medical history and physical examination

A detailed medical history and physical examination will be conducted, making sure that all inclusion criteria and no exclusion criteria are met.

##### 8.5.5 Screening investigations

Screening investigations will be carried out to confirm eligibility as detailed in Table 4 on page 30 for challenge participants and Table 5 on page 31 for contact participants.

All participants will be screened for baseline Nlac and Nmen colonisation. For potential challenge participants this will include a throat swab and nasal wash culture, whereas potential contact participants will only have a throat swab taken. All participants will have a screening ECG due to the use of ciprofloxacin to clear colonisation. All female participants of child-bearing potential will have a urine pregnancy test performed at screening and prior to challenge and antibiotic eradication if applicable. Challenge participants will have additional blood and urine samples taken which will be sent to the UHS clinical laboratory for analysis. Laboratory parameters will be considered according to UHS laboratory normal reference ranges shown in Table 11 on p58. Results that fall outside of these ranges may not be of clinical significance and will be considered on an individual basis.

#### 8.6 Inclusion and exclusion criteria

##### 8.6.1 Inclusion criteria – Challenge participants:

The challenge participant must satisfy all the following inclusion criteria to be eligible for the study:

1. Healthy adults aged 18 to 45 years inclusive on the day of enrolment
2. Fully conversant in the English language
3. Able and willing (in the investigator’s opinion) to comply with all study requirements
4. Able to correctly answer all questions in the pre-consent questionnaire
5. Provide written informed consent to participate in the study including agreement to abide by infection control guidelines during the study period
6. Provide written consent to allow the study team to discuss the participant’s medical history with the General Practitioner and access electronic medical records
7. Written informed consent provided by any bedroom sharer who is eligible to be enrolled as a contact participant (if applicable)
8. For females of child-bearing potential, a negative pregnancy test on the days of screening and challenge
9. Use of continuous effective contraception during the study (section 7.6.5)
10. Agreement to take antibiotic eradication therapy according to the study protocol
11. TOPS registration completed and no conflict found

##### 8.6.2 Exclusion criteria – Challenge participants:

The challenge participant may not enter the study if any of the following criteria apply:

1. Individuals who have a current infection at the time of inoculation
2. Use of systemic antibiotics within the period 30 days prior to inoculation
3. Current active smokers defined as having smoked or vaped in the week prior to inoculation
4. *N. lactamica* or *N. meningitidis* detected on throat swab or nasal wash taken at screening or at the pre-challenge visit
5. Individuals who have been challenged with wild-type or GM-Nlac in a previous controlled human infection study
6. Individuals who have received one or more doses of a meningococcus B vaccine (Bexsero or Trumenba)
7. History of allergy or intolerance to any component of the inoculum
8. Contraindications to the use of ciprofloxacin, specifically hypersensitivity to quinolones, a history of tendon disorders related to quinolone use, epilepsy, a personal or family history of aneurysm or congenital heart valve disease, and prolonged QT interval
9. Contraindications to the use of ceftriaxone, specifically known hypersensitivity to cephalosporins or severe penicillin allergies (e.g. anaphylaxis or Stevens Johnson Syndrome)
10. Any confirmed or suspected immunosuppressive or immune-deficient state, specifically terminal complement component deficiencies, eculizumab use, known HIV infection, malignancy, asplenia, recurrent severe infections and chronic (more than 14 days) immunosuppressant medication within the past 6 months (topical steroids are allowed)
11. Use of immunoglobulins or blood products within 3 months prior to enrolment
12. Household, close social or regular occupational contact with persons known to be immunosuppressed, specifically HIV infection with a CD4 count <200 cells/mm3; asplenia; any malignancy, recurrent, severe infections and chronic (more than 14 days) immunosuppressant medication within the past 6 months (topical steroids are allowed)
13. Household or regular occupational contact with children under 5 years or an older child with a tendency to co-sleep with the participant
14. Any abnormal finding on clinical examination or screening investigations assessed by the investigator to be clinically significant. In the event of abnormal test results, confirmatory repeat tests may be requested.
15. Pregnancy, lactation or intention to become pregnant during the study
16. Any other significant disease, disorder, or finding which may significantly increase the risk to the participant, affect their participation in the study or impair interpretation of the study data, for example recent surgery to the nasopharynx

##### 8.6.3 Inclusion criteria – Contact participants

The contact participant must satisfy all the following inclusion criteria to be eligible for the study:

1. Healthy adults aged 18 to 55 years inclusive on the day of enrolment
2. Fully conversant in the English language
3. Able and willing (in the investigator’s opinion) to comply with all study requirements
4. Able to correctly answer all questions in the pre-consent questionnaire
5. Provide written informed consent to participate in the study including agreement to abide by infection control guidelines during the study period
6. Provide written consent to allow the study team to discuss the participant’s medical history with the General Practitioner and access electronic medical records
7. For females of child-bearing potential, a negative pregnancy test on the days of screening and challenge
8. Use of continuous effective contraception during the study (section 7.6.5)
9. Agreement to take antibiotic eradication therapy according to the study protocol
10. TOPS registration completed and no conflict found

##### 8.6.4 Exclusion criteria – Contact participants

The contact participant may not enter the study if any of the following criteria apply:

1. Contraindications to the use of ciprofloxacin, specifically hypersensitivity to quinolones, a history of tendon disorders related to quinolone use, epilepsy, a personal or family history of aneurysm or congenital heart valve disease, and prolonged QT interval
2. Contraindications to the use of ceftriaxone, specifically known hypersensitivity to cephalosporins or severe penicillin allergies (e.g. anaphylaxis or Stevens Johnson Syndrome)
3. Any confirmed or suspected immunosuppressive or immune-deficient state, specifically terminal complement component deficiencies, eculizumab use, known HIV infection, malignancy, asplenia, recurrent severe infections and chronic (more than 14 days) immunosuppressant medication within the past 6 months (topical steroids are allowed)
4. Use of immunoglobulins or blood products within 3 months prior to enrolment
5. Household, close social or regular occupational contact with persons known to be immunosuppressed, specifically HIV infection with a CD4 count <200 cells/mm3; asplenia; any malignancy, recurrent, severe infections and chronic (more than 14 days) immunosuppressant medication within the past 6 months (topical steroids are allowed)
6. Household or regular occupational contact with children under 5 years or an older child with a tendency to co-sleep with the participant
7. Any abnormal finding on clinical examination or screening investigations, assessed by the investigator to be clinically significant. In the event of abnormal test results, confirmatory repeat tests may be requested.
8. Pregnancy, lactation or intention to become pregnant during the study
9. Any other significant disease, disorder, or finding which may significantly increase the risk to the participant, affect their participation in the study or impair interpretation of the study data

##### 8.6.5 Effective contraception

Challenge and contact participant pairs are required to use an effective form of contraception during this study. Acceptable forms of contraception include:

- Established use of oral, injected or implanted hormonal methods of contraception
- Placement of an intrauterine device or intrauterine system
- Barrier methods of contraception (condom or occlusive cap with spermicide)
- Male or female surgical sterilisation
- True abstinence when this is in line with the preferred and usual lifestyle of the participants

All challenge participants who do not have a contact participant are required to abstain from intimate contact with any other individual for the duration of the study

### 9 Conduct of the study

#### 9.1 Pre-challenge visit

Eligible challenge participants will attend for a pre-challenge visit between 5 and 10 days prior to challenge. Ongoing consent and eligibility will be confirmed, and a throat swab and nasal wash will be performed to ensure that they are still not colonised with Nlac or Nmen*.* If both are negative at this point it will be assumed that the participant is not colonised on the day of challenge (C0). Respiratory samples (nasosorption, inferior turbinate swab and saliva sample) will be taken for assessment of baseline mucosal immunological parameters. If the screening visit occurs within the 10 days prior to challenge then the pre-challenge visit can be conducted concurrently, without duplication of the microbiological samples.

#### 9.2 Challenge visit (C0)

##### 9.2.1 Challenge participant preparation

Prior to the challenge procedure, ongoing consent and understanding of the infection control guidelines will be confirmed with an opportunity to ask further questions. Eligibility will be re-confirmed with an interim medical history addressing any new medical issues since the screening visit. Special attention will be given to any possible signs of infection such as fever or respiratory symptoms. Vital signs will be checked, and a targeted physical examination will be performed if required. If there are any significant abnormalities found, the challenge may be postponed at the discretion of the study doctor. Females of child-bearing age will have a urine pregnancy test. Blood will be taken for assessment of baseline immunological parameters.

##### 9.2.2 Inoculum preparation

The inoculum will be prepared in the NIHR-CRF or UoS laboratory using a dedicated category II safety hood by technical staff according to the Standard operating procedure **GMNL17 (Lab) Preparation and monitoring of 4xrNlac inoculum for use in controlled human infection studies.** Individual inoculum doses will be prepared from a vial of GM-Nlac inoculum stock according to a dilution calculation based on the batch viability of inoculum vials, with a target dose of 100,000 (10^5^) CFU of each of the four GM strains for a total of 4 x 10^5^ CFU in 1 mL in phosphate buffered saline (PBS). More than one inoculum dose may be prepared from an individual vial if participants are being inoculated at the same time.

##### 9.2.3 Challenge procedure

The challenge procedure will be carried out in one of the containment level 2 environmental chambers within the NIHR-CRF and conducted by a study doctor with a second trained study team member. The inoculum will be administered as nose drops (500 μL into each nostril) with the participant lying supine with neck extended, according to the standard operating procedure **Performance of nasal inoculation for controlled human infection studies.**

After inoculation the participant will lie supine for at least 5 minutes and remain under observation within the NIHR-CRF for at least 15 minutes.

##### 9.2.4 Monitoring the inoculum dose

The target dose of 4xrNlac is 10^5^ CFU of each strain, so a total of 4 x 10^5^ CFU. This is based on our previous Nlac and GM-Nlac CHI studies in which 10^5^ CFU has been established as the standard inoculum dose.

A viable count will be performed on the inoculum at the time of preparation, and on the residual inoculum immediately after inoculation according to the standard operating procedure **GMNL17 (Lab) Preparation and monitoring of 4xrNlac inoculum for use in controlled human infection studies.** This will quantify the actual dose of GM-Nlac strains administered in total. A qPCR will also be performed to quantify the relative proportion of each strain.

It is anticipated that there will be some variability in the actual dose administered. Any individual inoculum found to be >5 or >10 times the intended total dose (i.e. 2 x 10^6^ or 4 x 10^6^ CFU respectively) will be reported as an adverse event, or adverse event of special interest, respectively, and will trigger reassessment of the batch viability and recalculation of the batch dilution calculation if required. The batch viability will also be checked monthly, or if there is a trend of increasing or decreasing inoculum viability over time.

An administered dose of <4 x 10^4^ CFU in total will be considered to be an inadequate inoculum. Significant loss or spillage of the inoculum may also be considered to be an inadequate inoculum at the discretion of the investigator. Challenge participants who are considered to have received an inadequate inoculum may be replaced as non-evaluable participants (section 12.1).

#### 9.3 Surveillance period

##### 9.3.1 Day 1 contact

Challenge participants will be contacted by telephone by a member of the study team on the day following their challenge (C1). During this call they will be asked to report the occurrence and severity of any solicited or unsolicited adverse events. They will be reminded to complete their daily diary card entry (section 8.3.2) and to contact the study team in the event of any grade 3 adverse events or if they have any concerns.

##### 9.3.2 Post-challenge diary

Challenge participants will be asked to complete a paper diary for the first 7 days following challenge. This will include a daily temperature check and the occurrence and severity of any solicited symptoms (Table 7), and any unsolicited adverse events. Participants will be asked to contact the study team if they have any concerns or if any grade 3 adverse events occur. Grading of adverse events will be explained at the challenge visit and included in the diary. The diary will be reviewed at the C4 and C7 visits.

##### 9.3.3 Scheduled follow up visits

Challenge participants will attend for post-challenge visits on day 4 (C4), day 7 (C7), day 14 (C14) and day 28 (C28) following inoculation for assessment of safety, colonisation and immunological parameters. At the C28 visit antibiotic eradication therapy (see section 8.4.1) will be administered to all challenge participants. They will then return for a final visit 1-4 days after the C28 visit for a post-eradication visit (PE). Procedures and clinical samples for each visit are detailed in the schedule of procedures in Table 4 on p30.

Contact participants will attend a C28 visit for assessment of safety and transmission of colonisation. This should ideally be on the same day as the challenge participant but can be any time within the window for the challenge participant C28 visit. If antibiotic eradication therapy is required then they will return for a post-eradication visit (PE) 1-4 days after they have taken the antibiotics.

##### 9.3.4 Additional safety visits

Challenge and contact participants will be given a 24-hour telephone number to contact the study team. They will be encouraged to contact the study team at any point during the study in the event of any symptoms developing. Further telephone contacts or additional safety visits may be arranged to review any participants who develop symptoms suggestive of a respiratory or systemic infection, at the discretion of the study team. Safety and colonisation will be assessed with additional samples taken and/or further review arranged if clinically indicated (8.5.1). Antibiotic eradication may be administered at the discretion of the investigator (8.5.2).

##### 9.3.5 Withdrawal visits

If a participant withdraws or is withdrawn from the study (8.6) then they will be invited to a withdrawal visit. This visit will be conducted according to the C28 schedule (Table 4 and 5), with a subsequent PE visit following antibiotic eradication where applicable (8.4.5).

##### 9.3.6 Assessment of safety

Safety data will be collected via the post-challenge diary card, and at each post-inoculation visit. Participants will be asked to report the occurrence and severity of any solicited symptoms which have occurred since the last visit (

Table 7). Any unsolicited adverse events and concomitant medication use will be recorded, and vital signs will be measured. Safety blood tests will be taken at the timepoints indicated in Table 4. If clinically indicated, then a targeted medical history and physical examination will be performed by a study doctor. In the event of safety concerns, additional laboratory tests may be conducted at the discretion of the investigator, e.g. respiratory viral PCR, blood culture. Further review at an additional visit or telephone contact may be arranged. See section 8.5 for further management in the event of safety concerns suggestive of potential GM-Nlac disease.

| Upper Respiratory Symptoms | Lower Respiratory Symptoms | Systemic Symptoms |
| --- | --- | --- |
| Rhinorrhoea | Cough | Fever or chills |
| Nasal congestion | Wheeze | Malaise |
| Sneezing | Dyspnoea (breathlessness) | Arthralgia (joint pains) |
| Sore throat |  | Myalgia (muscle pains) |
| Earache / discharge |  |  |
| Headache |  |  |

Table 7: Solicited symptoms

##### 9.3.7 Assessment of colonisation and transmission

Successful nasopharyngeal colonisation of challenge participants will be assessed by culture of throat swabs and nasal washes collected at the time points indicated in Table 4 and at any additional visits. Transmission to contact participants will be assessed by culture of throat swabs taken at the C28 visit, and any additional visits where relevant. Two throat swabs may be taken at some participant visits to allow optimisation of sampling and microbiological processes.

Clinical samples will be collected and processed according to standard operating procedures. Positive colonisation / transmission will be defined as the culture of one or more colonies of any challenge strain from a throat swab or nasal wash sample taken at any time following inoculation up to, and including, the C28 visit.

##### 9.3.8 Assessment of immunological parameters

The following clinical samples will be used to assess immunological parameters at baseline (C0 for blood and PC for respiratory mucosal samples) and at post-inoculation timepoints as indicated in Table 4, for challenge participants only.

- Blood sampling
- Respiratory samples - nasosorption strip, inferior turbinate swab and saliva samples

Venepuncture and respiratory sampling will be performed by trained, experienced study team members in accordance with clinical research facility standard operating procedures.

#### 9.4 Antibiotic eradication therapy

##### 9.4.1 Antibiotic choice

A single dose of 500mg ciprofloxacin will routinely be used to clear nasopharyngeal colonisation with the GM-Nlac strains. The rationale for this antibiotic choice is discussed in 3.4.3. This may be administered as immediate antibiotic therapy which will be taken at the study visit and directly observed by a study team member, or it will be prescribed as a delayed antibiotic therapy, to be taken by the participant at home following telephone instructions from a study doctor. A urinary pregnancy test will be performed on all female participants of child-bearing potential prior to administration of ciprofloxacin.

Alternative effective antibiotics may be used if clinically indicated, e.g. Rifampicin 600mg bd for 48 hours in the event of pregnancy or other contra-indication to ciprofloxacin becoming apparent, or ceftriaxone in a symptomatic participant if parenteral treatment were indicated. If a course of oral antibiotics is prescribed as immediate therapy, then the first dose will be directly observed by a member of the study team.

##### 9.4.2 Challenge participants

All challenge participants will receive immediate antibiotic eradication therapy at the C28 visit (or the withdrawal visit if withdrawing from the study) regardless of their colonisation status.

##### 9.4.3 Contact participants

Contact participants will be assessed for transmission of the GM-Nlac strains at the C28 visit (or the withdrawal visit if withdrawing from the study). Delayed antibiotic therapy will be administered to take home. A study team member will contact them when their colonisation status is known (24-72 hours following the visit) and they will be instructed to take the antibiotics if they are colonised, or to dispose of them if they are not colonised.

Contact participants may also be assessed for transmission during the course of the study if they attend an additional visit for safety concerns. If a contact participant was found to be colonised at an additional visit but did not receive antibiotics at that point (e.g. if symptoms have resolved prior to culture results being available), then they will be administered immediate antibiotics to be taken at the C28 / withdrawal visit.

##### 9.4.4 Antibiotic therapy due to safety concerns – challenge and contact participants

In the event of safety concerns arising, antibiotic therapy may be commenced during the course of the study as discussed in section 8.5.2.

##### 9.4.5 Post-eradication visit (PE)

All challenge participants, and all contact participants who have taken antibiotic therapy to clear colonisation, will have a further visit 1-4 days following completion of antibiotic therapy. Safety parameters will be checked and a throat swab (challenge and contact participants) and nasal wash (challenge participants only) will be taken to confirm clearance of GM-Nlac colonisation.

In the event that any GM-Nlac colonies are isolated from a PE throat swab or nasal wash, the participant will be invited to a further visit for an additional throat swab +/- nasal wash. If this remains positive, then further antibiotic administration will be considered on a case-by-case basis by the study steering committee with consultation with DSMB members where necessary.

#### 9.5 Management of safety concerns

##### 9.5.1 Consideration of possible GM-Nlac disease

As discussed in section 3.4, the GM-Nlac strains and non-capsulate and are not considered to pose any greater risk of pathogenesis in humans than the wild-type organism, i.e. they are not anticipated to cause any symptoms or disease. However, participants will be closely monitored, and any safety concerns will be assessed with consideration of possible relatedness to GM-Nlac disease.

Safety data will be collected from participants at each post-inoculation study visit, and at additional visits if any symptoms suggestive of respiratory or systemic infection develop between visits (8.3.6). Solicited and unsolicited adverse events and any abnormalities detected on measurement of vital signs, laboratory tests or physical examination will be assessed by a study doctor with consideration of potential alternative causes (e.g. hayfever, intercurrent viral illnesses) and the possibility of GM-Nlac disease. Further clinically relevant investigations may be conducted as part of this assessment. In the event of significant symptoms developing which are felt to be potentially attributable to GM-Nlac disease, then treatment with appropriate oral or parenteral antibiotics will be commenced following discussion with the Principal or Chief Investigator.

##### 9.5.2 Management of potential GM-Nlac disease

Participants with symptoms which are considered to be potentially attributable to GM-Nlac disease may receive a single dose of oral ciprofloxacin as antibiotic eradication therapy or may be commenced on a treatment course of effective oral antibiotics (e.g. penicillin/amoxicillin or ciprofloxacin). This decision will be made by the Chief Investigator and/or Principal Investigator taking into consideration the severity and duration of symptoms and the results of relevant clinical tests.

In the event of any suspicion of GM-Nlac bacteraemia or sepsis the participant will be referred to the acute medical services who will administer ceftriaxone and manage in a similar way to meningococcal disease.

In the unlikely event of participants experiencing severe illness, a 24/7 resuscitation service is available in the NIHR Clinical Research Facility, which is considered a hospital ward for resuscitation purposes, and which is located in the centre of the University Hospital of Southampton. A 24/7 intensive care service is also available within the University Hospital of Southampton NHS Foundation Trust.

##### 9.5.3 Study holding criteria

Safety parameters will be monitored closely throughout the study. The following criteria will trigger a study pause:

1. A clinical diagnosis of a respiratory or systemic infection requiring a treatment course of oral or parenteral antibiotics, assessed as definitely or probably related to GM-Nlac inoculation, with reference to section 0
2. Early antibiotic eradication therapy – antibiotic eradication therapy (i.e. a single dose of oral ciprofloxacin) given due to safety concerns in 2 of the first 5 participants, or 3+ participants overall

In the event of a study holding criterion being met the study will be paused for a safety review and the sponsor and DSMB will be informed. No further participants will be challenged until the data have been reviewed by the DSMB and study continuation approved. Information will be passed to participants’ GPs, Hampshire and Isle of Wight Health Protection Team South-East and UKHSA as appropriate.

#### 9.6 Withdrawal of participants

##### 9.6.1 Reasons for withdrawal

In accordance with the principles of the current revision of the Declaration of Helsinki (updated 2024) and any other applicable regulations, a participant has the right to withdraw from the study at any time and for any reason and is not obliged to give his reasons for doing so. In addition, a participant may withdraw/be withdrawn from further study procedures at any time in the interests of the participant’s health and well-being, or for any of the following reasons:

- Non-compliance with study requirements or infection control guidelines (*)
- Ineligibility (either arising during the study or retrospectively, having been overlooked at screening) including withdrawal of their bedroom sharer as a contact / challenge participant
- An adverse event (AE), which requires discontinuation of the study involvement or results in inability to continue to comply with study procedures
- Significant protocol deviation
- Administrative decision by the Investigator

The reason for withdrawal from further study procedures will be recorded in the Case Report Form (CRF).

##### 9.6.2 Non-compliance with study requirements

If a participant does not follow the instructions of the clinical team and is therefore compromising their own safety and that of the staff or environment, the challenge will be stopped and antibiotic eradication therapy will be given. This will be reported as an AE.

##### 9.6.3 Withdrawal of corresponding challenge / contact participant

If a challenge participant withdraws from the study, then their corresponding contact participant (if applicable) will also be withdrawn.

If a contact participant withdraws from the study, then their corresponding challenge participant will become ineligible if they have any continued “high risk of transmission activities” (*), e.g. bedroom sharing, with the withdrawn contact participant. The challenge participant will therefore be withdrawn unless both explicitly agree to cease bedroom sharing and other high-risk activities for the remainder of the study involvement, with the agreement of the Principal and/or Chief Investigator.

##### 9.6.4 Procedures for withdrawal

Withdrawing challenge and contact participants will be asked to attend a withdrawal visit. This visit will be conducted according to the C28 schedule (Table 4: Schedule of procedures – The GM Nlac Study – Pilot: Challenge participantsTable 4 and 5), with a subsequent PE visit following antibiotic eradication where applicable (8.4.5). Infection control guidelines (Table 6) should be adhered to until clearance of GM-Nlac colonisation has been confirmed.

Except in case of complete consent withdrawal, safety data collection will be continued. Samples collected prior to withdrawal will be used/stored unless the participant specifically requests otherwise. Data from participants withdrawn from the study will be included in the analysis of results relating to the study’s primary objective.

For all AEs, appropriate follow-up visits or medical care will be arranged, with the agreement of the participant, until the AE has resolved, stabilised or a non-study related causality has been assigned. Any participant who withdrew consent or is withdrawn from further study procedures may be replaced.

If participants withdraw from the study prior to its completion, they will be offered financial re-imbursement corresponding to the number of visits attended.

##### 9.6.5 Missing participant

In the unlikely event that a participant goes missing and/or is uncontactable by telephone, text or email after the challenge, the following stakeholders will be informed;

- The Chief and Principal Investigator
- The participant’s next of kin and/or corresponding contact/challenge participant if applicable
- The participant’s General Practitioner
- The study sponsor
- The hospital trust R&D department

### 10 Containment of GM-Nlac

#### 10.1 Potential for onward transmission of GM-Nlac

The GM-Nlac strains are not anticipated to have any greater potential for transmission or pathogenicity than wild-type Nlac. Nlac colonisation is restricted to humans, with the greatest risk of transmission being to those who share a bedroom with colonised individuals. In our previous GM *N. lactamica* study (57) we did not detect shedding from colonised participants, nor transmission to their bedroom sharers so we anticipate that the risk of onward transmission is low. However, every effort will be made to mitigate the small risk of onward transmission, particularly to any potentially more vulnerable individuals, or to young children who have increased potential for colonisation and onward transmission of wild-type Nlac (20, 41). Relevant mitigation strategies are detailed below.

#### 10.2 Informing stakeholders

DEFRA have been given full information regarding the four GM-Nlac strains contained within 4xrNlac, the results of pre-clinical testing, the planned studies and infection control procedures. Following consideration of this information, approval has been given for the deliberate release of these four strains as described in this protocol (DEFRA REF 25/R50/01).

Hampshire and Isle of Wight Health Protection Team South-East will be informed when the study commences. Participants’ GPs will be notified about their inclusion in these studies. Any unexpected occurrence of disease in participants will be notified to the DSMB. If the DSMB considers the event to be causally related to the GM, the study will be paused or stopped according to the rules in section 8.5.3, and this information will be passed on to UKHSA and the participant’s GP.

#### 10.3  Participant requirements

##### 10.3.1 Contact participants

The highest risk of transmission of GM-Nlac will be to individuals sharing a bedroom with challenge participants during the study period. Therefore, if potential challenge participants anticipate sharing a bedroom with any individual during the study period then they will be invited to participate as a contact participant, with a maximum of one contact participant per challenge participant. Potential contact participants will give informed consent and undergo a medical screening to ensure eligibility prior to inoculation of the challenge participant. If any potential bedroom sharers are not willing or eligible to participate as contact participants, then the challenge participant will not be eligible for the study.

Contact participants will be screened for transmission of the GMOs by a throat swab taken at the end of the study, at an additional visit if symptoms suggestive of respiratory or systemic infection occur, or if they, or their corresponding challenge participant, withdraw prior to the end of the study. Any contact participants found to be colonised will be given antibiotic therapy to clear colonisation.

##### 10.3.2 Eligibility criteria

Challenge and contact participants will be enrolled according to specific inclusion and exclusion criteria to ensure that they are not at any increased risk of disease or harm from colonisation with the GM-*N. lactamica* strains or any other study procedures such as antibiotic administration, and that the potential risk of onward transmission to other humans, particularly to immunocompromised individuals, is minimised. The eligibility criteria are detailed in section 7.6.

##### 10.3.3 Infection control rules

Both challenge and contact participants will be required to adhere to infection control rules for the duration of their involvement in the study (Table 6). These will be explained in a written participant information sheet which participants will receive prior to screening and they will be verbally explained by a study doctor at the screening appointment. Understanding of these rules will be confirmed as part of the pre-consent questionnaire, and agreement to abide by these rules will be included in the written informed consent form.

#### 10.4 Laboratory infection control procedures

GM-*N. lactamica* is routinely handled in University of Southampton laboratories under Health & Safety Executive notification GM57/14.3. The GM-*N. lactamica* stock vials, inoculum and all participant samples potentially contaminated with the GM strains will be processed according to study specific SOPs within category 2 microbiological safety cabinets by trained personnel wearing appropriate personal protective equipment (disposable gloves and laboratory coat).

All waste potentially contaminated with the GMOs will be treated according to site standard operating procedures (SOPs) for disposing of GM contaminated waste. Briefly: materials used during the preparation of the inoculum will be decontaminated overnight in efficacious biocide (e.g. an at-least 2 % (w/v) Virkon solution or an at- least 5 % Biocleanse solution). All potentially GM-contaminated materials will be disposed of into a clinical waste bag or sharps bin which will be placed into an autoclave box. All waste will then be inactivated by autoclaving prior to disposal and removal from the site. All associated procedures have been validated, site autoclaves are accredited annually with contracts in place for regular equipment servicing and maintenance.

#### 10.5 Clinical infection control procedures

The inoculation procedure and any clinical sampling to investigate the colonisation of the GMOs by the participant (throat swabs, nasal washes, nasal swabs) will be carried out in one of the containment level 2 environmental chambers within the NIHR-CRF to assure the inoculum will be administered to the participant only, without posing any risk of infection to other people or the environment.

The study team will wear disposable gloves, apron and a surgical mask during the inoculation. All used material will be disposed of in clinical waste bags and processed following NHS guidelines. Any material potentially contaminated with one of the GMOs will be disposed of according to the Standard operating procedure **SCBR/GEN/099 Disposing of GM Waste**.

The chamber will be cleaned according to hospital guidelines, specifically the glass windows and inside walls of the hood will be cleaned with 70 % (v/v) alcohol solution (e.g. ethanol or industrial methylated spirits) after each participant visit, and the environmental suite will be cleaned with actichlor at the end of each day of participant visits.

### 11 Laboratory procedures

#### 11.1 Screening and safety clinical samples

Screening and safety samples will include blood for routine haematological (full blood count) and biochemical analysis (renal and liver profile, C-reactive protein), urine toxicology screening and respiratory swabs for viral PCR. These samples will be labelled with the participant’s details and hospital number and sent directly to the University Hospital Southampton clinical laboratory for routine processing.

#### 11.2 Research clinical samples

Clinical samples taken for analysis of research endpoints will include respiratory samples for microbiological analysis (throat swabs and nasal washes), mucosal samples for immunological analysis (nasosorption and saliva samples) and blood samples for immunological analysis. These samples will be clearly labelled with the study code, participant’s unique anonymised identifier, sample ID and time point, and recorded in the case report form and the appropriate laboratory source document. Research samples and accompanying documentation will not be labelled with any personal identifiable information.

#### 11.3 Research sample processing

Research clinical samples may be processed in the NIHR-CRF or UoS laboratories or at the laboratories of the co-investigator team. Standard operating procedures will be followed for all laboratory work and will be recorded on the appropriate laboratory source document. Investigators will be trained in these procedures and will conform to established laboratory safety standards.

#### 11.4 Microbiological analyses

Colonisation will be assessed by microbiological analysis of throat swabs and nasal wash samples. Samples will be plated on X-gal-supplemented GC agar plates. Putative Nlac colonies will be identified on the basis of colony morphology and colouration in the first instance, being the only *Neisseria* species to grow as blue colonies on these plates. Biochemical testing (API-NH, bioMérieaux) can be used to confirm the identity of non-blue colonies other *Neisseria* spp. e.g. Nmen.

Total genomic DNA will be extracted from the remaining volume of each sample eluate and analysed using a bespoke qPCR assay for each of the four strains contained within 4xrNlac. This will confirm the presence and total genome count of each strain, allowing determination of the relative density of the resident 4xrNlac population. Comparison will be made between the nasal wash and throat swab samples to assess GM-Nlac colonisation density using qPCR.

Other potentially relevant microbiological assays may be performed on samples at the discretion of the investigator.

#### 11.5 Immunological analyses

Following initial processing, all samples will be stored at -80ºC or liquid nitrogen as appropriate, in secured and temperature-controlled facilities within the NIHR-CRF or UoS laboratories. Samples transported for analysis at the laboratories of co-investigator team members will be transported under secure temperature monitored conditions with appropriate contracts and approvals in place.

The immunological response to colonisation will be assessed by analysis of samples at baseline in comparison to post-inoculation timepoints. Blood samples will be used to assess meningococcal serum bactericidal activity and serum IgG specific for each FHbp variant and NadA. Respiratory samples will be used to assess mucosal IgA specific for each FHbp variant and NadA.

Additionally, clinical samples will be used to develop and optimise cellular immunological assays to determine T and B cell responses to GM-Nlac colonisation. Other potentially relevant immunological assays may be performed on samples at the discretion of the investigator, including the use of sequencing based-technology platforms such as single cell RNA sequencing and T-cell receptor (TCR) sequencing.

### 12 Safety monitoring

#### 12.1 Adverse event definitions

| Adverse Event (AE) | Any untoward medical occurrence in a participant, including a dosing error, which may occur during or after administration of the inoculum and does not necessarily have a causal relationship with the intervention. An AE can therefore be any unfavourable and unintended sign (including an abnormal laboratory finding), symptom or disease temporally associated with the study intervention, whether or not considered related to the study intervention. |
| --- | --- |
| Adverse Reaction (AR) | Any untoward or unintended response to the inoculum. This means that a causal relationship between the inoculum and an AE is at least a reasonable possibility, i.e., the relationship cannot be ruled out. All cases judged by either the reporting medical investigator or the sponsors as having a reasonable suspected causal relationship to the inoculum (i.e. possibly, probably or definitely related to the inoculum) will qualify as adverse reactions. |
| Unexpected Adverse Reaction (UAR) | An adverse reaction, the nature or severity of which is not consistent with the applicable information about the inoculum in the protocol, is considered as an unexpected adverse reaction. |
| Adverse Event of Special Interest (AESI) | An adverse event of special interest (serious or non-serious) is one of scientific and medical concern specific to the inoculum, for which ongoing monitoring and rapid communication by the investigator to the Sponsor can be appropriate. Such an event might warrant further investigation in order to characterise and understand it. Depending on the nature of the event, rapid communication by the Sponsor to other parties (e.g., regulators, DSMB) might also be warranted. |

| Serious Adverse Event (SAE) | An SAE is an AE that results in any of the following outcomes, whether or not considered related to the study intervention.   - Death (i.e., results in death from any cause at any time) - Life-threatening event (i.e., the participant was, in the view of the investigator, at immediate risk of death from the event that occurred). This does not include an AE that, if it occurred in a more serious form, might have caused death. - Persistent or significant disability or incapacity (i.e. substantial disruption of one’s ability to carry out normal life functions). - Hospitalisation other than admission in the NIHR-CRF, regardless of length of stay, even if it is a precautionary measure for continued observation. Hospitalisation (including inpatient or outpatient hospitalisation for an elective procedure) for a pre-existing condition that has not worsened unexpectedly does not constitute a serious AE. - An important medical event (that may not cause death, be life threatening, or require hospitalisation) that may, based upon appropriate medical judgment, jeopardise the participant and/or require medical or surgical intervention to prevent one of the outcomes listed above. Examples of such medical events include allergic reaction requiring intensive treatment in an emergency department or clinic, blood dyscrasias, or convulsions that do not result in inpatient hospitalisation. - Congenital anomaly or birth defect. |
| --- | --- |
| Serious Adverse Reaction (SAR) | An adverse event (expected or unexpected) that is both serious and, in the opinion of the reporting investigator or sponsors, believed to be possibly, probably or definitely due to the inoculum or any other study treatments, based on the information provided in the protocol. |
| Suspected Unexpected Serious Adverse Reaction (SUSAR) | An SAE that is unexpected and thought to be possibly, probably or definitely related to the inoculum. |

Table 8: Adverse event definitions

#### 12.2 Severity assessment

##### 12.2.1 Solicited and unsolicited clinical adverse events

Participants will be asked to report solicited symptoms (Table 7) and unsolicited adverse events (any other symptoms or adverse events) at each study visit. These will be graded by the study team in discussion with participants with reference to the scale in Table 9:

| **Grade 0** | None |
| --- | --- |
| **Grade 1** | Mild: Transient or mild discomfort (< 48 hours); no interference with activity, no medical intervention/therapy required |
| **Grade 2** | Moderate: Mild to moderate limitation in activity - some assistance may be needed; no or minimal medical intervention/therapy required |
| **Grade 3** | Severe: Marked limitation in activity, some assistance usually required; medical intervention/therapy required |

Table 9: Severity grading for clinical adverse events

##### 12.2.2 Vital signs

Participants will have vital signs measured at each study visit and if required in the event of a safety concern. The clinical significance of any out-of-range vital signs will be considered by the investigator and if deemed clinically significant, they will be graded according to Table 10.

##### 12.2.3 Laboratory parameters

Participants will have safety bloods taken as per protocol and additionally if required in the event of a safety concern. The clinical significance of any out-of-range laboratory parameters will be considered by the investigator and if deemed clinically significant, they will be graded according to Table 11.

| **Vital signs*** | **Grade 1**  **(mild)** | **Grade 2 (moderate)** | **Grade 3**  **(severe)** |
| --- | --- | --- | --- |
| Temperature (°C) ** | 38.0 – 38.4 | 38.5 – 38.9 | ≥ 39 |
| Tachycardia (beats per minute) | 101 – 115 | 116 – 130 | >130 |
| Bradycardia (beats per minute)*** | 45 – 50 | 40 – 45 | < 40 |
| Hypertension (systolic, mmHg) | 141 – 169 | 170 – 179 | > 180 |
| Hypertension (diastolic, mmHg) | 91 – 95 | 96 – 100 | >100 |
| Hypotension (systolic, mmHg) | 85 – 89 | 80 – 84 | < 80 |
| Oxygen Saturation (%) | 92 – 94 | 90-91 | < 90 |
| RR (breaths per minute | 21 – 22 | 23 – 25 | > 25 |

Table 10: Vital sign adverse event grading

Based on FDA toxicity grading guidelines with adaptation according to UHS/RCP NEWS normal ranges

*Participant should be at rest for all vital sign measurements

**Oral temperature; no recent hot or cold beverages or smoking

***When resting heart rate is between 60-100 beats per minute. Use clinical judgement when characterising bradycardia among some healthy populations, for example, conditioned athletes

| **Adverse event** | | **Normal range*** | **Grade 1 (mild)** | **Grade 2 (moderate)** | **Grade 3 (severe)** |
| --- | --- | --- | --- | --- | --- |
| **Haematology** | | | | | |
| Haemoglobin  (g/L) | Female | 120 – 150 | 105 – 113 | 90 – 104 | < 90 |
|  | Male | 130 – 170 | 115 – 125 | 100 – 114 | < 100 |
| White cell count  (x 10^9^/L) | High | 4.00 – 10.00 | 11.00 – 15.00 | 15.01 – 20.00 | > 20.00 |
|  | Low | 4.00 – 10.00 | 2.50 – 3.50 | 1.5 – 2.49 | < 1.50 |
| Neutrophil count (x 10^9^/L) | | 2.00 – 7.00 | 1.50 – 1.90 | 1.00 – 1.49 | < 1.00 |
| Lymphocyte count (x 10^9^/L) | | 1.50 – 4.00 | 0.75 – 1.00 | 0.50 – 0.74 | < 0.50 |
| Platelet count (x 10^9^/L) | | 150 – 400 | 125 – 140 | 100 – 124 | < 100 |
| **Biochemistry** | | | | | |
| C reactive protein (mg/l) | | 0 – 5 | 10 – 19 | 20 – 50 | > 50 |
| Sodium (mmol/L) | Low | 133 – 146 | 131 – 132 | 130 | < 130 |
|  | High | 133 – 146 | 147 | 148 | > 148 |
| Potassium (mmol/L) | Low | 3.5 – 5.3 | 3.3 – 3.4 | 3.1 – 3.2 | < 3.1 |
|  | High | 3.5 – 5.3 | 5.4 – 5.5 | 5.6 – 5.7 | > 5.7 |
| Urea (mmol/L) | | 2.5 – 7.8 | 8.2 – 9.3 | 9.4 – 11.0 | > 11.0 |
| Creatinine (µmol/L) | Female | 45 – 84 | 132 – 150 | 151 – 177 | > 177 |
|  | Male | 59 – 104 | 132 – 150 | 151 – 177 | > 177 |
| ALT (IU/L) | Female | 0 – 34 | 38 – 88 | 89 – 170 | > 170 |
|  | Male | 0 – 49 | 54 – 127 | 128 – 245 | > 245 |
| ALP (IU/L) | | 30 – 130 | 143 – 272 | 273 – 390 | > 390 |
| Bilirubin (µmol/L) | With LFT increase | 0 – 20 | 22 – 25 | 25.1 – 3 | > 30 |
|  | With normal LFTs | 0 – 20 | 22 – 32 | 32.1 – 40 | > 40 |

Table 11: Laboratory parameter adverse event grading

Based on FDA toxicity grading guidelines with adaptation according to UHS normal ranges

*University Hospital Southampton Clinical Laboratory reference range

#### 12.3 Causality assessment

For each AE, an assessment of the relationship of the AE to the study intervention (GM-Nlac challenge) or other study procedure will be undertaken. The relationship of the AE to GM-Nlac challenge or other study procedure will be categorised as unrelated, unlikely to be related, possibly related, probably related or definitely related (Table 12). A related AE refers to an AE for which there is a possible, probable or definite relationship to the GM-Nlac challenge or study procedure. The investigator will use clinical judgment to determine the relationship. Alternative causes of the AE, such as the natural history of pre-existing medical conditions, concomitant therapy, other risk factors and the temporal relationship of the event to the challenge will be considered and investigated.

| **No Relationship** | No temporal relationship to GM-Nlac challenge or study procedure; ***and***  Alternate aetiology (clinical state, environmental or other interventions); ***and***  Does not follow expected pattern of response to GM *N. lactamica challenge* or study procedure |
| --- | --- |
| **Unlikely** | Unlikely temporal relationship to GM-Nlac challenge or study procedure; ***and***  Alternate aetiology likely (clinical state, environmental or other interventions); ***and***  Does not follow expected pattern of response to GM-Nlac challenge or study procedure |
| **Possible** | Reasonable temporal relationship to GM-Nlac challenge or study procedure; ***or***  Event not readily produced by clinical state, environmental or other interventions; ***or***  Follows expected pattern of response to GM-Nlac challenge or study procedure |
| **Probable** | Reasonable temporal relationship to GM-Nlac challenge or study procedure; ***and***  Event not readily produced by clinical state, environment, or other interventions ***or***  Follows expected pattern of response to GM-Nlac challenge or study procedure |
| **Definite** | Reasonable temporal relationship to GM-Nlac challenge or study procedure; ***and***  Event not readily produced by clinical state, environment, or other interventions; ***and***  Follows expected pattern of response to GM-Nlac challenge or study procedure |

Table 12: Adverse event causality assessment

#### 12.4 Potential adverse events

##### 12.4.1 Inoculation

The inoculation with GM-Nlac suspension (0.5 mL per nostril) may cause some irritation of the nasal mucosa that will disappear within a few seconds. Very occasionally, instillation may induce coughing or sneezing, but this can be prevented by slow instillation down the superior wall of the nares.

##### 12.4.2 GM-Nlac disease

We have previously inoculated over 450 individuals with wild-type Nlac and 26 with prior GM-Nlac strains 4NB1 and 4YB2 with no AEs suggestive of Nlac or GM-Nlac disease and no significant safety concerns. We do not anticipate the strains contained in 4xrNlac to have any increased capacity for pathogenicity. In the unlikely event of a GM-Nlac strain causing disease we would anticipate that this would be either due to involvement of the respiratory tract (which would be signalled by fever and cough) or due to invasion of the bloodstream (which would be signalled by fever and features of sepsis akin to meningococcal disease). In the event of development any symptoms or laboratory parameter abnormalities suggestive of respiratory or systemic infection, the participant will be reviewed by a study doctor.

##### 12.4.3 Phlebotomy

The maximum volume of blood drawn over the study period (218 mls) should not compromise these healthy participants. There may be minor bruising, local tenderness or pre-syncopal symptoms associated with venepuncture, which will not be documented as AEs if they occur.

##### 12.4.4 Respiratory sampling

The clinical samples taken by throat swab, nasal wash and nasosorption sampling can cause some irritation of the pharyngeal/nasal mucosa and can induce coughing or sneezing. This nasal discomfort will disappear within a few minutes and will not be recorded as an AE. The collection of saliva samples is not expected to cause any discomfort.

##### 12.4.5 Antibiotic eradication therapy

The side effects of ciprofloxacin in young adults may include:

- Gastrointestinal disturbance such as abdominal ache, diarrhoea and nausea
- Tiredness, dizziness and headaches
- Rash and itching
- Facial swelling - very rarely breathing difficulties may occur with facial swelling
- Arthralgia, myalgia and tendon disorders
- Eye and vision disorders
- QT interval prolongation

Participants will be advised to contact the clinical study team if side effects occur.

Female participants using the oral contraceptive pill will be advised to use alternative forms of contraception for two weeks following eradication therapy as ciprofloxacin may interfere with the oral contraceptive pill.

#### 12.5 Data and Safety Monitoring Board

An independent DSMB will be established prior to the start of the study to provide oversight of safety and study conduct.

The DSMB will review the study protocol and safety procedures prior to commencement of the study. Formal interactions with the DSMB will be as per the DSMB terms of reference. In addition to this the DSMB will be contacted for independent review in the following circumstances:

- If a study holding criterion is met (8.5.3)
- If the study steering committee or sponsor have concerns about the safety of a participant or the general public

The outcome of each DSMB review, including recommendations regarding study continuation, will be communicated directly to the study investigators and documentation of all reviews will be kept in the study master file.

The role of the DSMB is to provide independent overview of study conduct and safety and to advise on study continuation. The ultimate decision for the continuation of the study lies with the Chief Investigator.

All correspondence between investigator and DSMB will be conveyed by the investigator to the Sponsor.

#### 12.6 Reporting procedures

##### 12.6.1 Documentation of adverse events

All AEs will be recorded in the participant’s paper CRF. Solicited symptoms with grading will be recorded in the study visit page. An adverse event form will be completed for unsolicited adverse events and vital signs / laboratory parameters deemed to be clinically significant. Adverse events will be summarised in regular safety reports for review by the DSMB.

##### 12.6.2 Reporting procedures for serious AEs (SAEs)

In order to comply with current regulations on serious adverse event reporting to regulatory authorities, the event will be documented accurately and notification deadlines respected. SAEs will be reported to the principal investigator and/or chief investigator immediately when the study team is aware of their occurrence. The sponsor will be notified as soon as possible (within 24 hours) when the investigators are aware of their occurrence. The DSMB will be notified of SAEs deemed possibly, probably or definitely related to GM-Nlac challenge; SAEs will not normally be reported to the ethical committee(s) unless there is a clinically important increase in occurrence rate, an unexpected outcome, or a new event that is likely to affect safety of study participants, at the discretion of the Chief Investigator. In addition to the expedited reporting above, the investigator shall include all SAEs in the annual safety report.

##### 12.6.3 Adverse events of special interest

Adverse events of special interest will be reported as SAEs. These are:

- Severe hypersensitivity reactions to the inoculum (e.g. anaphylaxis)
- Overdosing of the inoculum (>10 x the total intended dose i.e. 4 x 10^6^ CFU in total)
- GM-Nlac disease as defined in section 8.5.3

##### 12.6.4 Reporting procedures for SUSARs

The chief investigator will report all SUSARs to the sponsor within 3 days, and the ethical committee(s) within 15 days of the study team being aware of their occurrence. The chief investigator will also inform all investigators concerned of relevant information about SUSARs that could adversely affect the safety of participants. In addition, the chief investigator will report any SUSARs relating to licensed products used in the trial (ciprofloxacin) to the Medicine and Healthcare products Regulatory Agency (MHRA) using the electronic ‘Yellow Card’ System.

All SUSARs and deaths occurring during the study will be reported to the sponsor. For all deaths, any autopsy reports and relevant medical reports will be made available for reporting to the relevant authorities.

##### 12.6.5 Safety reviews

The first challenge participant will be inoculated individually. Safety data including laboratory parameters will be reviewed by the Principal and/or Chief Investigator by day 6-8 post inoculation. Providing there are no safety concerns, further challenge participants will be inoculated individually or in pairs, with safety reviews at day 6-8 post inoculation, up to a total of 5 challenge participants. The remaining challenge participants will be inoculated in groups of a maximum of five with safety reviews at day 6-8 post inoculation. The decision to inoculate the next group of challenge participants will be taken by the investigators, with discussion with the DSMB if there are any safety concerns.

The safety profile will be assessed on an on-going basis by the study steering committee with discussion with the DSMB where required.

##### 12.6.6 Regular study reports

A safety report will be written after inoculation of the fifth challenge participantand submitted to the DSMB. A further safety report will be written following completion of follow up of the final participant. An annual safety report for the study will be prepared by the anniversary of the first approval date of the study from the regulatory authority. This will be submitted to the sponsor, the DSMB and the HRA.

##### 12.6.7 Procedures to be followed in the event of abnormal findings

Abnormal clinical findings from medical history, examination or blood tests, will be assessed as to their clinical significance. If a test is deemed clinically significant, it may be repeated, to ensure it is not a single occurrence. If a test remains clinically significant, the participant will be informed, and appropriate medical care arranged as appropriate with the permission of the participant. Decisions to exclude the participant from enrolling in the study or to withdraw a participant from the study will be at the discretion of the investigator.

### 13 Statistical analysis

#### 13.1 Sample size

No formal sample size calculation has been performed for this open-label pilot study. The recruitment target is ten challenge participants, as this is considered to be sufficient to allow an estimation of colonisation fraction and to provide initial safety data, and to allow the optimisation and validation of immunological and microbiological assays for use in downstream studies. Non-evaluable challenge participants may be replaced as discussed in section 5.3.

#### 13.2 Statistical analysis

Safety endpoints will be analysed using safety data from all challenge participants who have been inoculated with 4rxNlac, with assessment of solicited and unsolicited adverse events during the follow-up period overall, and with comparison between colonised and non-colonised challenge participants. Safety data will also be analysed for all enrolled contact participants.

Microbiological and immunological endpoints will be analysed using only those challenge participants who are considered to be evaluable (see section 5.3). Transmission to contact participants will be analysed using microbiological data from contact participants for whom the corresponding challenge participant was colonised.

### 14 Quality control and quality assurance

#### 14.1 Study oversight

The study steering committee will include the Lead Scientific Investigator, Chief Investigator, Principal Investigator and key co-investigators as detailed on p3-4. This committee will oversee progress and management of the study.

The DSMB (11.5) will provide independent oversight of the conduct and safety of the study and provide advice to the study steering committee regarding continuation of the study in the event of any safety concerns.

#### 14.2 Study amendments

Any amendments to study documents will follow established HRA and REC requirements. Amendments to the study that appear necessary during the course of the study will be agreed by the study steering committee and sponsor. No amendments to this protocol will be made without consultation with, and agreement of, the Sponsor. If agreement is reached concerning the need for an amendment, it will be produced in writing by the chief investigator and will be made a formal part of the protocol following ethical and regulatory approval (NRES-REC SOPs – Version 7.7 April 2025: <https://www.hra.nhs.uk/about-us/committees-and-services/res-and-recs/research-ethics-committee-standard-operating-procedures/>).

An administrative change to the protocol is one that modifies administrative and logistical aspects of a protocol but does not affect participant safety, the objectives of the study or study progress. An administrative change does not require UK ethical committee or regulatory approval.

The chief investigator is responsible for ensuring that changes to an approved study, during the period for which regulatory and ethical committee(s) approval has already been given, are not initiated without regulatory and ethical committee(s)’ review and approval except to eliminate apparent immediate hazards to a participant.

#### 14.3 Risk assessment

A risk assessment will be prepared and agreed with the Sponsor prior to study commencement. This will be reviewed as necessary over the course of the study to reflect significant changes to the protocol, relevant safety data, or outcomes of monitoring activities.

#### 14.4 Monitoring

A monitoring plan will be prepared and agreed with the Sponsor prior to study commencement. Monitoring will be performed by the Sponsor or delegated representatives according to International Conference on Harmonisation (ICH) Good Clinical Practice (GCP). Following written standard operating procedures, the monitors will verify that the clinical study is conducted and data are generated, documented and reported in compliance with the protocol, GCP and the applicable regulatory requirements. The investigator will provide direct access to all study related source data/documents and reports for the purpose of monitoring and auditing by the sponsor and inspection by local and regulatory authorities.

#### 14.5 Quality Control, Quality Assurance and statutory inspection

The UHS R&D department QA staff will provide Quality Assurance (QA) for the study and perform internal audits to check that the study is being conducted, data recorded, analysed and accurately reported according to the protocol, Sponsor’s SOPs and in compliance with ICH GCP. The audits will also include laboratory activities according to an agreed audit schedule. The internal audits will supplement the sponsor’s monitoring process and will review processes not covered by the sponsor’s monitor.

A Quality control (QC) plan will be established at the start of the study, as per local SOP. The Sponsor, study site and ethical committee may carry out audit to ensure compliance with the protocol, GCP and appropriate regulations. GCP inspections may also be undertaken by the regulatory authority to ensure compliance with protocol and national regulations. The sponsor will assist in any inspections.

#### 14.6 Investigator procedures

Approved standard operating procedures (SOPs) will be used at all clinical and laboratory sites.

#### 14.7 Protocol deviation

Any deviations from the protocol will be documented in a protocol deviation form and filed in the site study master file within the NIHR-CRF.

#### 14.8 Serious breaches

A “serious breach” is defined as a breach of the protocol or, of the principles of Good Clinical Practice which is likely to affect to a significant degree the safety or physical or mental integrity of the research participants, or the scientific value of the research.

In the event that a serious breach is suspected the Sponsor will be informed as soon as possible and in turn will notify the REC and external safety committee within 7 days.

#### 14.9 Study completion/termination

The study may be terminated early at the discretion of the Chief Investigator, Sponsor or DSMB if there are safety concerns, concerns about compliance with GCP or other appropriate regulations, poor recruitment or new information becomes available which has an impact on the scientific validity or safety of the study.

#### 14.10 Exploitation and dissemination

The study steering committee will be involved in writing and reviewing drafts of the manuscripts, abstracts, press releases and any other publications arising from the study. Findings will be published in peer reviewed journals as soon as possible, even where results prove negative. The authors will acknowledge that the study has been funded by the Medical Research Council. The results of the study will be disseminated at relevant international scientific meetings. Participants will be sent a newsletter with a lay summary of the results of the study once the results are available. This will provide information regarding the study in general and not their individual results.

### 15 Regulatory and ethical requirements

#### 15.1 Declaration of Helsinki

The Investigator will ensure that this study is conducted according to the principles of the current revision of the Declaration of Helsinki (2024).

#### 15.2 ICH guidelines for Good Clinical Practice

The Investigator will ensure that this study is conducted in full conformity to the ICH guidelines for GCP (ICH E6(R3) 2025).

#### 15.3 Informed consent

Written informed consent will be gained from all participants following the provision of detailed information about the aims of the study, the level of involvement required, and the risks involved. Participants will be provided with an information sheet prior to the start of the study either in print form or via email. They will be encouraged to use the contact details on this form to contact the study team for further information if necessary. Prior to screening the participants’ understanding of the study and risks involved will be explored including the use of a pre-consent questionnaire, after which they will be asked to sign a consent form.

#### 15.4 Informing participants’ General Practitioners

A letter describing the study and the participant’s involvement will be sent to their General Practitioner (GP) on the day of the screening visit for both challenge and contact participants. This will include contact details for the research team. For challenge participants, the GP will be asked for confirmation of the medical history.

#### 15.5 Research ethics committee

The protocol, informed consent forms, other written participant information and the advertising material will be submitted to the REC and HRA for written approval, using the UK Integrated Research Application System. The investigator will submit and, where necessary, obtain approval from the REC for all subsequent amendments to the protocol and associated study documents. A non-substantial amendment does not require UK ethical committee approval (NRES-REC SOPs – Version 7.7 April 2025: <https://www.hra.nhs.uk/about-us/committees-and-services/res-and-recs/research-ethics-committee-standard-operating-procedures/>). The investigator will notify deviations from the protocol or SAEs occurring at the site to the sponsor and will notify the REC of these if necessary, in accordance with procedures.

#### 15.6 Participant confidentiality

All data will be anonymised; participant data will be identified by a unique study number in paper documentation and electronic files. Files containing identifiable information will be stored in secured locations within the NIHR-CRF. Only the sponsor representative, investigators, the clinical monitor, the ethical committee(s) and the regulatory authorities will have access to the records.

### 16 Data management

#### 16.1 Data handling

The chief investigator will have responsibility for delegating the receiving, entering, cleaning, querying, analysing and storing of all data that accrues from the study. Study team members will enter study data into participant case report form (CRFs) and laboratory source documents (LSDs) which will be in a paper format. This includes safety data, laboratory data (clinical, microbiological and immunological) and outcome data. Link-anonymised data will be transcribed into a study specific database (15.4). All source data, participant CRFs and the electronic files will be stored securely, in compliance with the General Data Protection Regulation (GDPR) 2018.

#### 16.2 Participant identification

Each challenge and contact participant will be allocated a unique link-anonymised participant ID during the pre-screening period. The format of the participant ID will distinguish between challenge and contact participants and will link challenge-contact participant pairs where applicable. This participant ID will be used for all study documentation including case report forms and laboratory source documents and electronic files, and to label clinical samples for analysis in the research laboratories. Personally identifiable information will not be used on study documentation or in electronic files containing study data. No personally identifiable information will be sent to co-investigator laboratories.

Personally identifiable information will be recorded on participant recruitment and reimbursement records, informed consent forms, antibiotic and inoculum prescription / administration records and correspondence with the GP and public health authorities where applicable. These will be stored securely within the NIHR-CRF or in password protected files on the University Hospital Southampton computer system in accordance with GCP, regulatory and sponsor requirements. Clinical samples sent to the hospital clinical laboratory for processing will be labelled with personal details in accordance with hospital laboratory requirements.

#### 16.3 Source documents

Source documents are original documents, data, and records from which the participant’s data are obtained. For this study these will include, but are not limited to; GP records and correspondence and hospital electronic records including laboratory results. CRF entries will be considered source documentation of study data as the site of the original recording. In this study this will include, but is not limited to medical history, medication records, vital signs, physical examination records, urine assessments, blood results, adverse event data and details of study interventions. All source documents, excluding electronic hospital records, will be filed in the NIHR-CRF.

#### 16.4 Study database

Anonymised study data will be transferred to a secure study database (ALEA) by a member of the study team which will be retained in accordance with the University of Southampton’s Research Data Management policy. Transcription will be checked by a separate study team member and the monitor.

Only the Site Investigator and authorised personnel should enter or change data in the electronic Case Report Forms (eCRFs) in ALEA. An audit trail will be incorporated into the eCRFs whereby any changes to the data originally entered will be documented. A table of all changes including the original value, new value, field, relevant visit details, who made the changes and why the changes were made, will be stored in a table in the study database.

Quality of the data entered into the eCRF data fields will be controlled by limited data entry, drop down options and predefined data formats. Range checks for chosen fields will automatically appear where data points are outside of a pre‐specified range. Verification and explanation for the data point will be required and will subsequently appear in the query log for the Data Manager or Study Manager to check.

#### 16.5 Record keeping

The investigators will maintain and retain appropriate medical and research records and essential documents for this study in compliance with ICH E6 GCP and regulatory and institutional requirements for the protection of confidentiality of participants. The chief investigator, co-investigators and clinical research nurses will have access to records. The investigators will permit authorised representatives of the sponsor, regulatory agencies and the monitors to examine (and when required by applicable law, to copy) clinical records for the purposes of quality assurance reviews, audits and evaluation of the study safety and progress. A study site file will be maintained in the NIHR-CRF.

#### 16.6 Data protection

The study protocol, documentation, data and all other information generated will be held in strict confidence, and compliant with the GDPR. No information concerning the study or the data will be released to any unauthorised third party, without prior written approval of the sponsor.

### 17 Financing and insurance

#### 17.1 Financing

The study will be funded by the Medical Research Council (Grant MR/X019284/1 (“Safety and immunogenicity of nasal inoculation with recombinant *Neisseria lactamica* expressing Factor H binding protein and Neisseria Adhesin A”) with additional infrastructure support from the Southampton NIHR-CRF and the Southampton Biomedical Research Centre (BRC).

#### 17.2 Insurance

The University of Southampton has a specialist insurance policy in place, which would operate in the event of any participant suffering harm as a result of their involvement in the research.

#### 17.3 Participant reimbursement

Challenge and contact participants will be re-imbursed for their time, travel and inconvenience as below:

- Attendance at screening and follow-up visits - £75 per visit
- Attendance at pre-challenge and post-eradication check visits - £50 per visit
- Attendance at challenge visit - £125
- Incentive for completion of all study visits and procedures (challenge participants only) - £200
- Travel expenses – up to £25 per visit

Challenge participants will therefore be re-imbursed a maximum of £1000 (including up to £200 travel re-imbursement, plus additional payment for additional visits if required). Contact participants will be reimbursed a maximum of £275 (including up to £75 travel reimbursement, plus additional payment for additional visits if required).

If a participant withdraws or is withdrawn from the study prior to its completion, they will be offered financial re-imbursement corresponding to the number of visits attended.

### 19 References

1.	Gold R, Goldschneider I, Lepow ML, Draper TF, Randolph M. Carriage of Neisseria meningitidis and Neisseria lactamica in infants and children. J Infect Dis. 1978;137(2):112-21.

2.	Pace D, Pollard AJ. Meningococcal disease: clinical presentation and sequelae. Vaccine. 2012;30 Suppl 2:B3-9.

3.	Agier L, Martiny N, Thiongane O, Mueller JE, Paireau J, Watkins ER, et al. Towards understanding the epidemiology of Neisseria meningitidis in the African meningitis belt: a multi-disciplinary overview. Int J Infect Dis. 2017;54:103-12.

4.	GBD Meningitis Collaborators. Global, regional, and national burden of meningitis, 1990-2016: a systematic analysis for the Global Burden of Disease Study 2016. Lancet Neurol. 2018;17(12):1061-82.

5.	WHO. Defeating meningitis by 2030: a global road map. Geneva; 2021.

6.	Walsh L, Clark SA, Derrick JP, Borrow R. Beyond the usual suspects: Reviewing infections caused by typically-commensal Neisseria species. J Infect. 2023;87(6):479-89.

7.	Everts RJ, Speers D, George ST, Ansell BJ, Karunajeewa H, Ramos RD. Neisseria lactamica arthritis and septicemia complicating myeloma. J Clin Microbiol. 2010;48(6):2318.

8.	Wang CY, Chuang YM, Teng LJ, Lee LN, Yang PC, Kuo SH, Hsueh PR. Bacteraemic pneumonia caused by Neisseria lactamica with reduced susceptibility to penicillin and ciprofloxacin in an adult with liver cirrhosis. J Med Microbiol. 2006;55(Pt 8):1151-2.

9.	Zavascki AP, Fritscher L, Superti S, Dias C, Kroth L, Traesel MA, et al. First case report of Neisseria lactamica causing cavitary lung disease in an adult organ transplant recipient. J Clin Microbiol. 2006;44(7):2666-8.

10.	Orden B, Amerigo MA. Acute otitis media caused by Neisseria lactamica. Eur J Clin Microbiol Infect Dis. 1991;10(11):986-7.

11.	Schifman RB, Ryan KJ. Neisseria lactamica septicemia in an immunocompromised patient. J Clin Microbiol. 1983;17(5):934-5.

12.	Wilson HD, Overman TL. Septicemia due to Neisseria lactamica. J Clin Microbiol. 1976;4(3):214-5.

13.	Lauer BA, Fisher CE. Neisseria lactamica meningitis. Am J Dis Child. 1976;130(2):198-9.

14.	Brown NM, Ragge NK, Speller DC. Septicaemia due to Neisseria lactamica--initial confusion with Neisseria meningitidis. J Infect. 1987;15(3):243-5.

15.	Denning DW, Gill SS. Neisseria lactamica meningitis following skull trauma. Rev Infect Dis. 1991;13(2):216-8.

16.	Hansman D. Meningitis caused by Neisseria lactamica. N Engl J Med. 1978;299(9):491.

17.	Changal KH, Raina A, Altaf SS. Neisseria lactamica Causing a Lung Cavity and Skin Rash in a Renal Transplant Patient: First Report from India. Case Rep Infect Dis. 2016;2016:1932963.

18.	Pandey AK, Cleary DW, Laver JR, Maiden MCJ, Didelot X, Gorringe A, Read RC. Neisseria lactamica Y92-1009 complete genome sequence. Stand Genomic Sci. 2017;12:41.

19.	Rouphael NG, Stephens DS. Neisseria meningitidis: biology, microbiology, and epidemiology. Methods Mol Biol. 2012;799:1-20.

20.	Bennett JS, Griffiths DT, McCarthy ND, Sleeman KL, Jolley KA, Crook DW, Maiden MC. Genetic diversity and carriage dynamics of Neisseria lactamica in infants. Infect Immun. 2005;73(4):2424-32.

21.	Cartwright KA, Stuart JM, Jones DM, Noah ND. The Stonehouse survey: nasopharyngeal carriage of meningococci and Neisseria lactamica. Epidemiol Infect. 1987;99(3):591-601.

22.	MacLennan J, Kafatos G, Neal K, Andrews N, Cameron JC, Roberts R, et al. Social behavior and meningococcal carriage in British teenagers. Emerg Infect Dis. 2006;12(6):950-7.

23.	Laver JR, Hughes SE, Read RC. Neisserial Molecular Adaptations to the Nasopharyngeal Niche. Adv Microb Physiol. 2015;66:323-55.

24.	Trotter CL, Gay NJ, Edmunds WJ. The natural history of meningococcal carriage and disease. Epidemiol Infect. 2006;134(3):556-66.

25.	Dale AP, Theodosiou AA, Gbesemete DF, Guy JM, Jones EF, Hill AR, et al. Effect of colonisation with Neisseria lactamica on cross-reactive anti-meningococcal B-cell responses: a randomised, controlled, human infection trial. Lancet Microbe. 2022;3(12):e931-e43.

26.	Evans CM, Pratt CB, Matheson M, Vaughan TE, Findlow J, Borrow R, et al. Nasopharyngeal colonization by Neisseria lactamica and induction of protective immunity against Neisseria meningitidis. Clin Infect Dis. 2011;52(1):70-7.

27.	Simmons G, Martin D, Stewart J, Bremner D. Carriage of N. lactamica in a population at high risk of meningococcal disease. Epidemiol Infect. 2000;125(1):99-104.

28.	Cleary PR, Calvert N, Gee S, Graham C, Gray S, Kaczmarski E, et al. Variations in Neisseria meningitidis carriage by socioeconomic status: a cross-sectional study. J Public Health (Oxf). 2016;38(1):61-70.

29.	Olsen SF, Djurhuus B, Rasmussen K, Joensen HD, Larsen SO, Zoffman H, Lind I. Pharyngeal carriage of Neisseria meningitidis and Neisseria lactamica in households with infants within areas with high and low incidences of meningococcal disease. Epidemiol Infect. 1991;106(3):445-57.

30.	Coen PG, Cartwright K, Stuart J. Mathematical modelling of infection and disease due to Neisseria meningitidis and Neisseria lactamica. Int J Epidemiol. 2000;29(1):180-8.

31.	Maiden MC, Ibarz-Pavon AB, Urwin R, Gray SJ, Andrews NJ, Clarke SC, et al. Impact of meningococcal serogroup C conjugate vaccines on carriage and herd immunity. J Infect Dis. 2008;197(5):737-43.

32.	Kristiansen PA, Ba AK, Ouedraogo AS, Sanou I, Ouedraogo R, Sangare L, et al. Persistent low carriage of serogroup A Neisseria meningitidis two years after mass vaccination with the meningococcal conjugate vaccine, MenAfriVac. BMC Infect Dis. 2014;14:663.

33.	Bolgiano B, Moran E, Beresford NJ, Gao F, Care R, Desai T, et al. Evaluation of Critical Quality Attributes of a Pentavalent (A, C, Y, W, X) Meningococcal Conjugate Vaccine for Global Use. Pathogens. 2021;10(8).

34.	Read RC, Baxter D, Chadwick DR, Faust SN, Finn A, Gordon SB, et al. Effect of a quadrivalent meningococcal ACWY glycoconjugate or a serogroup B meningococcal vaccine on meningococcal carriage: an observer-blind, phase 3 randomised clinical trial. Lancet. 2014;384(9960):2123-31.

35.	Marshall HS, McMillan M, Koehler AP, Lawrence A, Sullivan TR, MacLennan JM, et al. Meningococcal B Vaccine and Meningococcal Carriage in Adolescents in Australia. N Engl J Med. 2020;382(4):318-27.

36.	McMillan M, Koehler AP, Lawrence A, Sullivan TR, Bednarz J, MacLennan JM, et al. B Part of It School Leaver Study: A Repeat Cross-Sectional Study to Assess the Impact of Increasing Coverage With Meningococcal B (4CMenB) Vaccine on Carriage of Neisseria meningitidis. J Infect Dis. 2022;225(4):637-49.

37.	Cartwright KA, Stuart JM, Robinson PM. Meningococcal carriage in close contacts of cases. Epidemiol Infect. 1991;106(1):133-41.

38.	Kristiansen BE, Tveten Y, Jenkins A. Which contacts of patients with meningococcal disease carry the pathogenic strain of Neisseria meningitidis? A population based study. BMJ. 1998;317(7159):621-5.

39.	Wilder-Smith A, Barkham TM, Ravindran S, Earnest A, Paton NI. Persistence of W135 Neisseria meningitidis carriage in returning Hajj pilgrims: risk for early and late transmission to household contacts. Emerg Infect Dis. 2003;9(1):123-6.

40.	Simmons G, Martin D, Stewart J, Bremner D. Carriage of N. lactamica in a population at high risk of meningococcal disease. Epidemiology and infection. 2000;125(1):99-104.

41.	Theodosiou AA, Bogaert D, Cleary DW, Dale AP, Gbesemete DF, Guy JM, et al. Controlled human infection model of Neisseria lactamica in late pregnancy investigating mother-to-infant transmission in the UK: a single-arm pilot trial. Lancet Microbe. 2025:100986.

42.	Fraser A, Gafter-Gvili A, Paul M, Leibovici L. Antibiotics for preventing meningococcal infections. Cochrane Database Syst Rev. 2006(4):CD004785.

43.	Gaunt PN, Lambert BE. Single dose ciprofloxacin for the eradication of pharyngeal carriage of Neisseria meningitidis. J Antimicrob Chemother. 1988;21(4):489-96.

44.	Pugsley MP, Dworzack DL, Horowitz EA, Cuevas TA, Sanders WE, Jr., Sanders CC. Efficacy of ciprofloxacin in the treatment of nasopharyngeal carriers of Neisseria meningitidis. J Infect Dis. 1987;156(1):211-3.

45.	Deasy AM, Guccione E, Dale AP, Andrews N, Evans CM, Bennett JS, et al. Nasal Inoculation of the Commensal Neisseria lactamica Inhibits Carriage of Neisseria meningitidis by Young Adults: A Controlled Human Infection Study. Clin Infect Dis. 2015;60(10):1512-20.

46.	Gbesemete D. Controlled human infection of healthy adults with lyophilized Neisseria lactamica induces asymptomatic, immunogenic nasopharyngeal carriage in the UK and Mali. Under review by OFID.

47.	Vogel U, Taha MK, Vazquez JA, Findlow J, Claus H, Stefanelli P, et al. Predicted strain coverage of a meningococcal multicomponent vaccine (4CMenB) in Europe: a qualitative and quantitative assessment. Lancet Infect Dis. 2013;13(5):416-25.

48.	Capecchi B, Adu-Bobie J, Di Marcello F, Ciucchi L, Masignani V, Taddei A, et al. Neisseria meningitidis NadA is a new invasin which promotes bacterial adhesion to and penetration into human epithelial cells. Mol Microbiol. 2005;55(3):687-98.

49.	Metruccio MM, Pigozzi E, Roncarati D, Berlanda Scorza F, Norais N, Hill SA, et al. A novel phase variation mechanism in the meningococcus driven by a ligand-responsive repressor and differential spacing of distal promoter elements. PLoS Pathog. 2009;5(12):e1000710.

50.	Alamro M, Bidmos FA, Chan H, Oldfield NJ, Newton E, Bai X, et al. Phase variation mediates reductions in expression of surface proteins during persistent meningococcal carriage. Infect Immun. 2014;82(6):2472-84.

51.	Johswich KO, McCaw SE, Islam E, Sintsova A, Gu A, Shively JE, Gray-Owen SD. In vivo adaptation and persistence of Neisseria meningitidis within the nasopharyngeal mucosa. PLoS Pathog. 2013;9(7):e1003509.

52.	Masignani V, Comanducci M, Giuliani MM, Bambini S, Adu-Bobie J, Arico B, et al. Vaccination against Neisseria meningitidis using three variants of the lipoprotein GNA1870. J Exp Med. 2003;197(6):789-99.

53.	Madico G, Welsch JA, Lewis LA, McNaughton A, Perlman DH, Costello CE, et al. The meningococcal vaccine candidate GNA1870 binds the complement regulatory protein factor H and enhances serum resistance. J Immunol. 2006;177(1):501-10.

54.	Schneider MC, Prosser BE, Caesar JJ, Kugelberg E, Li S, Zhang Q, et al. Neisseria meningitidis recruits factor H using protein mimicry of host carbohydrates. Nature. 2009;458(7240):890-3.

55.	Lucidarme J, Tan L, Exley RM, Findlow J, Borrow R, Tang CM. Characterization of Neisseria meningitidis isolates that do not express the virulence factor and vaccine antigen factor H binding protein. Clin Vaccine Immunol. 2011;18(6):1002-14.

56.	Giuliani M, Bartolini E, Galli B, Santini L, Lo Surdo P, Buricchi F, et al. Human protective response induced by meningococcus B vaccine is mediated by the synergy of multiple bactericidal epitopes. Sci Rep. 2018;8(1):3700.

57.	Laver JR, Gbesemete D, Dale AP, Pounce ZC, Webb CN, Roche EF, et al. A recombinant commensal bacteria elicits heterologous antigen-specific immune responses during pharyngeal carriage. Sci Transl Med. 2021;13(601).

58.	MHRA. Fluoroquinolone antibiotics: must now only be prescribed when other commonly recommended antibiotics are inappropriate. 2024.

59.	UKHSA. Guidance for public health management of meningococcal disease in the UK. 2024.

60.	Summary of medicinal product characteristics: Ciprofloxacin hydrochloride. 2024.

61.	Parikh SR, Andrews NJ, Beebeejaun K, Campbell H, Ribeiro S, Ward C, et al. Effectiveness and impact of a reduced infant schedule of 4CMenB vaccine against group B meningococcal disease in England: a national observational cohort study. Lancet. 2016;388(10061):2775-82.
